# Sleep-Stage Dynamics Predict Current Sleep-Disordered Breathing and Future Cardiovascular Risk

**DOI:** 10.1101/2025.07.31.25332545

**Authors:** Michal Bechny, Akifumi Kishi, Yasuhiro Tomita, Marco Scutari, Julia van der Meer, Markus Schmidt, Claudio Bassetti, Athina Tzovara, Francesca Faraci

## Abstract

Sleep-disordered breathing (SDB) is a major contributor to cardiovascular morbidity and disrupts both the macrostructure and dynamics of sleep stages (W, N1, N2, N3, REM). While specific alterations in sleep macrostructure, such as reduced durations of N3 and REM, have been linked to cardiovascular risk, the predictive value of sleep-stage dynamics remains unexplored. Using data from the prospective Sleep Heart Health Study, we applied a flexible forest-based modelling approach to a carefully selected cohort of 2579 subjects free from prior cardiovascular events and sleep-altering medications to minimize confounding. First, we demonstrate that a random forest classifier reliably identifies moderate-to-severe SDB (apnea–hypopnea index; AHI >15), achieving AUROC=76.1%, from sleep-stage architecture, dynamics, and common risk factors (demographics, BMI, smoking status) alone, without direct respiratory measurements. This highlights a dependency chain in which SDB correlates with altered sleep patterns that, in turn, encode cardiovascular risk. Second, a random survival forest robustly predicted future cardiovascular events (concordance-index=73.3%) over >10 years follow-up. Comparable results with and without including AHI as a predictor indicate that sleep patterns encode cardiovascular risk independently of direct SDB measurement. Partial dependence analyses revealed monotonic SDB risk profiles and predominantly U-shaped associations for cardiovascular risk, identifying ranges of total sleep time, wake after sleep onset, and REM/N3 continuity linked to minimal or elevated risk. Notably, rare transitions such as N3 →N1 or REM →N3, even occurring once per night, emerged as sensitive markers of cardiovascular vulnerability, increasing risk by up to 10%. Our findings extend prior evidence on linear associations between sleep macrostructure and cardiovascular outcomes, revealing non-linear patterns and positioning sleep dynamics as promising non-invasive biomarkers for diagnostics and early risk stratification.

## Introduction

Sleep-disordered breathing (SDB), particularly obstructive sleep apnea (OSA), is a well-established contributor to both cardiovascular morbidity^1–3^ and mortality^4–6^. The pathophysiology of SDB involves intermittent hypoxia, intrathoracic pressure swings, and repeated arousals, which activate the sympathetic nervous system, trigger oxidative stress, and induce systemic inflammation and endothelial dysfunction—processes that collectively accelerate vascular remodelling and atherogenesis^2,3,7,8^. In parallel, SDB promotes metabolic dysregulation through impaired insulin sensitivity and altered adipokine signalling, contributing to obesity and further elevating cardiovascular risk^2,8^. These mechanisms are reflected in many population-based studies. For example, moderate-to-severe SDB, defined by an apnea–hypopnea index (AHI) greater than 15, capturing the hourly rate of partial or complete breath arrests, was associated with a nearly 3-fold increased risk of incident ischemic stroke in men^1^. Similarly, SDB was prospectively linked with incident hypertension over four years, with over a 2-fold increased risk observed in those with AHI *≥* 5, independent of obesity and baseline blood pressure^9^.

Beyond its contribution to cardiovascular morbidity, SDB is also associated with increased risks of all-cause and cardiovascular mortality, with nearly double the risk of stroke or death compared to individuals without the condition, even after adjusting for major confounders^4^. Severe SDB has also been linked to a 46% higher risk of all-cause mortality over long-term follow-up^5^. Importantly, SDB may alter the temporal distribution of deaths: sudden cardiac death in individuals with OSA is more likely to occur during the night, particularly between midnight and 6 AM, in contrast to the early morning peak seen in the general population^10^. A severe SDB has been associated with a 2.5-fold increased risk of sudden cardiac death^6^.

Beyond respiratory disturbances, SDB leads to marked alterations in sleep macro-architecture, described by five sleep–wake states^11^: wakefulness (W), rapid eye movement (REM) sleep, and three non-REM (NREM) stages: light (N1, N2) and deep slow-wave (N3) sleep, each representing a distinct physiological state^12,13^. Specifically, SDB is associated with reduced proportions of N3 and REM sleep, critical for physical and cognitive restoration, respectively, as well as increased time in wake after sleep onset (WASO), and frequent awakenings and micro-arousals that fragment sleep continuity^14–16^. These changes lead to lighter, less efficient sleep dominated by N1 and N2 stages, which contributes to daytime symptoms and heightened sympathetic activity—an aspect of autonomic imbalance linked to cardiovascular risk^14,15^. These macro-structural abnormalities may serve as both markers and mediators of downstream cardiometabolic dysfunction.

In addition to altered stage composition, SDB disrupts the temporal continuity and organization of sleep stages—a feature referred to as sleep-stage dynamics^17–20^. The dynamics describe how individuals transition between stages over time and may capture subtle signatures of sleep instability that are not evident in static macrostructure metrics such as stage proportions or durations. The dynamic patterns have been shown to reflect diverse physiological and pathological conditions beyond SDB^21–29^. Importantly, specific sleep-stage transitions may reflect underlying physiological needs: frequent transitions into deep slow-wave sleep (N3) signal elevated homeostatic sleep pressure and physical restoration processes^13,30^, while disrupted or shortened REM periods signal impaired cognitive and emotional recovery^31–33^. Prior work has shown that SDB-individuals exhibit irregular and less predictable sleep-stage transitions, accompanied by abnormal heart rate variability patterns^17^. Transition-based modelling approaches have revealed elevated transition entropy and reduced stage persistence, indicating a loss of normal sleep structure in SDB subjects^18^. Recent analyses have further demonstrated that these dynamic patterns vary systematically by age, gender, and apnea severity, with distinct fragmentation profiles across REM and NREM stages^19,20^.

Notably, disrupted sleep macro-architecture has been linked to increased cardiovascular risk and mortality. Reduced slow-wave sleep (N3) is associated with incident hypertension, possibly reflecting impaired nocturnal blood pressure regulation^34,35^, while lower REM sleep has been linked to higher all-cause and cardiovascular mortality^36^. Diminished delta wave activity during sleep, related to reduced N3, has also been associated with long-term cardiovascular outcomes^37^. Abnormal total sleep duration, as well as poor self-reported sleep quality, show associations with cardiovascular and all-cause mortality^38–41^.

Although disrupted sleep macro-architecture has been repeatedly associated with cardiovascular outcomes and mortality^7,8,34–41^, the prognostic relevance of sleep-stage dynamics—able to capture detailed physiological signatures—remains unexplored. Since SDB affects both cardiovascular outcomes^1–6,9^ and sleep-stage dynamics^17–20^, these two domains are statistically linked, suggesting that sleep dynamics may encode predictive signals relevant for cardiovascular risk modelling. Furthermore, dynamic sleep patterns have been associated with a range of other conditions, including insomnia, chronic fatigue syndrome, pain syndromes (fibromyalgia), sleep bruxism, and neurocognitive impairment^21–29^, which may also contribute to cardiovascular vulnerability. This interplay highlights the potential of sleep dynamics as integrative, non-invasive digital markers for diagnostics and long-term cardiovascular risk assessment.

### Study contributions and research question

We present the first investigation to evaluate whether and eventually how sleep-stage dynamics, alongside conventional sleep macrostructure metrics and common risk factors (demographics, BMI, smoking status), carry prognostic value for long-term cardiovascular outcomes. Using data from the prospective, longitudinal, community-based Sleep Heart Health Study (SHHS)^42^, we define a primary analysis cohort consisting of individuals without prior cardiovascular events and free from medications altering sleep architecture or cardiovascular physiology (e.g., antidepressants, beta-blockers, diuretics, aspirin). This design reduces confounding and enhances the generalizability of our findings to broader populations. For modelling, we adopt forest-based approaches^43–46^, which offer a flexible non-parametric framework robust to overfitting, variable interactions, multicollinearity, and non-linear effects. We consider four groups of predictors, capturing static and dynamic properties of sleep and relevant risk factors: (i) *percentages of sleep-stage transitions* (e.g., W → N1) relative to the time after sleep onset characterizing sleep-stage dynamics; (ii) *conventional sleep metrics* (e.g., total sleep time [TST], WASO); (iii) *demographics* (age, gender); and known (iv) *risk factors* including body mass index (BMI) and smoking status.

Our study presents four key contributions:

1. **Identification of current SDB status:** We apply a Random Forest (RF) classifier to identify individuals with moderate-to-severe SDB (AHI *>* 15) and demonstrate that characteristic changes in sleep architecture and dynamics carry diagnostic information about underlying respiratory disturbance, and hence, a possible link to long-term cardiovascular health.
2. **Prediction of long-term cardiovascular risk:** We use a Random Survival Forest (RSF) to estimate long-term cardiovascular risk. By comparing models with and without an AHI predictor characterizing the SDB severity, we assess whether the four sets of predictors alone can encode the prognostic information attributable to SDB.
3. **Validation and generalizability:** We assess the RF and RSF models through cross-validation within the primary study cohort and across additional SHHS subgroups, including baseline or follow-up polysomnography (PSG) data from individuals with prior cardiovascular events or medication use. A further validation is performed using the external *Bern Sleep-Wake Registry* (BSWR). In BSWR, SDB predictions are directly evaluated, while cardiovascular risk estimates are regressed against various sleep disorders and other comorbidities.
4. **Interpretability and novel markers via partial effects:** We use partial dependence plots from the R(S)F models to assess how individual predictors influence both SDB and cardiovascular risk, revealing non-linear effects, candidate thresholds for clinical risk-stratification, and suggesting novel diagnostic and prognostic markers.

## Results

The *Results* section is organized following our main objectives: *(i) Descriptive statistics*, summarizing demographics, SDB prevalence, and clinically established sleep markers, stratified by the occurrence of future cardiovascular events in the primary study cohort used for model development; *(ii) Performance of the RF classifier* for identifying current moderate-to-severe SDB (AHI >15); *(iii) Performance of RSF for predicting cardiovascular risk*, assessed both with and without inclusion of the AHI predictor. Sections (ii) and (iii) also report the partial effects of individual predictors to investigate their risk associations, and evaluate model performance across SHHS test subgroups and the external BSWR data set.

### Descriptive statistics

Table 1 summarizes demographic characteristics, SDB prevalence and severity, and sleep macro-structure metrics for the primary SHHS1(E = 0, M = 0) cohort, which excludes individuals with prior cardiovascular events (E = 1) or sleep-altering medications (M = 1). Statistics are presented for the entire cohort (*overall*) and stratified by the occurrence of a cardiovascular event during the follow-up period of up to 15 years. Cardiovascular events refer to a pooled composite outcome, including diagnoses such as stroke, myocardial infarction, heart failure, or surgical procedures such as coronary revascularisation (cf. *Data sets*).

**Table 1.** Descriptive characteristics of SHHS1 (E = 0, M = 0) cohort stratified by cardiovascular event status. Continuous variables are reported as mean (SD) and compared using Welch’s two-sample *t*-test. Categorical variables, denoted by superscript ^*^, are reported as counts (percentages) and compared using the chi-squared test. When expected cell counts were less than 5, Fisher’s exact test was used instead.

| Variable | Overall | Event-free | Event developed | p-value |
| --- | --- | --- | --- | --- |
| N | 2579 | 2253 | 326 |  |
| Age | 59.43 (11.16) | 58.11 (10.59) | 68.57 (10.73) | <0.001 |
| Gender (Male)* | 1182 (45.8) | 1001 (44.4) | 181 (55.5) | <0.001 |
| Smoking* |  |  |  | 0.041 |
| Current | 281 (10.9) | 235 (10.4) | 46 (14.1) |  |
| Ex | 1010 (39.2) | 874 (38.8) | 136 (41.7) |  |
| Never | 1259 (48.8) | 1116 (49.5) | 143 (43.9) |  |
| NA | 29 (1.1) | 28 (1.2) | 1 (0.3) |  |
| BMI | 27.73 (4.92) | 27.66 (4.96) | 28.21 (4.63) | 0.062 |
| AHI | 16.00 (14.76) | 15.66 (14.59) | 18.34 (15.71) | 0.002 |
| SDB (AHI>15)* | 999 (38.7) | 844 (37.5) | 155 (47.5) | 0.001 |
| SDB category* |  |  |  | 0.012 |
| Mixed | 429 (16.6) | 362 (16.1) | 67 (20.6) |  |
| NREM-dominant | 78 (3.0) | 68 (3.0) | 10 (3.1) |  |
| REM-dominant | 369 (14.3) | 310 (13.8) | 59 (18.1) |  |
| AHI≤15 | 1580 (61.3) | 1409 (62.5) | 171 (52.5) |  |
| NA | 123 (4.8) | 104 (4.6) | 19 (5.8) |  |
| TST [mins] | 365.10 (63.93) | 366.36 (63.09) | 356.33 (68.90) | 0.008 |
| WASO [mins] | 90.14 (54.66) | 88.46 (53.80) | 101.74 (59.09) | <0.001 |
| SE [%] | 72.29 (12.01) | 72.53 (11.79) | 70.64 (13.34) | 0.008 |
| SL [mins] | 50.63 (43.05) | 51.11 (43.18) | 47.31 (42.00) | 0.136 |
| REML [mins] | 104.81 (140.70) | 104.30 (136.67) | 108.35 (166.06) | 0.627 |
| DL [mins] | 60.90 (185.96) | 58.67 (182.37) | 76.32 (208.77) | 0.109 |
| W [%] | 19.65 (11.57) | 19.28 (11.31) | 22.19 (12.95) | <0.001 |
| N1 [%] | 4.02 (2.84) | 3.99 (2.81) | 4.20 (3.05) | 0.200 |
| N2 [%] | 45.34 (11.43) | 45.28 (11.26) | 45.78 (12.52) | 0.455 |
| N3 [%] | 14.70 (9.50) | 14.95 (9.49) | 12.93 (9.35) | <0.001 |
| REM [%] | 16.30 (6.13) | 16.50 (6.10) | 14.89 (6.14) | <0.001 |

Out of the 2579 subjects, 326 experienced at least one cardiovascular event during follow-up. When compared to subjects who did not develop the event, these individuals were significantly older at baseline (mean age 68.6 vs. 58.1 years), more likely to be male (55.5% vs. 44.4%), and had a different composition of smoking status profiles (e.g., 14.1% vs. 10.4% current smokers, and 43.9% vs. 49.5% never smokers). BMI did not differ between the compared groups, potentially due to group differences in demographics and their interactions with smoking status. These demographic and lifestyle differences are consistent with established cardiovascular risk factors^3,9,47–50^.

Subjects who developed events had higher mean AHI values (18.3 vs. 15.7 events/hour) and greater prevalence of moderate-to-severe SDB (47.5% vs. 37.5%), reflecting the link between SDB and cardiovascular risk^1–8^. However, since AHI increases with age, this association may also reflect age distribution differences^47,48^.

Sleep macrostructure also differed: those who developed events had shorter TST and longer WASO, resulting in lower sleep efficiency 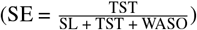. They also showed altered sleep-stage composition, with higher W%, and reduced N3 and REM%—stages critical for physical and cognitive restoration. These changes likely reflect both elevated cardiovascular risk and age- or SDB-related alterations of sleep structure.

These statistics provide a descriptive overview of group-level trends and are not intended as clinically conclusive findings. Rather, they highlight the need for a more flexible modelling approach that can simultaneously account for multiple confounding variables, capture their interactions, and accommodate potential non-linear effects. Accordingly, in the following sections, we employ Random Forest and Random Survival Forest models to numerically isolate the contributions of key risk factors—such as age, smoking, BMI, and SDB severity—to cardiovascular outcomes and sleep-related metrics.

### Identification of current SDB status

#### Predictors and training of RF

To identify the presence of SDB, we trained an RF classifier using a total of 34 predictors: 25 sleep-stage transition proportions capturing sleep dynamics; 5 conventional sleep macrostructure metrics not encoded in dynamics (TST, WASO, and latencies to sleep onset, N3, and REM); and 4 commonly recognized risk factors accounting for demographic and lifestyle variation (age, gender, BMI, and smoking status). We did not include SE, as it is a function of TST, WASO, and SL. The binary outcome label indicated moderate-to-severe SDB (positive class, AHI*>*15) versus no-or-mild SDB (negative class, AHI ≤ 15), supported by evidence linking AHI*>*15 to substantially elevated risks of stroke, cardiovascular morbidity, and mortality^1,5,6^.

Given the imbalance between 38.7% of positive and 61.3% of negative cases, we used a quantile-based RF implemented in `randomForestSRC` R package^51,52^, effectively mitigating class dominance. To quantify the uncertainty in performance metrics (e.g., AUROC, accuracy), we performed repeated training using 5-fold cross-validation on anticluster-based data splits (see *Methods*) of the primary cohort SHHS1(E = 0, M = 0). For generalization assessments on the remaining SHHS subsets and BSWR, and the interpretation of partial effects, a final RF model was estimated on the complete set of 2579 subjects.

All RF models used default hyperparameters: a minimum terminal node size of 1, 10 candidate split points per variable, an AUROC-based splitting criterion, and the square root of the number of considered predictors as the number of variables to try to split each node. Each forest consisted of 1000 trees, and missing input values were imputed using the built-in out-of-bag terminal node imputation algorithm^46,51^.

#### Performance and generalization of RF

The first part of Table 2 reports the mean (standard deviation) performance metrics in the primary cohort SHHS1(E=0, M=0), assessed using cross-validation. Across five anticluster splits, the data set contained approximately 316 controls (AHI≤15) and 200 positive cases (AHI*>*15) on average. The RF classifier demonstrated strong discriminative ability, achieving an AUROC of 76.1 (2.2)% for distinguishing moderate-to-severe SDB cases from healthy-to-mild controls based on sleep parameters (i.e., transition %’s, macrostructure) combined with other variables (demographics, BMI, smoking status) alone. The strong performance was further supported by a strong positive correlation of 0.45 (0.05) between the predicted probabilities of the positive class and the subject’s actual AHI values. The RF achieved an accuracy of 70.0 (1.6)%, sensitivity of 46.8 (4.6)%, specificity of 84.7 (1.1)%, and precision of 65.9 (1.9)%. Considering that SDB is typically quantified using respiratory and oxygen desaturation signals, the obtained performance metrics indicate a favourable balance between case detection and a low false positive rate.

**Table 2.**
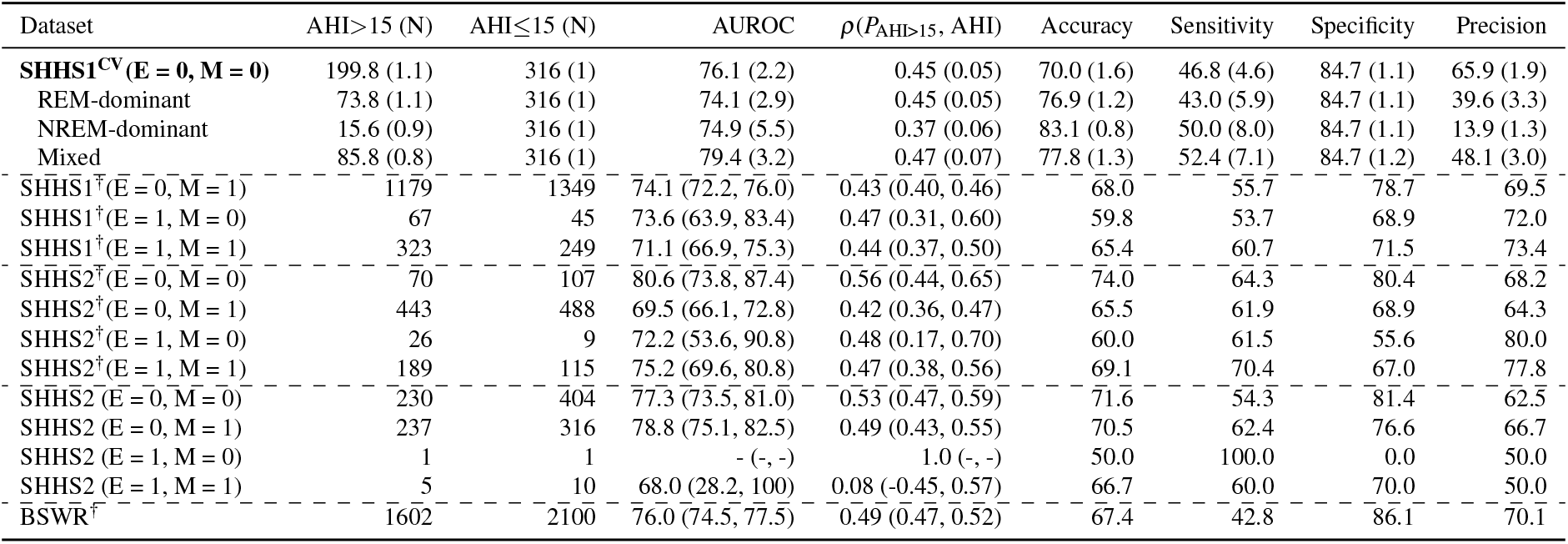
Identification of moderate-to-severe sleep-disordered breathing (AHI*>*15) in different testing datasets of subjects with previous cardiovascular events (E = 1) or medications (M = 1) from baseline (SHHS1) and follow-up (SHHS2) Sleep Heart Health Study and Berner Sleep-Wake Registry (BSWR). Cross-validation assessment in the primary study cohort is presented as mean (standard deviation) and highlighted by bold font and ^CV^ superscript. Datasets containing out-of-domain subjects are highlighted by ^†^ superscript. The table presents numbers (N) of cases (AHI*>*15) and controls (AHI≤15), 95% confidence intervals for Area Under the Receiver Operating Characteristic (AUROC) and Pearson’s correlation coefficient of predicted probability of positive class (*P*_AHI>15_) with actual AHI value, and the achieved performance metrics (%).

| Dataset | AHI > 15 (N) | AHI ≤ 15 (N) | AUROC | $\rho(P_{\text{AHI} > 15}, \text{AHI})$ | Accuracy | Sensitivity | Specificity | Precision |
| --- | --- | --- | --- | --- | --- | --- | --- | --- |
| **SHHS1<sup>CV</sup>(E = 0, M = 0)** | 199.8 (1.1) | 316 (1) | 76.1 (2.2) | 0.45 (0.05) | 70.0 (1.6) | 46.8 (4.6) | 84.7 (1.1) | 65.9 (1.9) |
| REM-dominant | 73.8 (1.1) | 316 (1) | 74.1 (2.9) | 0.45 (0.05) | 76.9 (1.2) | 43.0 (5.9) | 84.7 (1.1) | 39.6 (3.3) |
| NREM-dominant | 15.6 (0.9) | 316 (1) | 74.9 (5.5) | 0.37 (0.06) | 83.1 (0.8) | 50.0 (8.0) | 84.7 (1.1) | 13.9 (1.3) |
| Mixed | 85.8 (0.8) | 316 (1) | 79.4 (3.2) | 0.47 (0.07) | 77.8 (1.3) | 52.4 (7.1) | 84.7 (1.2) | 48.1 (3.0) |
| SHHS1<sup>†</sup>(E = 0, M = 1) | 1179 | 1349 | 74.1 (72.2, 76.0) | 0.43 (0.40, 0.46) | 68.0 | 55.7 | 78.7 | 69.5 |
| SHHS1<sup>†</sup>(E = 1, M = 0) | 67 | 45 | 73.6 (63.9, 83.4) | 0.47 (0.31, 0.60) | 59.8 | 53.7 | 68.9 | 72.0 |
| SHHS1<sup>†</sup>(E = 1, M = 1) | 323 | 249 | 71.1 (66.9, 75.3) | 0.44 (0.37, 0.50) | 65.4 | 60.7 | 71.5 | 73.4 |
| SHHS2<sup>†</sup>(E = 0, M = 0) | 70 | 107 | 80.6 (73.8, 87.4) | 0.56 (0.44, 0.65) | 74.0 | 64.3 | 80.4 | 68.2 |
| SHHS2<sup>†</sup>(E = 0, M = 1) | 443 | 488 | 69.5 (66.1, 72.8) | 0.42 (0.36, 0.47) | 65.5 | 61.9 | 68.9 | 64.3 |
| SHHS2<sup>†</sup>(E = 1, M = 0) | 26 | 9 | 72.2 (53.6, 90.8) | 0.48 (0.17, 0.70) | 60.0 | 61.5 | 55.6 | 80.0 |
| SHHS2<sup>†</sup>(E = 1, M = 1) | 189 | 115 | 75.2 (69.6, 80.8) | 0.47 (0.38, 0.56) | 69.1 | 70.4 | 67.0 | 77.8 |
| SHHS2(E = 0, M = 0) | 230 | 404 | 77.3 (73.5, 81.0) | 0.53 (0.47, 0.59) | 71.6 | 54.3 | 81.4 | 62.5 |
| SHHS2(E = 0, M = 1) | 237 | 316 | 78.8 (75.1, 82.5) | 0.49 (0.43, 0.55) | 70.5 | 62.4 | 76.6 | 66.7 |
| SHHS2(E = 1, M = 0) | 1 | 1 | - (-, -) | 1.0 (-, -) | 50.0 | 100.0 | 0.0 | 50.0 |
| SHHS2(E = 1, M = 1) | 5 | 10 | 68.0 (28.2, 100) | 0.08 (-0.45, 0.57) | 66.7 | 60.0 | 70.0 | 50.0 |
| BSWR<sup>†</sup> | 1602 | 2100 | 76.0 (74.5, 77.5) | 0.49 (0.47, 0.52) | 67.4 | 42.8 | 86.1 | 70.1 |

Table 2 also presents performance metrics for different SDB phenotypes: *REM-dominant* (AHI*>*15 in REM sleep only; ∼74 cases), *NREM-dominant* (AHI*>*15 in non-REM sleep only; ∼16 cases), and *Mixed* (AHI ∼15 in both REM and NREM sleep; ∼86 cases). Although the original RF was optimized for overall SDB discrimination (i.e., AHI*>*15 vs. AHI≤15), the AUROC remained high across these subgroups: approximately 74% for REM- and NREM-dominant SDB and 79.4% for *Mixed* cases. Other performance metrics were similarly robust, with a drop in precision for NREM-dominant SDB due to the small number of cases.

Next, Table 2 provides 95% confidence intervals (CIs) for AUROC and correlation coefficients, along with performance metrics for additional test subsets from the SHHS1 baseline study (including participants on medications or with prior cardiovascular events) and the SHHS2 follow-up. Stratification further considered whether PSG recordings came from previously unseen (out-of-domain) individuals, not included in RF training, highlighted by the ^†^ superscript. The results indicate that SDB detection remained robust even for SHHS1 participants with medications or prior events, achieving AUROC values >71% in all cases. Similar performance was observed across all subsets in the follow-up study: for the SHHS2 participants without medications or prior events used in RF training, SHHS2(E = 0, M = 0), AUROC reached 77.3%, and in unseen test follow-up medication- and event-free participants, SHHS2^†^(E = 0, M = 0), performance further improved to 80.6%, underscoring the model’s strong generalizability and robustness to domain shifts, as the follow-up participants are naturally older from their baseline assessment. The performance on all test strata is particularly strong, given that post-event or medicated individuals exhibit altered sleep patterns^53–56^, and these cohorts are on average older (cf. Supplementary Tables 4-11).

The RF maintained robust performance in the out-of-domain BSWR (Supplementary Tables 1-2), which primarily includes patients with diverse sleep disorders rather than a general population sample, as in SHHS. Despite the altered sleep patterns expected in BSWR, the RF achieved an AUROC of 76.0%, a correlation of 0.49 between predicted probability and AHI, an accuracy of 67.4%, sensitivity of 42.8%, specificity of 86.1%, and a precision of 70.1%. These results highlight strong discriminative ability and high precision, indicating that individuals with high predicted probabilities are frequently true SDB cases.

### SDB risk-profiles via partial effects of RF

As described in *Methods*, the trained RF enables quantification of partial effects for individual predictors *X*_*j*_. These effects mathematically represent the change in the RF output (i.e., the predicted probability of moderate-to-severe SDB) when iterating over all (or, for computational efficiency, a subset of) observed values of *X*_*j*_, substituting these and averaging predictions across all *N* observations in the data set while holding other variables fixed. As *N* predicted outcomes are obtained for each considered value of *X*_*j*_, the expected partial effect is estimated as the mean, and uncertainty is expressed using quantile-based confidence intervals computed around this estimate. Plotting partial effects and their associated intervals yields a “risk profile” that shows how variations in *X*_*j*_ relate to the RF-predicted SDB-probability.

Figure 1 illustrates the partial effects of demographic (age, gender) and lifestyle (BMI, smoking) variables on the probability of moderate-to-severe SDB (AHI≥15). Both age and BMI exhibit sigmoidal profiles, reflecting an accelerated risk increase within specific ranges before reaching a plateau. For age, the predicted SDB probability rises sharply from 33% to 45% between 45 and 70 years. A similar, but even steeper, trend is observed for BMI: the risk increases from about 30% at BMI ≤25 to *>*50% for BMI*>*35. Partial effects further indicate a 6% higher predicted risk in males (36% in females vs. 42% in males). These associations are consistent with established age, obesity, and gender differences in SDB prevalence^47,48,57^. Interestingly, ex-smokers show a 1% higher predicted risk compared to both current and never-smokers, potentially reflecting post-cessation weight gain^49,50^ or underlying health issues prompting cessation. It is important to note that these partial effects are contingent upon the population characteristics of the RF training cohort. In particular, the SHHS design oversampled individuals with snoring to increase statistical power for long-term cardiovascular outcomes^42^. Consequently, baseline SDB risk estimates (e.g., for individuals in their early 40s) may be inflated relative to the general population.

**Figure 1.**
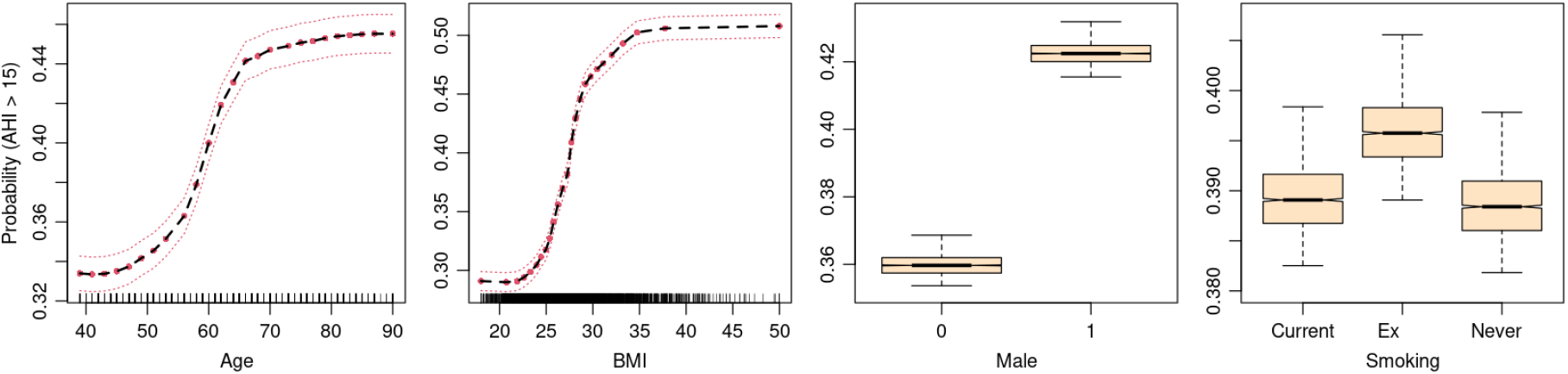
Partial effects and their 95% CIs for the risk of moderate-to-severe sleep-disordered breathing (AHI*>*15) for the age in years, Body Mass Index (BMI), gender (0 = female, 1 = male), and smoking status. Data points for continuous predictors are shown as ticks on the x-axis.

Figure 2 shows the partial effects of sleep macro-architecture markers used as RF inputs. The SDB risk increases markedly when TST falls below 300 minutes (5 hours) and when WASO exceeds 100 minutes, reflecting reduced sleep efficiency and prolonged wakefulness, likely due to apnea-related arousals. In contrast, the sleep-onset latency (SL) shows no clear association with SDB risk, whereas delays in entering REM sleep (REML) and deep sleep (DL) beyond 100 minutes are associated with an approximate 2% increase in risk.

**Figure 2.**
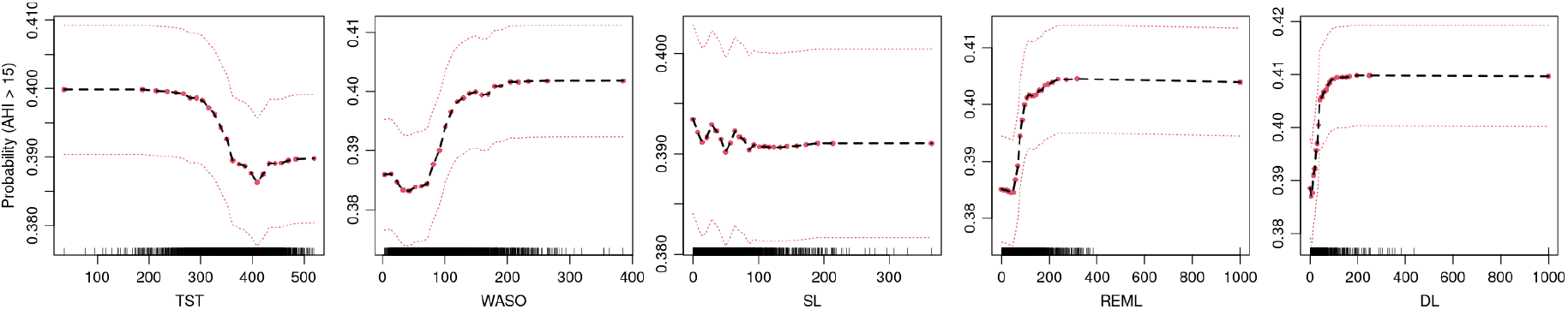
Partial effects and their 95% CIs for the risk of moderate-to-severe sleep-disordered breathing (AHI*>*15) for the minutes of Total Sleep Time (TST), Wake After Sleep Onset (WASO), Sleep Latency (SL), REM Latency (REM), and Deep-sleep Latency (DL). Data points are shown as ticks on the x-axis.

Figure 3 details partial effects for sleep-stage transition proportions *p*_*i, j*_ (cf. *Methods*), computed relative to the total number of sleep-stages from sleep onset to the end of the PSG recording. As shown in Table 1, the average after-onset PSG-duration (sleep period time) of 455.2 minutes (TST + WASO = 365.10 + 90.14) corresponds to approximately 910 epochs, such that a 1% (= 0.01) change in transition proportion corresponds to roughly 9 transitions per PSG. For interpretability, we focus on transitions associated with ≥2% change in predicted risk, supported by non-overlapping deviations in 95% CI profiles of their partial effects.

- *W-transitions*: The SDB risk increases by ∼2% when *p*_*W,N*1_ *>* 0.04 (∼36 transitions), reflecting frequent awakenings followed by light sleep in SDB subjects. Similarly, *p*_*W,N*2_ *>* 0.02 (∼18 transitions) is associated with *>*2% higher risk, suggesting that SDB subjects experience heightened homeostatic sleep pressure and atypically bypass the intermediate N1 stage after awakenings. Notably, *p*_*W,REM*_ *>* 0.08 corresponds to a dramatic risk increase (*>*5%), indicating that direct transitions from wake to REM, possibly related to higher restorative pressure or REM sleep fragmentation, are highly sensitive markers of SDB.
- *N1-transitions*: The SDB-risk increases by *>*6% when *p*_*N*1,*W*_ *>* 0.01 (∼9 transitions), suggesting frequent arousals of SDB subjects from lightest sleep. Notably, *p*_*N*1,*REM*_ *>* 0.005 (∼4 transitions) is linked to a ∼2% higher risk, possibly reflecting atypical transitions to REM from N1 in SDB patients, similar to those from W.
- *N2-transitions*: Higher proportions of *p*_*N*2,*W*_ *>* 0.03 (∼27 transitions) correspond to a ∼8% SDB-risk increase, indicative of frequent awakenings from N2, likely linked to disruptions driven by experienced apnea events.
- *N3-transitions*: A SDB-risk increase ∼3% is observed when *p*_*N*3,*N*3_ *<* 0.1, corresponding to ∼90 uninterrupted epochs of N3, suggesting that SDB subjects are often unable to achieve more than 45 minutes of continuous deep sleep over the entire night.
- *REM-transitions*: The SDB-risk rises by ∼3% when *p*_*REM,W*_ *>* 0.015 (∼13 transitions), highlighting increased REM-awakenings in SDB. In contrast, higher persistence within REM (*p*_*REM,REM*_ *>* 0.2, ∼180 epochs, = 1.5 hours) links to a lower risk, suggesting difficulties in retaining uninterrupted REM-sleep in SDB.

**Figure 3.**
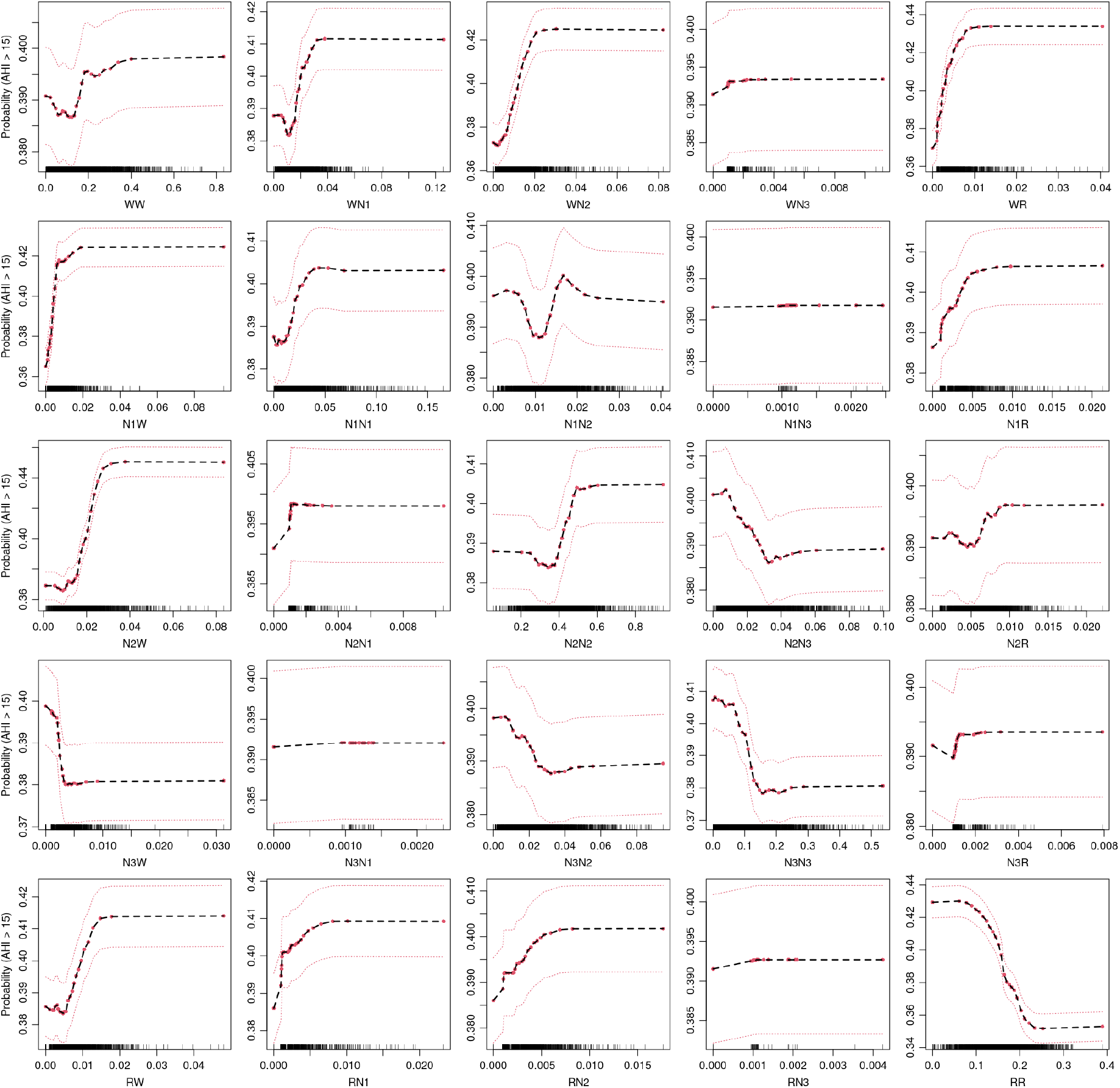
Partial effects and their 95% CIs for the risk of moderate-to-severe sleep-disordered breathing (AHI*>*15) for relative frequencies of individual transitions between sleep-stages (W, N1, N2, N3, REM = R). Each subplot’s x-axis label indicates the direction of the transition (e.g., WN1 corresponds to transitions from W to N1). Data points are shown as ticks on the x-axis.

The estimated partial effects mechanistically explain RF decisions when identifying SBD. In addition, they characterize risk profiles, which may suggest thresholds that can be used as clinical instruments (markers) for diagnostics and risk stratification.

### Prediction of long-term cardiovascular risk

#### Predictors and training of RSF

To quantify the risk of future cardiovascular events, we trained a RSF using the same 34 predictors as for RF, including 25 sleep-stage transition proportions, 5 sleep macro-structure metrics (TST, WASO, and latencies to sleep onset, N3, and REM), and 4 demographic and lifestyle variables (age, gender, BMI, and smoking status). To evaluate the added prognostic value of including a direct measure of SDB, we trained RSF models both with and without apnea–hypopnea index (AHI) as a predictor. As described in *Methods*, the RSF target was a survival outcome comprising an event indicator (1 if the participant experienced a cardiovascular event after the sleep study, 0 otherwise) and the time-to-event, defined as the number of days since the PSG study to either the first event or the most recent follow-up, if censored. The RSF aimed to map the links between survival outcomes and the included predictors, and to quantify their influence via partial dependence analysis subsequently.

The RSF was trained using `randomForestSRC` R package^51,52^ on the primary cohort SHHS1(E = 0, M = 0), to minimize confounding and maximize the generalizability to a general, medications- and prior-event-free population. To assess uncertainty in predictive performance and discrimination metrics (e.g., time-dependent AUROC [tdAUROC], Integrated Brier Score [IBS], Harrell’s concordance index [C-index], log-rank test), we applied a 5-fold cross-validation with anticluster-based splits (*see Methods*). For generalization assessments on remaining SHHS subsets and BSWR, and interpretation of partial effects, a final RSF was trained on the complete set of 2579 subjects.

During training, default RSF hyperparameters were used: a minimum terminal node size of 15, 10 candidate split points per variable, C-index splitting criterion, and the square root of the number of predictors as the number of variables to try to split each node. Each RSF comprised 1000 trees, and missing input values were imputed using the built-in out-of-bag terminal node imputation algorithm^46^.

#### Performance and generalization of RSF

Table 3 and Supplementary Table 12 summarize the performance and discrimination metrics of RSF trained while including and omitting the AHI as a predictor, respectively. Results are shown for the primary study cohort and for subjects across SHHS1 and SHHS2 subsets who were free of prior cardiovascular events at the time of PSG assessment. The first column of each table, labelled SHHS1^CV^(E = 0, M = 0), reports cross-validation results within the primary cohort, expressed as a mean (standard deviation) for each metric. Each anticluster fold comprised, on average, about 516 subjects, of whom 65 developed a cardiovascular event during follow-up and 451 remained event-free. The RSF’s ability to rank individuals by their actual cardiovascular risk was evaluated using three standard metrics: C-index, IBS, and tdAUROC. For C-index and tdAUROC, values of 100% indicate perfect discrimination, 50% random chance, and 0% inverted predictions. In contrast, lower IBS values indicate better calibration. For all metrics, cardiovascular risk was quantified via the RSF-predicted *mortality score*, representing an individual’s standardized risk relative to a hypothetical population of subjects of matched characteristics^51,52^.

**Table 3.**
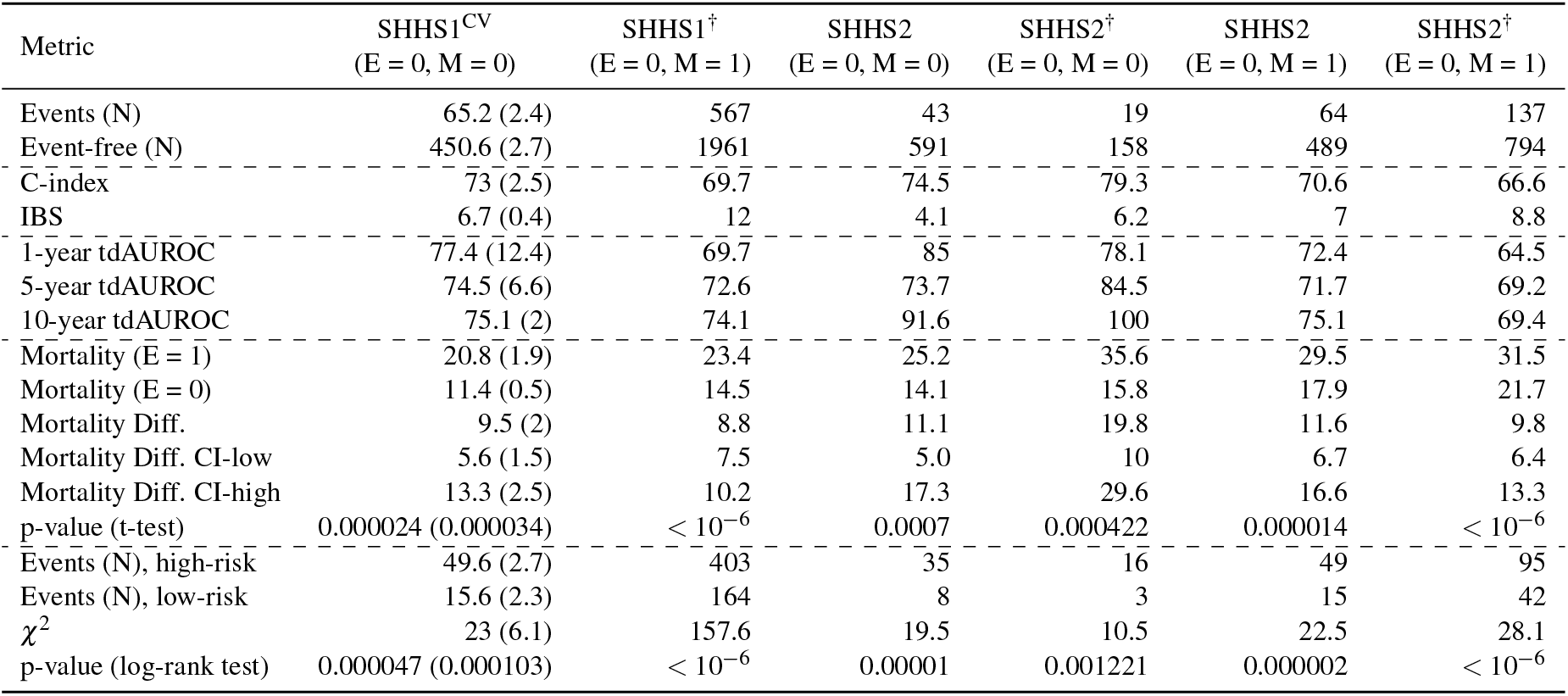
Performance of the Random Survival Forest (RSF) model including AHI predictor, evaluated across various datasets of subjects with no previous cardiovascular events (E = 0). The ^CV^ superscript denotes performance obtained via 5-fold cross-validation (CV) on the in-domain event- and medication-free (E = M = 0) baseline cohort SHHS1(E = 0, M = 0). All other columns evaluate the performance of the final RSF model fitted to the entire baseline cohort, applied to potentially out-of-domain subjects (^†^) from either the baseline (SHHS1) or follow-up (SHHS2) studies, including subgroups taking medication (M = 1). For each scenario, the number of subjects with events and without events (event-free) is reported. Model performance is assessed using Harrell’s Concordance Index (C-index), Integrated Brier Score (IBS), and the time-dependent Area Under the Receiver Operating Characteristic curve (tdAUROC) at 1, 5, and 10 years. Discriminatory ability is evaluated via two-sided t-tests comparing predicted mortality between event and non-event subjects, including 95% confidence intervals (CI) for the difference (Diff.). Additionally, log-rank tests with Chi-squared (*χ*^2^) statistics compare event rates between high- and low-risk groups stratified by median predicted mortality. For the in-domain CV, mean (SD) of all metrics is reported.

Notably, a comparison of Table 3 and Supplementary Table 12 reveals no measurable performance gain from including AHI. The performance metrics achieved with vs. without AHI predictor were statistically identical: C-index at 73.0 (2.5) vs. 73.3 (2.5), IBS at 6.7 (0.4) for both cases, and the (1, 5, 10)-year tdAUROC at [77.4 (12.4), 74.5 (6.6), 75.1 (2.0)] vs. [77.1 (12.2), 74.9 (6.2), 75.3 (2.3)], respectively. Considering a 2*σ* bound around the mean as an indicator of significant difference, there is not a single case in which the two models would differ. Comparable performance between the two RSFs is further confirmed by a discriminatory assessment using a t-test, which compares the mean difference in predicted mortality between subjects who developed an event and those who did not, and a log-rank test comparing high- and low-risk individuals identified by the median threshold on predicted mortality. For RSF with and without AHI, the cross-validation-averaged mean differences were 9.5 (95% CI: 5.6–13.3, p-val ∼2.4 ×10^*−*5^) and 9.8 (95% CI: 5.8–13.7, p-val ∼1.3 × 10^*−*5^), respectively, whereas the log-rank *χ*^2^ statistics were 23 (p-val ∼4.7 × 10^*−*5^) and 26 (p-val ∼5 × 10^*−*6^), respectively. Finally, Supplementary Figures 1 and 13 illustrate the distribution of the primary study population concerning cardiovascular cases, survivors, and censoring, as well as the performance metrics (tdAUROC, IBS) for the RFS with and without AHI, respectively, yielding identical trends.

This robust finding suggests that, despite the well-known clinical evidence linking SDB to future cardiovascular events^1–8^, directly including AHI as a predictor does not provide any measurable improvement in model performance when common risk factors (demographics, BMI, smoking status), sleep macro-architecture and dynamics are already controlled for. This observation can be interpreted from two complementary perspectives. First, as shown in the previous experiment, the same set of predictors can effectively capture SDB-related patterns. It is therefore likely that correlations between SDB and cardiovascular risk are indirectly encoded in these features, allowing the RSF to leverage this information without requiring explicit measurement of AHI. Second, the onset of cardiovascular events may be influenced by broader alterations in sleep macrostructure and dynamics that extend beyond SDB-specific patterns. The RSF, being a flexible and highly capable modelling approach (1000 trees), is well-suited to detect such complex relationships. These could include subtle patterns reflecting the downstream effects of other “hidden” conditions, such as neuropsychiatric, metabolic, renal, or other comorbidities that might be simultaneously associated both with altered sleep patterns^58–60^ and an increased risk of cardiovascular events^61–64^.

Table 3 and Supplementary Table 12 further present the generalization performance of the final RSF with and without the AHI predictor, respectively, on remaining test cohorts of subjects without prior cardiovascular events. The results are stratified by medication use (M = 1) and indication of out-of-domain subjects (^†^), unseen by RSF during their training. In all scenarios, the models with and without AHI achieved stable and comparable performance across cohorts, with C-index values ranging 66.6–79.3% and 66.6–78.3%, IBS values 4.1–12% and 4.1–11.9%, 1-year tdAUROC 64.5–85% and 64.1–85.2%, 5-year tdAUROC 69.2–84.5% and 69.2–83.4%, and 10-year tdAUROC 69.4–100% and 69–100%, respectively. Strong discriminative performance was further supported by significant differences in predicted mortality between event-free individuals and those who developed cardiovascular events, as determined by a t-test and a log-rank test comparing high- and low-risk groups, in both RSF models and all scenarios. This suggests that RSF performance in subjects without prior events is robust to domain shifts, as medication-taking subjects are typically older, and to potential alterations in sleep induced by medications.

Supplementary Tables 13 and 14 present results for baseline (SHHS1) and follow-up (SHHS2) subjects who had experienced at least one cardiovascular event before the corresponding PSG assessment. These individuals were generally older and exhibited higher incidence rates (in their case, recurrence) of cardiovascular events. For comparison, the mean (standard deviation) age in the primary SHHS1(E = 0, M = 0) cohort used for RSF training was 59.4 (11.2) years with a 12.6% incidence (=326/2579) of cardiovascular events, whereas SHHS1(E = 1, M = 0) subjects’ age was 68.6 (11.9) years with an incidence of 53.6%, and SHHS1(E = 1, M = 1) had subjects’ age of 70.7 (9.7) years with an incidence of 55.9%. The SHHS2 cohorts were, on average, even older. Despite these differences, predicted cardiovascular risk remained informative in these populations (with C-index and tdAUROC exceeding 50% and IBS below 50% in most cases). However, overall discriminatory ability was notably reduced. This decline in performance is not unexpected, as the RSF models were trained on a considerably younger, event-free population and are unlikely to generalize well to post-event subjects whose sleep patterns are likely to substantially differ^53–56^.

Finally, Supplementary Figures from Sections 5 and 6 depict the distribution of cardiovascular cases, survivors, and censoring, as well as the performance metrics (tdAUROC, IBS) for individual study subgroups concerning study wave (SHHS1, SHHS2), medications, prior cardiovascular events, and training vs. unseen subjects, for RFS with and without AHI, respectively.

#### Cardiovascular risk-profiles via partial effects of RSF

To examine associations between risk factors (demographics, BMI, smoking), sleep patterns (macrostructure, dynamics), and long-term cardiovascular outcomes, we analyzed partial effects from the RSF model, including AHI as a predictor. This adjustment accounts for SDB (AHI) and reduces potential bias. For comparison, we also assessed partial effects from the RSF model without AHI, which showed similar predictive performance.

Figure 4 shows the partial effects of demographic factors (age, gender), lifestyle variables (BMI, smoking status), and SDB severity (AHI) on the 10-year cardiovascular event-free probability, interpreted as the complement of risk. The event-free probability remains stable until ∼age 55, then declines, with a steep drop beyond age 80. BMI and AHI exhibit similar risk profiles due to their strong correlation. The highest event-free probability is observed for BMI values between 22–27; risk increases by ∼1% at lower BMI (possibly linked to smoking-related leanness) and up to 3% for BMI*>*35. A minimal cardiovascular risk is associated with an AHI of ≤15, supporting the clinical threshold that distinguishes mild from moderate SDB. Interestingly, near-zero AHI values are associated with a slight increase in risk, possibly due to poor sleep efficiency (low TST, high WASO), which can reduce AHI yet elevate cardiovascular risk. Partial effects also suggest a 1.5% higher risk in males and a ∼2% increase for current smokers versus never-smokers. Ex-smokers show a modest 0.5% higher risk than never-smokers but 1.5% lower risk than current smokers. These associations align with established evidence linking age, obesity, SDB, and smoking to elevated cardiovascular risk^1–3,9^.

**Figure 4.**
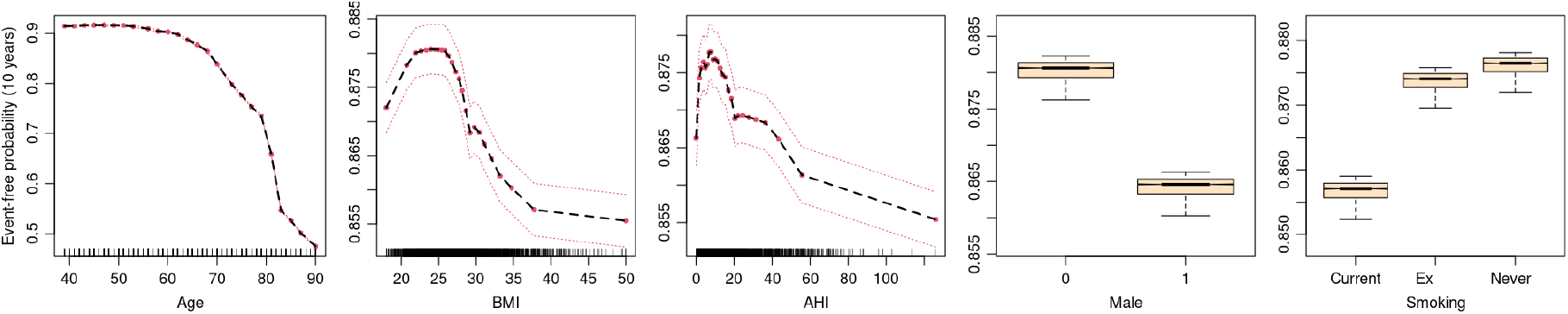
Partial effects and their 95% CIs for 10-year cardiovascular event-free probability for the age in years, Body Mass Index (BMI), Apnea-Hypopnea Index (AHI), gender (0 = female, 1 = male), and smoking status, for RSF with AHI predictor. Data points for continuous predictors are shown as ticks on the x-axis.

Figure 5 shows partial effects of sleep macro-architecture markers on 10-year event-free probability. Unlike Figure 2, where these predictors (except sleep latency, SL) exhibited monotonic, sigmoidal risk profiles for SDB, their associations with cardiovascular risk are predominantly U-shaped. This pattern suggests a “healthy optimum” range with maximal event-free probability, beyond which risk increases bidirectionally. Optimal TST appears around 360 minutes, with both short and long durations, reflecting insufficient sleep and hypersomnia, respectively, linked to up to a 3% increase in cardiovascular risk at the extremes. Short TST may arise from socially or psychiatrically induced sleep deprivation, renal dysfunction, or medical conditions. At the same time, long TST may result from central disorders of hypersomnolence, chronic medical conditions like metabolic dysfunction, systemic inflammation, neurodegeneration, or frailty^38–40^, all of which contribute to cardiovascular vulnerability. The lowest risk occurs at WASO of 50–80 minutes. Both very low and very high WASO are associated with elevated risk: low WASO (∼1.5% risk increase) may indicate sleep deprivation, frailty, advanced age, or neurodegenerative changes, while extremely high WASO (>200 minutes) suggests severe sleep fragmentation that may originate in insomnia, depression, chronic pain, or SDB. Sleep latency (SL) below 20 minutes or above 120 minutes corresponds to a ∼1.5% higher risk. Prolonged SL may reflect hyperarousal, insomnia, or anxiety, whereas very short SL could signal excessive sleepiness due to sleep deprivation (e.g., due to long-term SDB); however, SL can also be affected by discomfort or unfamiliarity with PSG recording. Very short REM latency (REML) is associated with a ∼3.5% risk increase, potentially reflecting sleep deprivation, narcolepsy, depression, or metabolic dysregulation. Prolonged REML (>200 minutes) shows a modest risk increase, possibly driven by SDB^32,36^. Similarly, both very short and prolonged deep-sleep latency (DL) are associated with higher risk (∼1.5% and ∼3.5%, respectively), likely reflecting chronic sleep deprivation, impaired sleep homeostasis, or fragmentation from SDB and depression^16,35^.

**Figure 5.**
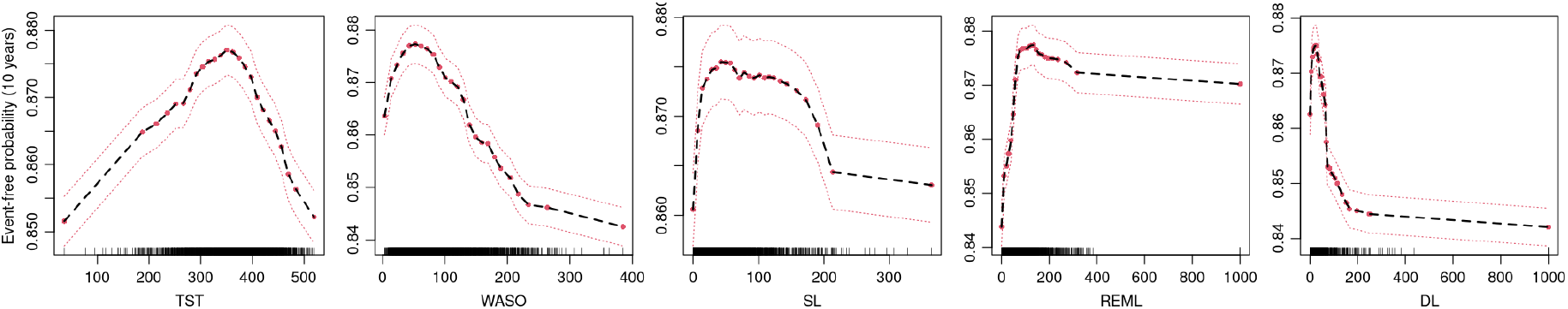
Partial effects and their 95% CIs for 10-year cardiovascular event-free probability for the minutes of Total Sleep Time (TST), Wake After Sleep Onset (WASO), Sleep Latency (SL), REM Latency (REM), and Deep-sleep Latency (DL), for RSF with AHI predictor. Data points are shown as ticks on the x-axis.

Figure 6 shows the partial effects of individual sleep-stage transition proportions *p*_*i, j*_ (cf. *Methods*), computed relative to the total number of sleep-stage epochs after sleep onset. On average, a post-onset PSG spans ∼910 epochs, so a 1% (= 0.01) change in transition proportion corresponds to roughly 9 transitions per PSG. We focus on clinically relevant transitions with partial effects linked to *≥*2% changes in predicted event-free probability, supported by non-overlapping 95% CIs.

1. *W-transitions*: The trend in *p*_*W,W*_ mirrors WASO, with continuous wakefulness beyond 50% linked to a 4% increase in cardiovascular risk. The optimal range for *p*_*W,N*1_ (a marker of fragmentation) is 0.01–0.03, with risk rising by 2% above 0.1. Direct transitions from W to N2, bypassing normal N1 sleep initiation, increase risk by 2% beyond 0.04. Notably, *p*_*W,N*3_ *>* 0.004 (*∼*3 transitions) may signal severe homeostatic sleep pressure and associate with up to a 6% risk increase.
2. *N1-transitions*: Frequent N1 →W transitions (*p*_*N*1,*W*_ *>* 0.02) reflecting heightened sleep instability, rise risk by >2%. Prolonged periods of light sleep (*p*_*N*1,*N*1_ *>* 0.15, ∼1 hour) show a linear association with ∼2% higher risk. Notably, even a single N1→N3 transition (*p*_*N*1,*N*3_ ∼ 0.001) links to >6% risk, suggesting abnormal homeostatic responses. Further, *p*_*N*1,*REM*_ *>* 0.01, reflecting atypical REM initiation, shows exponential risk increases, reaching 2% risk increase.
3. *N2-transitions*: Cardiovascular risk rises sharply (>7%) when *p*_*N*2,*N*1_ *>* 0.004 (*∼*3 transitions), indicating instability in intermediate NREM state. The lowest risk occurs with uninterrupted N2 sleep comprising 30–50% of post-onset time; deviations increase risk by >3%, particularly when *p*_*N*2,*N*2_ *>* 0.6, possibly reflecting reduced progression into N3 or REM sleep. Both insufficient (*p*_*N*2,*N*3_ *<* 0.005) and excessive (*p*_*N*2,*N*3_ *>* 0.06) deep sleep transitions are associated with an elevated risk, reflecting disturbed sleep pressure.
4. *N3-transitions*: Deep-sleep disruptions strongly predict cardiovascular risk. N3-awakenings, *p*_*N*3,*W*_ *>* 0.1 increase risk by *∼*2%. Atypical *p*_*N*3,*N*1_ *>* 0.001 transitions link to up to 10% higher risk per single occurrence. Minimal continuous N3 sleep (*p*_*N*3,*N*3_ ≈ 0) increases risk by 4%, while rare *p*_*N*3,*REM*_ *>* 0.001 associate with *∼*4% risk increase.
5. *REM-transitions*: REM awakenings, *p*_*REM,W*_, outside the [0.05, 0.15] range increase risk by up to 2%. The *p*_*REM,N*1_ > 0.01 transitions exhibit exponential risk growth (up to 3%), indicating heightened cortical arousals. Atypical *p*_*REM,N*3_ are linked to 8–12% higher risk for one or two nightly events, respectively. Optimal continuous REM sleep (*p*_*REM,REM*_) lies between [0.15, 0.25] corresponding to ∼1.5 hours of uninterupted REM over the night, while deviations are associated with >2% risk. The REM sleep disruptions likely reflect autonomic imbalance, mood-related dysregulation, or compensatory mechanisms.

**Figure 6.**
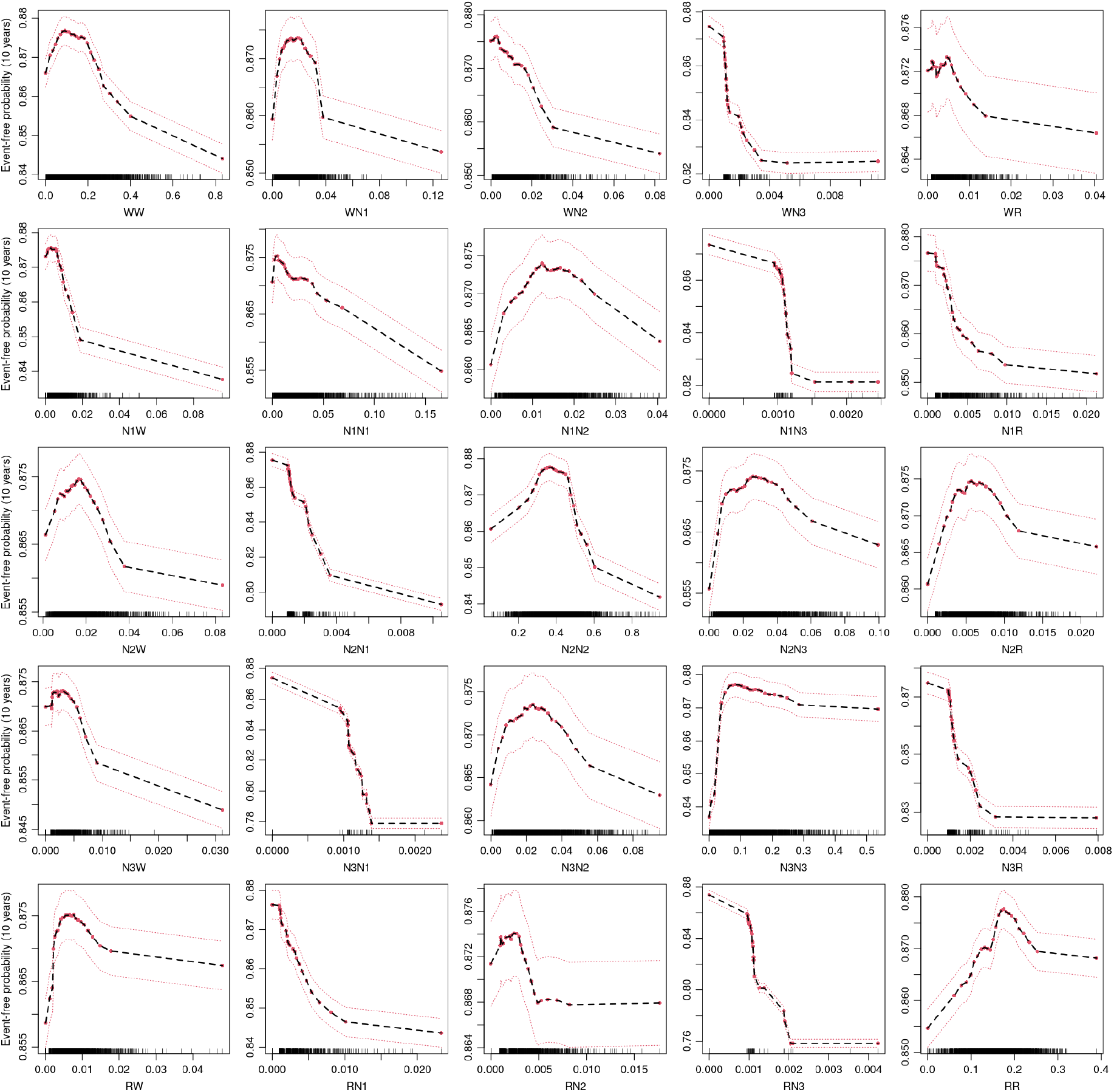
Partial effects and their 95% CIs for 10-year cardiovascular event-free probability for the relative frequencies of transitions between sleep-stage (W, N1, N2, N3, REM), for RSF with AHI predictor. Each subplot’s x-axis label indicates the direction of the transition (e.g., WN1 corresponds to transitions from W to N1). Data points are shown as ticks on the x-axis.

By comparing the partial effects from the RSF and RF models, we observe that while SDB risk is primarily associated with monotonic trends (e.g., lower TST, REM, N3, and higher WASO), cardiovascular risk often exhibits non-linear patterns. This likely reflects the influence of diverse clinical conditions beyond SDB that alter sleep parameters at both extremes. Finally, Supplementary Figures 25–27 display partial effects from the RSF model without the AHI predictor, showing trends consistent with those described above.

#### Correlation of predicted cardiovascular risk with sleep disorders and non-sleep comorbidities

The generalization tests above suggest that our RSF model accurately predicts cardiovascular risk, even under domain shifts involving older individuals and those on confounding medications. Leveraging the clinically rich BSWR data set, which spans a full spectrum of sleep disorders and selected non-sleep comorbidities, we next investigated how these conditions correlate with predicted cardiovascular risk. Each sleep disorder exhibits a distinct pattern of sleep disruption, which may differentially influence cardiovascular risk. Our goal was to determine whether specific sleep disorders and comorbidities are systematically associated with changes in predicted risk. Identifying such associations could provide insight into the cardiovascular relevance of individual sleep pathologies and help highlight high-risk patient subgroups.

Supplementary Table 2 summarizes the prevalence of major sleep-disorder classes and specific clinical conditions across the BSWR cohort and their associations with moderate-to-severe SDB (AHI *>* 15). Supplementary Table 3 provides further details on the demographic and clinical profiles (gender, age, BMI, and AHI) of each condition, comparing them to those of strictly healthy individuals. All seven major classes of sleep disorders significantly differed from healthy controls, showing higher mean age, BMI, and AHI, suggesting that individuals with sleep disorders tend to present with a riskier cardiovascular profile. While the healthy group comprised mostly women (58%), all sleep disorder categories—except insomnia and hypersomnia—had a significantly higher proportion of men (≥ 61.9%).

Across all major sleep disorder classes, we observed higher average predicted cardiovascular risk. However, this increase cannot be directly attributed to the disorders themselves, as individuals with sleep disorders also differed significantly from controls in demographics, BMI, and AHI. To address this, we quantified the adjusted risk using logistic regression models with log-odds-transformed predicted risk as the outcome, adjusting for age, gender, BMI, AHI, and a binary indicator for the specific condition. Each model was estimated in a case-control design contrasting healthy individuals with those affected by a given condition. The adjusted effect, reported in the final column of Supplementary Table 3, represents the systematic percentual increase in predicted cardiovascular risk relative to healthy controls, after accounting for differences in demographics, BMI, and AHI.

Even the adjusted models revealed significant risk increases across all major sleep disorder classes: SDB was associated with a 17.8%, 95% CI: (9.5, 26.8), increase in risk, insomnia 12.4% (2.9, 22.7), hypersomnia 11.0% (3.7, 18.9), movement-related disorders 20.3% (8.6, 33.2), parasomnias 24.5% (13.3, 36.9), circadian rhythm disorders 27.5% (10.3, 47.3), and isolated symptoms or normal variants 10.9% (2.9, 19.5). Specific conditions within these categories exhibited variability, as evidenced by a 39.2% (23.5, 56.9) increase for central sleep apnea, a 44.4% (27.0, 64.1) increase for NREM parasomnias, and non-significant effects for narcolepsy type 2, short-term insomnia, and idiopathic hypersomnia. Among comorbidities, the largest risk increase was observed for neurodegenerative diseases, 45.8% (21.4, 75.1); diabetes, 39.4% (13.9, 70.7); and current cardiac disease or prior event, 25.1% (8.7, 44.0).

These observed associations between sleep disorders, comorbidities, and elevated cardiovascular risk underscore the interrelationship between sleep and cardiovascular health. They also suggest that distinct patterns of sleep disruption, attributable to specific conditions, elevate the cardiovascular risk at different levels. However, further studies in cohorts with long-term monitored cardiovascular outcomes are needed to validate these observations, as such data are not currently available in BSWR.

## Discussion

A wide range of clinical conditions, including both sleep disorders and non-sleep comorbidities, can disrupt sleep macrostructure by altering total sleep duration, increasing fragmentation, and modifying the distribution of sleep stages^11–13,65–67^. In particular, SDB, affecting up to 23.4% and 49.7% middle to older-aged women and men^48^, respectively, induces characteristic macrostructural changes, including reductions in REM and N3 sleep, increased fragmentation, and altered stage composition^14–16,18^. SDB has also been linked to elevated cardiovascular morbidity and mortality, including hypertension, stroke, and sudden cardiac death^1–9^. Statistically, if a condition such as SDB alters both sleep macrostructure and the cardiovascular health, macrostructural sleep patterns may carry predictive information about cardiovascular risk, at least in an associative sense. Supporting this, reduced total sleep duration, as well as lower proportions of N3 and REM sleep, have been associated with increased cardiovascular risk^34–40^. Emerging evidence suggests that sleep-stage dynamics—the temporal patterns of transitions between stages—may offer deeper insights into physiological regulation and disease-specific signatures^17–24^. While prior studies focused on static macrostructural features, the prognostic relevance of dynamic sleep patterns for cardiovascular outcomes remains largely unexplored. Since alterations in sleep dynamics, like those in macrostructure, often arise as downstream effects of conditions such as SDB, diabetes, chronic pain, and neurodegenerative disorders, these patterns may encode signals relevant to cardiovascular risk.

Our study leveraged data from the SHHS, a prospective cohort originally designed to investigate the relationship between SDB and cardiovascular risk^42^. We used these data to examine the dependency chain linking SDB (a major cardiovascular risk factor), sleep characteristics (macrostructure, dynamics), common risk factors (demographics, BMI, smoking status), and long-term cardiovascular outcomes. Specifically, we assessed (i) whether SDB can be predicted from sleep parameters and common risk factors, and (ii) whether long-term cardiovascular risk can be predicted using the same features, both with and without explicit knowledge of SDB severity. To quantify these relationships, we applied forest-based methods^43,46^—Random Forest (RF) for SDB identification and Random Survival Forest (RSF) for cardiovascular risk prediction. These models are well-suited for capturing complex, non-linear relationships, robust to overfitting and multicollinearity, and support interpretability through partial dependence analyses. Notably, we analyzed R(S)F partial effects to determine whether individual predictors exhibited predominantly linear associations, as often assumed in prior studies using restrictive methods such as ANOVA^34^ or regression-based models^35–40^, or displayed non-linear patterns suggesting ranges of clinical optima with minimal risk. All models were trained on a carefully stratified SHHS baseline cohort, which was free from prior cardiovascular events and medication use, thereby minimizing confounding and enhancing the generalizability of our findings to a broader population.

The RF demonstrated that SDB can be reliably detected from the considered predictors. Cross-validation in the primary study cohort yielded an AUROC of 76.1% for identifying moderate-to-severe SDB (AHI *>*15), with strong generalization across REM- and NREM-dominant phenotypes (74.1-74.9%) and mixed SDB (79.4%). SDB detection remained robust even in unseen subgroups with prior events or medications from SHHS1 (73.6–74.1%), SHHS2 follow-up cohorts (69.5–80.6%), and in a fully out-of-domain clinical BSWR (76.0%). These findings demonstrate that SDB can be reliably inferred from sleep parameters and common risk factors only, even in medication or prior-event-confounded subgroups, and without direct access to respiratory signals typically required for clinical diagnosis. Partial dependence analyses revealed predominantly monotonic trends, with SDB risk increasing sharply above age 50, BMI *>*25, in males and ex-smokers, consistent with existing evidence^2,3,8,15,47–50^. Macrostructural sleep markers of SDB included TST *<*300 minutes, WASO *>*100 minutes, and prolonged REM and N3 latencies, confirming that apneic events cause fragmented and inefficient sleep, with delayed progression into restorative states^15,16,19^, known to be important for brain recreation. Novel insights emerged from sleep-stage transition proportions (*p*_*i*= from, *j*= to_) proposed in our previous work^20^, where *p*_*i, j*_ = 0.01 corresponds to roughly nine transitions per night while *p*_*i,i*_ = 0.1 indicates about 45 minutes of uninterrupted time in stage *i*. Several transitions proved to be highly sensitive markers, associated with >5% increases in SDB risk, including *p*_*W,N*2_ *>* 0.02, *p*_*W,REM*_ *>* 0.01, *p*_*N*1,*W*_ *>* 0.02, *p*_*N*2,*W*_ *>* 0.02, and reduced REM continuity (*p*_*REM,REM*_ *<* 0.1). While prior studies have quantified the effects of how the SDB alters sleep macrostructure and dynamics^17–20,22^, our findings suggest that these patterns alone enable effective screening of SDB, with partial effects providing mechanistic insight into these associations.

The RSF models further quantified the extent to which cardiovascular risk can be stratified from the same set of predictors. Two versions were trained: one that included SDB severity (AHI) and one that excluded it. Strikingly, inclusion of AHI did not improve performance on any discrimination or calibration metric assessing predictive capability to capture cardiovascular risk. For example, the cross-validation yielded C-indices of 73.0% and 73.3%, 10-year tdAUROCs of 75.1% and 75.3%, and IBS values of 6.7% for models with and without AHI, respectively, with significant log-rank tests having p-values of the same order. These findings suggest that in a flexible-enough model (RSF), demographic factors, BMI, smoking status, and sleep parameters sufficiently capture pathological signatures of cardiovascular risk, to the point that adding AHI offers no additional predictive benefit. This likely reflects the ability of these predictors to encode not only SDB-related patterns (as shown by the SDB-identification experiment) but also other pathological, possibly undiagnosed processes—such as diabetes^61^, renal dysfunction^62^, cancer^68^, pain syndromes^64^, or neurodegeneration^63^—that may jointly influence sleep and cardiovascular outcomes.

Supporting this, RSF partial effects of individual predictors revealed non-linear, often U-shaped risk profiles (in contrast to the monotonic effects in RF for SDB detection), suggesting clinical optima and thresholds of increased cardiovascular risk. Minimal risk was observed for age under 55 years, BMI ∈ [20, 25], AHI <15, and never-smokers. Macrostructural markers of minimal risk included TST ∈ [300, 400] minutes and WASO ∈ [40, 100] minutes, while deviations from these ranges—along with excessively short or long sleep-onset, REM, and N3 latencies—were associated with higher risk. Sleep-stage continuities in N2, N3, and REM stages exhibited protective ranges at *p*_*N*2,*N*2_ ∈ [0.3, 0.5], *p*_*N*3,*N*3_ ∈ [0.1, 0.3], and *p*_*REM,REM*_ ∈ [0.15, 0.25], corresponding to about [135, 225], [45,135], and [67.5, 112.5] minutes, respectively. These U-shaped risk profiles confirm prior associations between reduced TST, N3, and REM sleep durations with cardiovascular morbidity and mortality^35–40^, while extending them by showing that risk also increases above optimal values—a nuance not captured in earlier studies constrained to linear models. In addition, rare or highly atypical transitions, seldom observed in healthy sleep, were strongly associated with sharply monotonically increased risk. For instance, >3% risk increase was linked to *p*_*W,N*3_ *>* 0.002, *p*_*N*1,*W*_ *>* 0.03, *p*_*N*1,*N*3_ *>* 0.001, *p*_*N*2,*N*1_ *>* 0.003, *p*_*N*2,*N*2_ *>* 0.6, *p*_*N*3,*N*1_ *>* 0.001, *p*_*N*3,*N*3_ *<* 0.05, *p*_*N*3,*REM*_ *>* 0.002, *p*_*REM,N*1_ *>* 0.0075, *p*_*REM,N*3_ *>* 0.001, and absence of continuous REM sleep (*p*_*REM,REM*_ ≈ 0). Notably, even a single occurrence of such atypical transitions (e.g., *p*_*N*1,*N*3_, *p*_*N*3,*N*1_, *p*_*REM,N*3_) during a night may serve as a sensitive marker of cardiovascular risk, whether driven by SDB or other underlying conditions. Our findings extend the existing knowledge that sleep dynamics are not only useful for describing present clinical conditions, but also provide signals correlating with future health events.

Movel validation confirmed strong generalization of RSF predictions across SHHS subgroups with medication use (C-index >66.6%, IBS ≤12%, tdAUROC >69%, and significant log-rank test in all baseline or follow-up subgroups). However, performance was reduced in subjects with prior cardiovascular events, likely due to altered sleep-wake patterns caused by events (cf.^53–56^) and also much older age. In the BSWR data set, predicted cardiovascular risk was positively associated with all seven major sleep disorder classes (SDB, insomnia, hypersomnia, parasomnias, movement disorders, circadian-rhythm disorders, and isolated symptoms), with estimated adjusted increases in cardiovascular risk ranging from 10.9% to 27.5% compared to healthy controls. Among non-sleep comorbidities, neurodegenerative diseases, diabetes, and existing cardiac disease were associated with the highest increases (>25%). These findings collectively support the strong interplay between sleep and cardiovascular health.

## Conclusions

Our study demonstrates that sleep macrostructure and stage dynamics jointly encode sensitive markers of both current SDB and long-term cardiovascular risk. Leveraging carefully curated data from the large prospective SHHS cohort, we show that SDB can be reliably identified from sleep patterns and demographics alone, without the need for direct respiratory measurements. While we confirm established associations between short duration of TST, REM, and N3 with cardiovascular risk, the use of a flexible RSF modelling approach uncovered non-linear U-shaped relationships, revealing that excessive amounts of specific sleep features (including TST, REM) are also linked to increased risk—patterns overlooked by traditional linearly restricted methods. Notably, partial effects—providing insights into associations between risk and individual predictors—were largely monotonic for SDB, whereas cardiovascular risk exhibited predominantly U-shaped profiles, suggesting distinct physiological mechanisms and thresholds that could serve as novel markers in clinical decision-making. This suggests that cardiovascular vulnerability involves broader processes beyond SDB, reflected as downstream effects encoded in disrupted sleep. Hence, sleep architecture and dynamics act as a mirror of health, capturing signatures of current physiological states and predicting future disease risk. Together, they position sleep-stage patterns as promising, non-invasive biomarkers for diagnosing current conditions and stratifying long-term cardiovascular risk. With the rise of wearable technologies and automated sleep scoring, combined with additional biosignals such as respiratory patterns, oxygen saturation, and heart rate, these insights highlight the potential for large-scale, unobtrusive, and long-term monitoring, as well as future screening tools for cardiovascular health.

### Limitations

This study has several limitations. Despite using partial dependence analysis in the R(S)F framework, the quantified effects should be interpreted cautiously, as the modelling approach captures numerical associations and hence, partial effects should not be viewed as causal. In addition, as altered sleep patterns likely reflect downstream effects of different underlying conditions, the treatment interventions should target the root causes (e.g., SDB, diabetes management) rather than modifying sleep parameters in isolation. Next, our models were trained on participants free from prior cardiovascular events and medications, which, although improving generalizability, limits applicability in these subgroups. While forest-based methods can internally handle interactions between predictors, incorporating explicit age–gender interactions may be valuable, particularly given the protective effect of pre-menopause on cardiovascular outcomes. Additionally, we modelled a pooled composite cardiovascular endpoint, which may obscure specific risk patterns for individual outcomes. Future work could leverage competing risk models to disentangle and address each cardiovascular event separately. External validation of cardiovascular risk predictions in the BSWR data set was limited to adjusted associations with clinical conditions due to the lack of standardized time-to-event data. Therefore, in our future work, we plan to integrate causal knowledge on clinical relations, expand modelling to include key pharmacological classes and menopausal status, and harmonize data across multiple cohorts tracking long-term outcomes to enable robust cross-cohort validation.

## Materials and Methods

### Data sets

#### Sleep Heart Health Study (SHHS)

The SHHS is a multi-center, prospective cohort study designed to investigate the cardiovascular consequences of sleep-disordered breathing (SDB) in middle-aged and older adults^42^. The SHHS recruited participants from existing population-based cardiovascular and respiratory cohorts across the United States between 1995 and 1998. The baseline exam (SHHS1) was conducted on 6441 subjects and included in-home overnight polysomnography (PSG), comprehensive medical questionnaires, and cardiovascular assessments. A subset of 3295 participants underwent a second PSG study (SHHS2) about 5–8 years later as part of a follow-up evaluation. Participants have been longitudinally followed for over a decade to track major cardiovascular events and mortality. All PSG recordings in SHHS1 and SHHS2 were conducted without positive airway pressure (PAP) therapy, allowing participants to be considered untreated at the time of measurement. The SHHS data set comprises PSG biosignal data with sleep scoring annotations, detailed medication information, demographic and anthropometric measures, and details on the timing and type of cardiovascular outcomes. This study design enables the investigation of both cross-sectional and longitudinal relationships between sleep architecture and cardiovascular risk in a community-based population. In total, data from the 5839 participants who consented to share their information are available for research purposes.

##### Cardiovascular events

The outcome of interest was defined as the first occurrence of any major cardiovascular event recorded in SHHS and following the PSG assessment, including both clinical diagnoses and surgical interventions: angina, angioplasty, coronary artery bypass graft (CABG), congestive heart failure (CHF), myocardial infarction (MI), myocardial infarction procedure (MIP), percutaneous transluminal coronary angioplasty (PTCA), revascularization procedures, coronary stenting, or stroke. These events represent a mixture of atherosclerotic disease manifestations and interventional procedures commonly conducted in high-risk individuals. For survival analyses, we defined the event time as the number of days from the PSG recording (SHHS1 or SHHS2) to the first occurrence of any listed event. If no event occurred, the number of days to the most recent follow-up contact or recorded death since the PSG study was used as the censoring time. In addition, we identified whether individuals had experienced any of these cardiovascular events prior to the PSG study and used this information for stratification and assessments of the models’ generalizability.

##### Medication-related confounders

To account for potential pharmacological confounding, we created a binary indicator variable at both baseline (SHHS1) and follow-up (SHHS2) identifying subjects who were taking medications known or suspected to alter sleep-stage composition or cardiovascular risk. Based on clinical expertise, we flagged use of medications from several categories listed in SHHS metadata: psychiatric agents (e.g., tricyclic antidepressants, monoamine oxidase inhibitors, other antidepressants, antipsychotics, benzodiazepines), neurological agents (e.g., dopaminergic medications for Parkinson’s disease, cholinesterase inhibitors for Alzheimer’s disease), and selected cardiovascular drugs (e.g., beta-blockers, alpha-blockers, ACE inhibitors with diuretics, vasodilators, loop and thiazide diuretics). Aspirin was also included due to its high use and reported influence on slow-wave sleep. Medication status of subjects was extracted using SHHS drug codes, and the resulting indicator variable was used to support stratification and generalizability assessments across subgroups with and without pharmacological confounding.

##### Cohort stratification and notation

We analyzed a total of 8442 PSG recordings, comprising 5791 baseline recordings from SHHS1 and 2651 follow-up recordings from SHHS2. Each of these PSGs of unique individuals was successfully linked to available clinical and demographic metadata, including medications, cardiovascular event histories, event dates, and censoring information. To support subgroup analyses and generalizability tests, we organized the data according to three key attributes: (i) *study wave* (SHHS1 or SHHS2); (ii) *prior cardiovascular events* at the time of PSG recording (**E = 1** if any event occurred before PSG, **E = 0** otherwise); and (iii) *presence of medications* known to influence sleep or cardiovascular physiology (**M = 1** if such medications were reported, **M = 0** if not). The Supplementary Table 4 presents the number of subjects/PSGs (N) in each stratum, along with the number of subjects who developed an event following the PSG study, and the distribution of subject ages and genders. Our modelling efforts focused primarily on baseline subjects with no previous events or confounding medications, SHHS1(E = 0, M = 0), enabling the most precise evaluation of how specific sleep or demographic patterns associate with current SDB and the development of future cardiovascular events, not confounded by medication intake or prior events. The remaining subsets of data were used for validation and robustness assessments.

#### Bern Sleep-Wake Registery (BSWR)

To assess external validity and generalizability, we exploited the Bern Sleep-Wake Registry (BSWR) from Inselspital, University Hospital Bern. The BSWR contains over two decades of clinical polysomnography (PSG) data, starting from 2000. Most individuals in the BSWR suffer from one or more sleep disorders, with annotations including demographic information, clinical diagnoses, and relevant comorbidities beyond sleep-related conditions. For this study, we excluded daytime PSG recordings, studies shorter than 3 hours, instances where patients failed to fall asleep, and recordings involving positive airway pressure (PAP) therapy. Our final data set included 3702 PSG recordings from 3417 unique individuals aged 0–91 years (62.8% males), with a conclusive sleep diagnosis, complete demographic data (age, gender), and a calculated apnea–hypopnea index (AHI) to quantify the severity of sleep-disordered breathing (SDB). We primarily used the BSWR for external validation of moderate-to-severe SDB detection (AHI *>* 15). As BSWR currently lacks harmonized and matched time-to-event histories of cardiovascular outcomes, it was not feasible to use it for direct validation of long-term cardiovascular risk prediction. Instead, we leveraged the rich clinical annotations within the BSWR to evaluate associations between the model-predicted cardiovascular risk and specific sleep diagnoses, as well as relevant non-sleep comorbidities (e.g., prior stroke, diabetes). This strategy enabled both intuitive clinical validation and an indirect quantification of how individual clinical conditions relate to predicted cardiovascular risk. The Supplementary Table 2 presents the occurrence of different clinical conditions, including conclusive sleep disorders and non-sleep comorbidities, across BSWR subjects, and their stratification by sleep-disordered breathing status (AHI*≤*15 vs AHI>15).

### Data Preprocessing

All PSG recordings from both the SHHS and BSWR cohorts were scored into five standard sleep stages: Wake, N1, N2, N3, and REM, according to AASM guidelines^11^. Older recordings originally scored using the Rechtschaffen and Kales (R&K) guidelines were harmonized by merging the N3 and N4 stages into a single AASM-compliant N3 stage. The apnea–hypopnea index (AHI) was computed using the *recommended* AASM definition (v2.2, 2015) and used to derive binary sleep-disordered breathing (SDB) labels, with moderate-to-severe SDB defined as AHI > 15 and no-to-mild SDB defined as AHI ≤15. Gender was encoded as a binary male indicator (1 = male, 0 = otherwise); in all cases, the non-male category corresponded to participants self-identifying as female. Smoking status was encoded as a categorical variable with four levels: *current, ex, never*, or *not-available* (NA). Established sleep macrostructure features—such as total sleep time (TST), Wake After Sleep Onset (WASO), and stage-specific latencies—were computed directly from the hypnograms. Sleep dynamics were captured, following our prior work^20^, as a 5 × 5 matrix of sleep-stage transition proportions **P**, where each entry *p*_*i, j*_ denotes the percentage of all epochs, relative to the time after sleep onset, during which a transition from stage *i* to stage *j* occurred. For cardiovascular risk modelling, survival objects were constructed using the time from the PSG study (either baseline SHHS1 or the follow-up SHHS2) to the first joint cardiovascular event or censoring at the last contact.

#### Prediction, Validation, and Effect Quantification using Random (Survival) Forests

##### Modelling approach and predictors

To detect moderate-to-severe SDB (AHI∼*>*15), we employed a binary *Random Forest* (RF) classifier. For long-term cardiovascular risk prediction, we used *Random Survival Forests* (RSF), a non-parametric extension of RF for right-censored time-to-event data using the log-rank test as a splitting criterion^46^. Both RF and RSF are ensemble-based approaches, known for their robustness to overfitting, ability to capture non-linear relationships, and resilience to multicollinearity and high-dimensional settings (cf.^43–45^), making them particularly advantageous when using many correlated predictors such as the 25 sleep-stage transition proportions (**P**), where functional dependencies exist as all sum up to 100%. Each RF or RSF model was trained using the following predictor sets: *sleep dynamics* captured by 25 transition proportions, *conventional sleep metrics* not encoded in dynamics features: TST, WASO, and (sleep, N3, REM)-latencies, and *demographic or lifestyle variables:* age, gender, BMI, and smoking status. Additionally, for RSF, we experimented by adding AHI as an additional predictor to compare performance in cardiovascular risk prediction with and without the inclusion of direct SDB measurement. For both RF and RSF, we used default values of hyperparameters provided by the `randomForestSRC` R package (v3.1.1), which have been shown to perform reliably^52^.

**Validation** was carried out in three tiers. *First*, we used 5-fold cross-validation (CV) on the primary SHHS-baseline cohort of subjects without prior cardiovascular events or medication use, SHHS1(E = 0, M = 0), with approximately equal-sized folds created using fast anticlustering^69^ balancing the distribution of demographics and outcomes (age, gender, AHI, SDB- and cardiovascular-events prevalence). *Second*, models were tested on the remaining cohorts from SHHS1 and SHHS2 (cf. Supplementary Table 4), with additional control for subjects used during the models’ training. This assessed performance and discriminative power in the same subjects but several years later, i.e., by using SHHS2(E = 0, M = 0), and also in out-of-domain subjects with medications or prior events, which typically have higher rates of SDB prevalence, cardiovascular events, and different distributions of demographics (cf. Supplementary Tables 5-11). *Third*, external generalization was tested on a completely out-of-domain BSWR clinical data set, which, unlike SHHS, primarily contains a symptomatic clinical population suffering from multiple sleep disorders and non-sleep comorbidities. Selection of subjects with evaluated AHI enabled direct generalizability assessment of RF model in BSWR. The log-odds-transformed RSF-predicted cardiovascular risk in BSWR was regressed against different clinical conditions present in BSWR, while controlling for age, gender, BMI, and AHI, allowing for assessments of possible links between various conditions and an elevated cardiovascular risk.

**Performance metrics** for RF classification included the Area Under the Receiver Operating Characteristic curve (AUROC), Pearson’s correlation between the predicted probability of moderate-to-severe SDB and the observed AHI, as well as accuracy, sensitivity, specificity, and precision. For RSF, model performance was evaluated using Harrell’s concordance index (C-index), the Integrated Brier Score (IBS), and time-dependent AUROC (tdAUROC), each assessing the model’s ability to rank individuals according to their actual risk. Discriminative ability of RSF was further assessed via two-sample t-tests comparing the mean predicted *mortality* scores—interpreted as individual risk estimates scaled to event frequency^52^—between those who did and did not experience a future event. Lastly, log-rank tests were used to compare event incidence between high- and low-risk groups, stratified by the median predicted mortality.

##### Towards novel markers through partial effects

Beyond evaluating performance of R(S)F models, we assessed their *partial effects* quantifying the isolated contribution of each predictor to model output. Specifically, we used the `partial.rfsrc` function from the `randomForestSRC` package^52^, which quantifies changes in expected model prediction when one predictor is varied over its domain while all other features are held fixed and averaged over their empirical joint distribution. For RF, this yielded the partial effect of each variable on the probability of present moderate-to-severe SDB; for RSF, it revealed how each predictor influences the long-term (10 years) cardiovascular risk. These partial dependence functions provide interpretable insights into potential non-linearities, plateaus, or U-shaped effects, enabling models’ explanation and importantly, supporting clinical interpretation of the relationship between specific sleep (dynamics and macrostructure) parameters and cardiopulmonary outcomes. Based on this, partial effects may serve as future diagnostic or risk stratification markers.

## Author contributions

M.B. conceptualized the study, developed the methodology, performed the analysis, drafted the manuscript, and incorporated feedback from all co-authors. A.K. contributed to the study design and provided detailed feedback on related work, clinical interpretation, and the discussion. Y.T. contributed to the clinical stratification of the SHHS medication groups and provided detailed feedback. M.Sc. provided feedback on the methodology and contributed to manuscript refinements. J.v.d.M. assisted with BSWR data curation and provided detailed feedback. M.Sch., C.B., A.T., and F.F. read the manuscript and provided feedback. All authors approved the final manuscript and agreed to be listed as co-authors.

## Acknowledgements

This work was partly supported by JST FOREST grant JPMJFR2156 to A.K.

## Data availability

The data sets analyzed in this study are subject to different accessibility conditions: *Bern Sleep Wake Registery (BSWR):* Due to patient confidentiality and ethical restrictions, the BSWR data set is not publicly available. However, de-identified data may be obtained from the JvdM upon reasonable request, subject to a data sharing agreement and approval by the relevant ethics committees. *Sleep Heart Health Study (SHHS):* This data set is publicly accessible through the National Sleep Research Resource (NSRR) at https://sleepdata.org/datasets/shhs. Researchers can request access by creating an NSRR account and agreeing to the data use terms and conditions.

## Code availability

The underlying code supporting the findings of this study is publicly available at https://github.com/SleepAndCardiovascularRisk.

## Ethics declarations

This study used de-identified data from the Sleep Heart Health Study (SHHS), obtained from the National Sleep Research Resource (NSRR). The SHHS received ethical approval from institutional review boards at all participating U.S. centers, and all participants provided written informed consent. In accordance with the Swiss Human Research Act (HRA), no additional ethical approval was required for the secondary use of this anonymized, publicly available dataset.

The secondary usage of the Bern Sleep-Wake Registry (BSWR) was approved by the local ethics committee (Kantonale Ethikkommission Bern [KEK]-Nr. 2022-00415), ensuring compliance with the HRA and the Ordinance on Human Research with the Exception of Clinical Trials (HRO). All methods were carried out in accordance with relevant guidelines and regulations. Written informed consent was obtained from all participants included in this study, only de-identified data were made available, and these were maintained with confidentiality throughout the study.

