## Supplementary Materials for "Sleep-Stage Dynamics Predict Current Sleep-Disordered Breathing and Future Cardiovascular Risk"

### Supplementary Materials:
### *Sleep-Stage Dynamics Predicts Current Sleep Disordered Breathing and Future Cardiovascular Risk*

Bechny Michal et al. (2025)

#### 1 Bern Sleep-Wake Registery (BSWR)

##### Descriptive statistics

**Table 1.** Characteristics of Berner Sleep-Wake Registry (BSWR) cohort stratified no-to-mild SDB (AHI $\leq$ 15) versus moderate-to-high SDB (AHI > 15). Continuous variables are reported as mean (SD) and compared using Welch's two-sample *t*-test. Categorical variables, denoted by superscript*, are reported as counts (percentages) and compared using the chi-squared test. When expected cell counts were less than 5, Fisher's exact test was used instead.

| Variable | Overall | AHI $\leq$ 15 | AHI > 15 | p-value |
|---|---|---|---|---|
| N | 3702 | 2100 | 1602 | |
| Age | 48.28 (19.37) | 42.64 (19.53) | 55.66 (16.46) | <0.001 |
| Gender (Male)* | 2325 (62.8) | 1151 (54.8) | 1174 (73.3) | <0.001 |
| Smoking* | | | | 0.017 |
| Current | 179 (4.8) | 99 (4.7) | 80 (5.0) | |
| Ex | 67 (1.8) | 27 (1.3) | 40 (2.5) | |
| Never | 221 (6.0) | 115 (5.5) | 106 (6.6) | |
| NA | 3235 (87.4) | 1859 (88.5) | 1376 (85.9) | |
| BMI | 26.94 (6.46) | 25.52 (6.23) | 28.81 (6.29) | <0.001 |
| AHI | 19.02 (20.19) | 6.31 (4.23) | 35.69 (20.71) | <0.001 |
| SDB (AHI>15)* | 1602 (43.3) | 0 (0.0) | 1602 (100.0) | <0.001 |
| SDB category* | | | | <0.001 |
| Mixed | 951 (25.7) | 0 (0.0) | 951 (59.4) | |
| NREM-dominant | 484 (13.1) | 0 (0.0) | 484 (30.2) | |
| REM-dominant | 137 (3.7) | 0 (0.0) | 137 (8.6) | |
| AHI$\leq$15 | 2100 (56.7) | 2100 (100.0) | 0 (0.0) | |
| NA | 30 (0.8) | 0 (0.0) | 30 (1.9) | |
| TST [mins] | 339.13 (89.41) | 354.88 (91.52) | 318.49 (82.16) | <0.001 |
| WASO [mins] | 64.83 (54.42) | 56.47 (49.68) | 75.78 (58.30) | <0.001 |
| SE [%] | 80.03 (14.69) | 82.42 (13.58) | 76.89 (15.47) | <0.001 |
| SL [mins] | 18.64 (25.84) | 18.20 (24.83) | 19.22 (27.11) | 0.235 |
| REML [mins] | 172.40 (186.46) | 161.62 (169.84) | 186.52 (205.42) | <0.001 |
| DL [mins] | 74.92 (185.01) | 50.74 (137.39) | 106.61 (229.36) | <0.001 |
| W [%] | 16.45 (13.74) | 14.04 (12.54) | 19.60 (14.58) | <0.001 |
| N1 [%] | 16.87 (10.92) | 13.05 (7.77) | 21.89 (12.34) | <0.001 |
| N2 [%] | 36.40 (12.46) | 39.00 (11.46) | 33.00 (12.89) | <0.001 |
| N3 [%] | 16.93 (10.68) | 19.36 (10.70) | 13.73 (9.78) | <0.001 |
| REM [%] | 13.35 (7.05) | 14.55 (6.93) | 11.79 (6.90) | <0.001 |

##### Occurrence of clinical conditions

Table 2 shows the number and percentage, N (%), of PSG recordings in the BSWR, stratified by conclusive sleep diagnoses and non-sleep comorbidities and by the presence of sleep-disordered breathing (SDB): no-to-mild (AHI≤15) versus moderate-to-severe (AHI>15). Out of 3702 recordings from 3417 unique subjects, 2100 had AHI≤15 and 1602 had AHI>15. A total of 88 recordings corresponded to healthy individuals without any sleep diagnosis or on-sleep comorbidity, or undergoing PSG as healthy controls in some of the Inselspital's conducted clinical studies, without any clinical condition identified. The classification of major sleep disorder categories in the table is consistent with ICSD-3 (International Classification of Sleep Disorders, Third Edition), which defines seven major groups, such as SDB, Insomnias, and Central Disorders of Hypersomnolence (Hypersomnias). Within the most prevalent classes, we also considered the most relevant/common diagnoses subcategories (e.g., OSA and CSA within SDB, NT1 and NT2 within Hypersomnias) versus the less frequent "Other" conditions (e.g., hypoventilation and hypoxia syndrome in SDB). Rare categories (e.g., circadian rhythm disorders) and those with non-specific clinical profiles (e.g., isolated symptoms and normal variants) were grouped into a single main class.

We further grouped the available comorbidities into broader condition categories. The *Brain* category comprises individuals with a history of major neurological events, including stroke, intracerebral hemorrhage, and traumatic brain injury (TBI), as well as cases with suspected TBI-related diagnoses. This grouping captures subjects with structural brain injuries that may influence sleep physiology or contribute to comorbid conditions. The *Neurodegenerative* category comprises individuals with a confirmed or probable diagnosis of a neurodegenerative disorder. This includes Parkinson's disease, atypical Parkinsonian syndromes, amyotrophic lateral sclerosis (ALS), dementia, and other specified neurodegenerative conditions. The *Headache* category includes individuals with a confirmed or probable diagnosis of migraine, tension-type headache, post-traumatic headache, cluster headache, trigeminal neuralgia, and other specified headache syndromes. The *Psychiatric* category includes individuals with a confirmed or probable psychiatric disorder. This encompasses depression, bipolar disorder, anxiety and panic disorders, conversion disorder, post-traumatic stress disorder (PTSD), attention-deficit/hyperactivity disorder (ADHD), and substance-related disorders (alcohol and drug abuse), as well as other specified psychiatric conditions. The *Diabetes* category includes individuals with a confirmed or probable diagnosis of diabetes mellitus. The *Cardial* category includes probable or confirmed hypertension, coronary heart disease, atrial fibrillation, and other cardiac comorbidities. The *Pulmonary* category includes individuals with a confirmed or probable diagnosis of chronic respiratory disease, including chronic obstructive pulmonary disease (COPD), asthma, and other specified pulmonary conditions.

**Table 2.** Number and percentage (N, %) of different health conditions in Berner Sleep-Wake Registry (BSWR), stratified by the presence of no-to-mild sleep-disordered breathing (SDB; AHI $\leq$ 15) versus moderate-to-severe SDB (AHI $>$ 15). Equality of proportions between SDB groups was assessed using the chi-squared test or Fisher's exact test when expected cell counts were less than 5. Results are reported as p-values, with significant differences highlighted as follows: for $p < 0.05$, for $p < 0.01$, and for $p < 0.001$.

| Diagnosis | N (%) | AHI $\leq$ 15 | AHI>15 | p-value |
|---|---|---|---|---|
| **Total** | 3702 | 2100 | 1602 | |
| **Healthy** | 88 (2.4) | 88 (4.2) | 0 (0.0) | <0.001 |
| **Sleep Disorders:** | | | | |
| Sleep-disordered breathing (SDB) | 2695 (72.8) | 1120 (53.3) | 1575 (98.3) | <0.001 |
| Obstructive Sleep Apnea (OSA) | 687 (18.6) | 323 (15.4) | 364 (22.7) | <0.001 |
| Central Sleep Apnea (CSA) | 111 (3.0) | 33 (1.6) | 78 (4.9) | <0.001 |
| Other | 1957 (52.9) | 786 (37.4) | 1171 (73.1) | <0.001 |
| Insomnias | 487 (13.2) | 337 (16.0) | 150 (9.4) | <0.001 |
| Chronic | 105 (2.8) | 81 (3.9) | 24 (1.5) | <0.001 |
| Short-term | 2 (0.1) | 2 (0.1) | 0 (0.0) | 0.602 |
| Other | 385 (10.4) | 257 (12.2) | 128 (8.0) | <0.001 |
| Hypersomnias | 715 (19.3) | 584 (27.8) | 131 (8.2) | <0.001 |
| Narcolepsy Type 1 (NT1) | 58 (1.6) | 36 (1.7) | 22 (1.4) | 0.488 |
| Narcolepsy Type 2 (NT2) | 8 (0.2) | 7 (0.3) | 1 (0.1) | 0.161 |
| Idiopathic Hypersomnia (IH) | 24 (0.6) | 24 (1.1) | 0 (0.0) | <0.001 |
| Excessive Daytime Sleepiness (EDS) | 388 (10.5) | 332 (15.8) | 56 (3.5) | <0.001 |
| Other | 310 (8.4) | 251 (12.0) | 59 (3.7) | <0.001 |
| Movement-related | 265 (7.2) | 166 (7.9) | 99 (6.2) | 0.051 |
| Restless Leg Syndrome (RLS) | 182 (4.9) | 101 (4.8) | 81 (5.1) | 0.789 |
| Periodic Limb Movement Disorder (PLMD) | 34 (0.9) | 27 (1.3) | 7 (0.4) | 0.012 |
| Other | 50 (1.4) | 38 (1.8) | 12 (0.7) | 0.009 |
| Parasomnias | 193 (5.2) | 124 (5.9) | 69 (4.3) | 0.036 |
| REM | 55 (1.5) | 49 (2.3) | 6 (0.4) | <0.001 |
| NREM | 128 (3.5) | 68 (3.2) | 60 (3.7) | 0.456 |
| Other | 17 (0.5) | 10 (0.5) | 7 (0.4) | 1.000 |
| Circadian-rhythm-related | 47 (1.3) | 32 (1.5) | 15 (0.9) | 0.152 |
| Isolated symptoms and norm variants | 1771 (47.8) | 994 (47.3) | 777 (48.5) | 0.502 |
| **Non-sleep Comorbidities:** | | | | |
| Brain | 64 (1.7) | 34 (1.6) | 30 (1.9) | 0.646 |
| Neurodegenerative | 81 (2.2) | 47 (2.2) | 34 (2.1) | 0.900 |
| Epilepsy | 49 (1.3) | 35 (1.7) | 14 (0.9) | 0.052 |
| Headache | 73 (2.0) | 51 (2.4) | 22 (1.4) | 0.030 |
| Psychiatric | 204 (5.5) | 145 (6.9) | 59 (3.7) | <0.001 |
| Diabetes | 41 (1.1) | 22 (1.0) | 19 (1.2) | 0.810 |
| Cardial | 134 (3.6) | 54 (2.6) | 80 (5.0) | <0.001 |
| Pulmonary | 50 (1.4) | 35 (1.7) | 15 (0.9) | 0.078 |

#### Characteristics of clinical conditions, predicted risk, and their comparison to healthy.

**Table 3.** Summary statistics for Gender (N (%) of males) and mean (standard deviation) for Age, Body Mass Index (BMI), Apnea-Hypopnea Index (AHI), and predicted cardiovascular risk (mortality) in Bern Sleep-Wake Registry (BSWR). The adjusted risk, reported as an estimate (95% CI), quantifies the systematic percentual difference in predicted risk for specific diagnoses (conclusive sleep disorders and non-sleep comorbidities) using logistic regression adjusting for gender, age, BMI, and AHI. Significant differences in comparison to healthy controls are highlighted as: * if p-val<0.05, ** if p-val<0.01, and *** if p-val<0.001.

| Diagnosis | Gender-Male | Age | BMI | AHI | Predicted Risk | Adjusted Risk [%] |
|---|---|---|---|---|---|---|
| **Healthy** | 37 (42.0%) | 32.86 (18.81) | 22.63 (5.31) | 2.68 (1.85) | 17.14 (4.10) | NA |
| **Sleep Disorders:** | | | | | | |
| Sleep-disordered breathing (SDB) | 1868 (69.3%)*** | 51.1 (19.0)*** | 27.6 (6.5)*** | 24.6 (20.8)*** | 27.9 (9.9)*** | 17.8 (9.5, 26.8)*** |
| Obstructive Sleep Apnea (OSA) | 476 (69.3%)*** | 51.9 (17.1)*** | 27.8 (6.5)*** | 22.7 (19.8)*** | 27.1 (8.4)*** | 18.5 (9.4, 28.3)*** |
| Central Sleep Apnea (CSA) | 74 (66.7%)*** | 45.6 (24.7)*** | 25.0 (6.0)** | 29.6 (22.9)*** | 28.0 (9.5)*** | 39.2 (23.5, 56.9)*** |
| Other | 1359 (69.4%)*** | 51.1 (19.3)*** | 27.7 (6.6)*** | 24.9 (20.8)*** | 28.2 (10.3)*** | 17.4 (8.8, 26.7)*** |
| Insomnias | 262 (53.8%) | 53.6 (15.4)*** | 26.6 (5.3)*** | 13.8 (15.7)*** | 26.7 (9.5)*** | 12.4 (2.9, 22.7)** |
| Chronic | 61 (58.1%)* | 50.4 (15.8)*** | 26.2 (4.4)*** | 9.5 (9.4)*** | 24.4 (9.3)*** | 20.4 (5.6, 37.3)** |
| Short-term | 2 (100.0%) | 61.0 (5.7)* | 25.5 (3.5) | 5.4 (1.1) | 27.2 (3.7) | 41.4 (-10.5, 123.3) |
| Other | 202 (52.5%) | 54.5 (15.3)*** | 26.6 (5.5)*** | 15.0 (16.8)*** | 27.4 (9.8)*** | 14.7 (4.5, 25.9)** |
| Hypersomnias | 346 (48.4%) | 39.5 (15.5)** | 26.6 (6.4)*** | 9.3 (13.1)*** | 21.1 (6.9)*** | 11.0 (3.7, 18.9)** |
| Narcolepsy Type 1 (NT1) | 27 (46.6%) | 37.3 (16.7) | 27.6 (5.0)*** | 14.4 (16.1)*** | 23.1 (7.5)*** | 22.9 (8.8, 38.9)** |
| Narcolepsy Type 2 (NT2) | 6 (75.0%) | 29.8 (20.6) | 23.9 (5.0) | 7.4 (8.8) | 23.3 (8.9) | 12.7 (-11.8, 44.0) |
| Idiopathic Hypersomnia (IH) | 2 (8.3%)** | 26.3 (7.0)** | 24.3 (3.8) | 2.7 (2.0) | 17.2 (3.2) | 13.7 (-1.7, 31.4) |
| Excessive Daytime Sleepiness (EDS) | 162 (52.3%) | 39.3 (15.0)** | 26.8 (6.8)*** | 9.5 (13.0)*** | 20.7 (6.3)*** | 9.7 (1.4, 18.8)* |
| Other | 179 (46.1%) | 40.1 (15.4)** | 26.6 (6.7)*** | 8.2 (12.0)*** | 20.9 (7.1)*** | 10.8 (2.9, 19.2)** |
| Movement-related | 164 (61.9%)** | 53.2 (17.5)*** | 27.3 (6.4)*** | 16.5 (18.5)*** | 28.2 (11.1)*** | 20.3 (8.6, 33.2)*** |
| Restless Leg Syndrome | 103 (56.6%)* | 58.2 (15.1)*** | 27.7 (6.5)*** | 19.3 (20.3)*** | 30.0 (11.8)*** | 23.4 (8.5, 40.4)** |
| Periodic Limb Movement Disorder | 23 (67.6%)* | 51.1 (17.7)*** | 26.2 (4.7)*** | 9.7 (9.8)*** | 27.2 (9.2)*** | 32.6 (13.2, 55.3)*** |
| Other | 38 (76.0%)*** | 37.2 (15.6) | 26.8 (6.7)*** | 11.3 (13.3)*** | 22.6 (7.4)*** | 12.9 (-0.5, 28.2) |
| Parasomnias | 128 (66.3%)*** | 53.9 (19.1)*** | 25.8 (5.1)*** | 14.3 (14.5)*** | 27.3 (8.5)*** | 24.5 (13.3, 36.9)*** |
| REM | 36 (65.5%)* | 34.8 (16.1) | 24.3 (4.0)* | 6.9 (8.1)*** | 21.1 (5.3)*** | 16.1 (3.6, 30.2)* |
| NREM | 83 (64.8%)** | 62.9 (12.4)*** | 26.2 (5.2)*** | 17.8 (15.7)*** | 30.2 (7.8)*** | 44.4 (27.0, 64.1)*** |
| Other | 14 (82.4%)** | 50.6 (20.8)** | 26.3 (5.4)* | 11.4 (8.3)*** | 28.2 (11.7)** | 17.3 (-9.2, 51.5) |
| Circadian-rhythm-related | 33 (70.2%)** | 44.5 (17.1)*** | 27.6 (5.6)*** | 14.5 (15.2)*** | 26.8 (9.5)*** | 27.5 (10.3, 47.3)** |
| Isolated symptoms and norm variants | 1162 (65.6%)*** | 52.0 (17.7)*** | 27.4 (6.0)*** | 18.6 (18.9)*** | 27.4 (10.4)*** | 10.9 (2.9, 19.5)** |
| **Non-sleep Comorbidities:** | | | | | | |
| Brain | 47 (73.4%)*** | 53.6 (14.2)*** | 27.4 (4.6)*** | 21.5 (21.8)*** | 25.6 (9.1)*** | 10.2 (-5.5, 28.5) |
| Neurodegenerative | 45 (55.6%) | 63.9 (9.7)*** | 26.2 (4.8)*** | 18.0 (17.8)*** | 29.9 (9.3)*** | 45.8 (21.4, 75.1)*** |
| Epilepsy | 35 (71.4%)** | 44.7 (14.3)*** | 26.5 (5.7)*** | 11.4 (12.3)*** | 23.0 (6.3)*** | 14.5 (0.9, 30.0)* |
| Headache | 36 (49.3%) | 42.2 (15.5)*** | 27.7 (7.1)*** | 16.4 (21.8)*** | 22.7 (8.4)*** | 13.0 (0.6, 27.0)* |
| Psychiatric | 105 (51.5%) | 45.8 (16.2)*** | 28.1 (6.2)*** | 12.9 (15.4)*** | 23.6 (7.5)*** | 21.2 (10.1, 33.3)*** |
| Diabetes | 29 (70.7%)** | 55.9 (14.3)*** | 30.0 (8.7)*** | 25.7 (25.5)*** | 28.6 (9.3)*** | 39.4 (13.9, 70.7)** |
| Cardial | 108 (80.6%)*** | 57.2 (12.4)*** | 30.7 (6.3)*** | 27.8 (24.2)*** | 29.5 (9.0)*** | 25.1 (8.7, 44.0)** |
| Pulmonary | 33 (66.0%)* | 46.5 (16.7)*** | 29.3 (8.0)*** | 15.7 (20.1)*** | 23.8 (9.5)*** | 19.0 (2.3, 38.3)* |

#### 2 Sleep Heart Health Study (SHHS)

**Table 4.** Subsets of SHHS database stratified based on prior cardiovascular event status (E) and medication use (M). Summary statistics include the number of subjects (N), the number who developed a cardiovascular event during follow-up (N-events), the mean (SD) of age in years, and the number (%) of males.

| Dataset | N | N-events | Age | Gender |
|---|---|---|---|---|
| SHHS1 (E = 0, M = 0) | 2579 | 326 | 59.43 (11.16) | 1182 (45.8) |
| SHHS1 (E = 0, M = 1) | 2528 | 567 | 64.97 (10.15) | 1157 (45.8) |
| SHHS1 (E = 1, M = 0) | 112 | 60 | 68.61 (11.86) | 67 (59.8) |
| SHHS1 (E = 1, M = 1) | 572 | 320 | 70.67 (9.68) | 354 (61.9) |
| SHHS2 (E = 0, M = 0) | 811 | 62 | 63.16 (10.49) | 358 (44.1) |
| SHHS2 (E = 0, M = 1) | 1484 | 201 | 68.73 (9.60) | 647 (43.6) |
| SHHS2 (E = 1, M = 0) | 37 | 15 | 70.97 (10.55) | 22 (59.5) |
| SHHS2 (E = 1, M = 1) | 319 | 178 | 73.52 (9.06) | 199 (62.4) |

##### SHHS1

**Table 5.** Descriptive characteristics of SHHS1 (E = 0, M = 1) cohort stratified by cardiovascular event status. Continuous variables are reported as mean (SD) and compared using Welch's two-sample *t*-test. Categorical variables, denoted by superscript*, are reported as counts (percentages) and compared using the chi-squared test. When expected cell counts were less than 5, Fisher's exact test was used instead.

| Variable | Overall | Event-free | Event developed | p-value |
|---|---|---|---|---|
| N | 2528 | 1961 | 567 | |
| Age | 64.97 (10.15) | 63.32 (9.97) | 70.68 (8.57) | <0.001 |
| Gender (Male)* | 1157 (45.8) | 859 (43.8) | 298 (52.6) | <0.001 |
| Smoking* | | | | <0.001 |
| Current | 221 (8.7) | 166 (8.5) | 55 (9.7) | |
| Ex | 1121 (44.3) | 831 (42.4) | 290 (51.1) | |
| Never | 1174 (46.4) | 954 (48.6) | 220 (38.8) | |
| NA | 12 (0.5) | 10 (0.5) | 2 (0.4) | |
| BMI | 28.64 (5.22) | 28.60 (5.24) | 28.77 (5.14) | 0.507 |
| AHI | 18.89 (16.90) | 18.27 (16.85) | 21.05 (16.90) | 0.001 |
| SDB (AHI>15)* | 1179 (46.6) | 863 (44.0) | 316 (55.7) | <0.001 |
| SDB category* | | | | <0.001 |
| Mixed | 574 (22.7) | 425 (21.7) | 149 (26.3) | |
| NREM-dominant | 108 (4.3) | 78 (4.0) | 30 (5.3) | |
| REM-dominant | 379 (15.0) | 271 (13.8) | 108 (19.0) | |
| AHI≤15 | 1349 (53.4) | 1098 (56.0) | 251 (44.3) | |
| NA | 118 (4.7) | 89 (4.5) | 29 (5.1) | |
| TST [mins] | 358.28 (63.24) | 359.36 (63.05) | 354.54 (63.82) | 0.110 |
| WASO [mins] | 95.97 (55.62) | 93.97 (54.94) | 102.90 (57.42) | 0.001 |
| SE [%] | 70.89 (12.11) | 71.06 (12.11) | 70.28 (12.11) | 0.175 |
| SL [mins] | 52.19 (42.78) | 53.47 (43.32) | 47.75 (40.56) | 0.005 |
| REML [mins] | 122.90 (169.81) | 122.04 (167.02) | 125.89 (179.24) | 0.634 |
| DL [mins] | 83.39 (228.77) | 81.04 (225.39) | 91.53 (240.14) | 0.336 |
| W [%] | 20.98 (11.79) | 20.58 (11.67) | 22.38 (12.09) | 0.001 |
| N1 [%] | 4.16 (2.85) | 4.13 (2.81) | 4.25 (3.00) | 0.383 |
| N2 [%] | 45.87 (12.38) | 45.80 (12.44) | 46.09 (12.19) | 0.622 |
| N3 [%] | 13.95 (9.99) | 14.22 (9.98) | 13.00 (9.98) | 0.010 |
| REM [%] | 15.04 (6.27) | 15.26 (6.23) | 14.28 (6.35) | 0.001 |

**Table 6.** Descriptive characteristics of SHHS1 (E = 1, M = 0) cohort stratified by cardiovascular event status. Continuous variables are reported as mean (SD) and compared using Welch's two-sample *t*-test. Categorical variables, denoted by superscript*, are reported as counts (percentages) and compared using the chi-squared test. When expected cell counts were less than 5, Fisher's exact test was used instead.

| Variable | Overall | Event-free | Event developed | p-value |
|---|---|---|---|---|
| N | 112 | 52 | 60 | |
| Age | 68.61 (11.86) | 64.25 (13.48) | 72.38 (8.74) | <0.001 |
| Gender (Male)* | 67 (59.8) | 26 (50.0) | 41 (68.3) | 0.075 |
| Smoking* | | | | 0.401 |
| Current | 10 (8.9) | 5 (9.6) | 5 (8.3) | |
| Ex | 55 (49.1) | 22 (42.3) | 33 (55.0) | |
| Never | 47 (42.0) | 25 (48.1) | 22 (36.7) | |
| BMI | 27.80 (4.96) | 27.73 (6.13) | 27.86 (3.72) | 0.896 |
| AHI | 20.99 (15.27) | 18.97 (12.37) | 22.74 (17.31) | 0.194 |
| SDB (AHI>15)* | 67 (59.8) | 30 (57.7) | 37 (61.7) | 0.814 |
| SDB category* | | | | 0.524 |
| Mixed | 26 (23.2) | 9 (17.3) | 17 (28.3) | |
| NREM-dominant | 9 (8.0) | 4 (7.7) | 5 (8.3) | |
| REM-dominant | 24 (21.4) | 14 (26.9) | 10 (16.7) | |
| AHI≤15 | 45 (40.2) | 22 (42.3) | 23 (38.3) | |
| NA | 8 (7.1) | 3 (5.8) | 5 (8.3) | |
| TST [mins] | 349.95 (73.14) | 370.17 (57.88) | 332.42 (80.58) | 0.006 |
| WASO [mins] | 101.90 (64.26) | 81.72 (46.60) | 119.39 (72.25) | 0.002 |
| SE [%] | 69.66 (13.70) | 73.91 (10.40) | 65.98 (15.16) | 0.002 |
| SL [mins] | 50.80 (41.73) | 49.58 (35.41) | 51.86 (46.79) | 0.774 |
| REML [mins] | 124.81 (196.57) | 126.16 (185.06) | 123.63 (207.58) | 0.946 |
| DL [mins] | 81.23 (222.33) | 44.66 (139.63) | 112.92 (271.93) | 0.105 |
| W [%] | 22.47 (13.87) | 18.03 (10.05) | 26.32 (15.55) | 0.001 |
| N1 [%] | 4.72 (3.27) | 4.22 (2.96) | 5.15 (3.49) | 0.134 |
| N2 [%] | 45.44 (13.11) | 47.31 (12.40) | 43.82 (13.58) | 0.160 |
| N3 [%] | 12.33 (9.73) | 14.30 (11.16) | 10.63 (8.01) | 0.046 |
| REM [%] | 15.04 (6.39) | 16.14 (5.68) | 14.08 (6.84) | 0.089 |

**Table 7.** Descriptive characteristics of SHHS1 (E = 1, M = 1) cohort stratified by cardiovascular event status. Continuous variables are reported as mean (SD) and compared using Welch's two-sample *t*-test. Categorical variables, denoted by superscript*, are reported as counts (percentages) and compared using the chi-squared test. When expected cell counts were less than 5, Fisher's exact test was used instead.

| Variable | Overall | Event-free | Event developed | p-value |
|---|---|---|---|---|
| N | 572 | 252 | 320 | |
| Age | 70.67 (9.68) | 68.50 (10.28) | 72.38 (8.83) | <0.001 |
| Gender (Male)* | 354 (61.9) | 153 (60.7) | 201 (62.8) | 0.670 |
| Smoking* | | | | 0.557 |
| Current | 46 (8.0) | 21 (8.3) | 25 (7.8) | |
| Ex | 305 (53.3) | 128 (50.8) | 177 (55.3) | |
| Never | 221 (38.6) | 103 (40.9) | 118 (36.9) | |
| BMI | 28.08 (5.14) | 27.81 (5.01) | 28.29 (5.23) | 0.271 |
| AHI | 22.06 (17.36) | 20.23 (16.35) | 23.50 (18.02) | 0.025 |
| SDB (AHI>15)* | 323 (56.5) | 131 (52.0) | 192 (60.0) | 0.067 |
| SDB category* | | | | 0.296 |
| Mixed | 164 (28.7) | 62 (24.6) | 102 (31.9) | |
| NREM-dominant | 34 (5.9) | 14 (5.6) | 20 (6.2) | |
| REM-dominant | 82 (14.3) | 37 (14.7) | 45 (14.1) | |
| AHI≤15 | 249 (43.5) | 121 (48.0) | 128 (40.0) | |
| NA | 43 (7.5) | 18 (7.1) | 25 (7.8) | |
| TST [mins] | 347.37 (69.96) | 354.70 (66.21) | 341.59 (72.37) | 0.026 |
| WASO [mins] | 101.47 (58.91) | 96.46 (56.00) | 105.42 (60.91) | 0.071 |
| SE [%] | 68.79 (13.51) | 70.26 (13.25) | 67.63 (13.61) | 0.021 |
| SL [mins] | 57.04 (49.31) | 55.19 (47.87) | 58.49 (50.44) | 0.427 |
| REML [mins] | 130.81 (202.14) | 114.44 (170.03) | 143.70 (223.64) | 0.086 |
| DL [mins] | 117.64 (274.90) | 90.64 (232.66) | 138.89 (302.73) | 0.037 |
| W [%] | 22.58 (12.94) | 21.30 (12.07) | 23.59 (13.51) | 0.035 |
| N1 [%] | 4.38 (3.22) | 4.38 (2.90) | 4.38 (3.46) | 0.995 |
| N2 [%] | 46.09 (13.68) | 46.30 (12.71) | 45.92 (14.42) | 0.745 |
| N3 [%] | 12.58 (10.50) | 12.94 (9.68) | 12.30 (11.10) | 0.463 |
| REM [%] | 14.37 (6.43) | 15.08 (6.31) | 13.81 (6.47) | 0.019 |

#### SHHS2

**Table 8.** Descriptive characteristics of SHHS2 (E = 0, M = 0) cohort stratified by cardiovascular event status. Continuous variables are reported as mean (SD) and compared using Welch's two-sample *t*-test. Categorical variables, denoted by superscript*, are reported as counts (percentages) and compared using the chi-squared test. When expected cell counts were less than 5, Fisher's exact test was used instead.

| Variable | Overall | Event-free | Event developed | p-value |
|---|---|---|---|---|
| N | 811 | 749 | 62 | |
| Age | 63.16 (10.49) | 62.52 (10.16) | 70.89 (11.39) | <0.001 |
| Gender (Male)* | 358 (44.1) | 321 (42.9) | 37 (59.7) | 0.015 |
| Smoking* | | | | 0.010 |
| Current | 62 (7.6) | 51 (6.8) | 11 (17.7) | |
| Ex | 296 (36.5) | 272 (36.3) | 24 (38.7) | |
| Never | 445 (54.9) | 418 (55.8) | 27 (43.5) | |
| NA | 8 (1.0) | 8 (1.1) | 0 (0.0) | |
| BMI | 27.72 (4.69) | 27.70 (4.69) | 27.90 (4.73) | 0.749 |
| AHI | 15.84 (15.38) | 15.46 (15.16) | 20.35 (17.33) | 0.016 |
| SDB (AHI>15)* | 300 (37.0) | 268 (35.8) | 32 (51.6) | 0.019 |
| SDB category* | | | | 0.003 |
| Mixed | 158 (19.5) | 143 (19.1) | 15 (24.2) | |
| NREM-dominant | 20 (2.5) | 18 (2.4) | 2 (3.2) | |
| REM-dominant | 106 (13.1) | 96 (12.8) | 10 (16.1) | |
| AHI≤15 | 511 (63.0) | 481 (64.2) | 30 (48.4) | |
| NA | 16 (2.0) | 11 (1.5) | 5 (8.1) | |
| TST [mins] | 384.11 (60.69) | 385.78 (59.02) | 363.86 (75.76) | 0.006 |
| WASO [mins] | 155.78 (65.97) | 154.41 (65.70) | 172.35 (67.59) | 0.040 |
| SE [%] | 64.53 (10.36) | 64.74 (10.28) | 62.03 (11.10) | 0.047 |
| SL [mins] | 59.44 (39.19) | 60.15 (39.28) | 50.88 (37.36) | 0.073 |
| REML [mins] | 99.65 (107.53) | 97.15 (100.56) | 129.87 (168.91) | 0.021 |
| DL [mins] | 48.33 (132.58) | 47.30 (132.59) | 60.79 (132.83) | 0.442 |
| W [%] | 28.45 (10.67) | 28.16 (10.53) | 32.03 (11.75) | 0.006 |
| N1 [%] | 3.79 (3.76) | 3.72 (3.80) | 4.58 (3.09) | 0.084 |
| N2 [%] | 40.38 (9.28) | 40.35 (9.30) | 40.79 (9.05) | 0.715 |
| N3 [%] | 11.95 (7.86) | 12.18 (7.88) | 9.19 (7.19) | 0.004 |
| REM [%] | 15.43 (5.40) | 15.60 (5.39) | 13.41 (5.18) | 0.002 |

**Table 9.** Descriptive characteristics of SHHS2 (E = 0, M = 1) cohort stratified by cardiovascular event status. Continuous variables are reported as mean (SD) and compared using Welch's two-sample $t$-test. Categorical variables, denoted by superscript$^{*}$, are reported as counts (percentages) and compared using the chi-squared test. When expected cell counts were less than 5, Fisher's exact test was used instead.

| Variable | Overall | Event-free | Event developed | p-value |
|---|---|---|---|---|
| N | 1484 | 1283 | 201 | |
| Age | 68.73 (9.60) | 67.95 (9.51) | 73.72 (8.68) | <0.001 |
| Gender (Male)* | 647 (43.6) | 537 (41.9) | 110 (54.7) | 0.001 |
| Smoking* | | | | 0.082 |
| Current | 101 (6.8) | 80 (6.2) | 21 (10.4) | |
| Ex | 644 (43.4) | 552 (43.0) | 92 (45.8) | |
| Never | 719 (48.5) | 634 (49.4) | 85 (42.3) | |
| NA | 20 (1.3) | 17 (1.3) | 3 (1.5) | |
| BMI | 28.72 (5.30) | 28.79 (5.32) | 28.26 (5.19) | 0.190 |
| AHI | 18.60 (16.29) | 18.18 (16.17) | 21.31 (16.85) | 0.011 |
| SDB (AHI>15)* | 680 (45.8) | 569 (44.3) | 111 (55.2) | 0.005 |
| SDB category* | | | | 0.036 |
| Mixed | 376 (25.3) | 310 (24.2) | 66 (32.8) | |
| NREM-dominant | 52 (3.5) | 43 (3.4) | 9 (4.5) | |
| REM-dominant | 226 (15.2) | 195 (15.2) | 31 (15.4) | |
| AHI≤15 | 804 (54.2) | 714 (55.7) | 90 (44.8) | |
| NA | 26 (1.8) | 21 (1.6) | 5 (2.5) | |
| TST [mins] | 373.67 (71.17) | 376.93 (70.01) | 352.85 (75.07) | <0.001 |
| WASO [mins] | 166.48 (74.83) | 163.83 (73.64) | 183.40 (80.18) | 0.001 |
| SE [%] | 62.26 (11.95) | 62.72 (11.78) | 59.34 (12.59) | <0.001 |
| SL [mins] | 64.18 (46.50) | 64.55 (46.24) | 61.85 (48.15) | 0.445 |
| REML [mins] | 118.48 (131.83) | 114.62 (119.67) | 143.11 (190.70) | 0.004 |
| DL [mins] | 67.58 (182.98) | 62.35 (173.68) | 100.95 (231.55) | 0.005 |
| W [%] | 30.44 (12.33) | 29.91 (12.04) | 33.81 (13.60) | <0.001 |
| N1 [%] | 3.88 (3.05) | 3.79 (2.41) | 4.43 (5.61) | 0.006 |
| N2 [%] | 40.32 (10.44) | 40.51 (10.36) | 39.10 (10.89) | 0.077 |
| N3 [%] | 11.03 (8.09) | 11.20 (8.09) | 9.90 (8.02) | 0.033 |
| REM [%] | 14.34 (5.69) | 14.58 (5.60) | 12.75 (6.04) | <0.001 |

**Table 10.** Descriptive characteristics of SHHS2 (E = 1, M = 0) cohort stratified by cardiovascular event status. Continuous variables are reported as mean (SD) and compared using Welch's two-sample $t$-test. Categorical variables, denoted by superscript$^*$, are reported as counts (percentages) and compared using the chi-squared test. When expected cell counts were less than 5, Fisher's exact test was used instead.

| Variable | Overall | Event-free | Event developed | p-value |
|---|---|---|---|---|
| N | 37 | 22 | 15 | |
| Age | 70.97 (10.55) | 69.45 (12.55) | 73.20 (6.43) | 0.296 |
| Gender (Male)* | 22 (59.5) | 11 (50.0) | 11 (73.3) | 0.281 |
| Smoking* | | | | 0.242 |
| Current | 2 (5.4) | 2 (9.1) | 0 (0.0) | |
| Ex | 17 (45.9) | 8 (36.4) | 9 (60.0) | |
| Never | 18 (48.6) | 12 (54.5) | 6 (40.0) | |
| BMI | 27.74 (5.06) | 27.34 (5.73) | 28.34 (3.98) | 0.562 |
| AHI | 24.87 (16.70) | 24.80 (19.32) | 24.96 (12.56) | 0.979 |
| SDB (AHI>15)* | 27 (73.0) | 15 (68.2) | 12 (80.0) | 0.676 |
| SDB category* | | | | 0.689 |
| Mixed | 15 (40.5) | 7 (31.8) | 8 (53.3) | |
| NREM-dominant | 3 (8.1) | 2 (9.1) | 1 (6.7) | |
| REM-dominant | 8 (21.6) | 5 (22.7) | 3 (20.0) | |
| AHI≤15 | 10 (27.0) | 7 (31.8) | 3 (20.0) | |
| NA | 1 (2.7) | 1 (4.5) | 0 (0.0) | |
| TST [mins] | 362.89 (65.95) | 358.98 (75.93) | 368.63 (49.76) | 0.668 |
| WASO [mins] | 182.27 (74.72) | 188.80 (89.27) | 172.70 (47.33) | 0.528 |
| SE [%] | 59.95 (10.58) | 59.23 (12.05) | 61.00 (8.24) | 0.623 |
| SL [mins] | 65.80 (41.20) | 65.36 (42.79) | 66.43 (40.20) | 0.939 |
| REML [mins] | 78.95 (50.29) | 75.27 (50.41) | 84.33 (51.38) | 0.598 |
| DL [mins] | 45.00 (62.72) | 37.82 (37.15) | 55.53 (88.58) | 0.407 |
| W [%] | 32.88 (11.30) | 33.72 (13.45) | 31.66 (7.39) | 0.594 |
| N1 [%] | 3.76 (2.34) | 3.87 (2.67) | 3.60 (1.84) | 0.739 |
| N2 [%] | 39.74 (9.57) | 39.09 (10.67) | 40.70 (7.96) | 0.623 |
| N3 [%] | 10.38 (7.23) | 9.20 (6.12) | 12.12 (8.52) | 0.232 |
| REM [%] | 13.23 (5.87) | 14.13 (6.92) | 11.92 (3.69) | 0.266 |

**Table 11.** Descriptive characteristics of SHHS2 (E = 1, M = 1) cohort stratified by cardiovascular event status. Continuous variables are reported as mean (SD) and compared using Welch's two-sample *t*-test. Categorical variables, denoted by superscript*, are reported as counts (percentages) and compared using the chi-squared test. When expected cell counts were less than 5, Fisher's exact test was used instead.

| Variable | Overall | Event-free | Event developed | p-value |
|---|---|---|---|---|
| N | 319 | 141 | 178 | |
| Age | 73.52 (9.06) | 71.02 (9.87) | 75.50 (7.84) | <0.001 |
| Gender (Male)* | 199 (62.4) | 92 (65.2) | 107 (60.1) | 0.410 |
| Smoking* | | | | 0.471 |
| Current | 24 (7.5) | 13 (9.2) | 11 (6.2) | |
| Ex | 161 (50.5) | 73 (51.8) | 88 (49.4) | |
| Never | 129 (40.4) | 54 (38.3) | 75 (42.1) | |
| NA | 5 (1.6) | 1 (0.7) | 4 (2.2) | |
| BMI | 28.03 (4.54) | 28.00 (4.38) | 28.05 (4.67) | 0.922 |
| AHI | 23.56 (17.73) | 23.25 (17.40) | 23.80 (18.03) | 0.784 |
| SDB (AHI>15)* | 194 (60.8) | 90 (63.8) | 104 (58.4) | 0.386 |
| SDB category* | | | | 0.822 |
| Mixed | 122 (38.2) | 55 (39.0) | 67 (37.6) | |
| NREM-dominant | 14 (4.4) | 6 (4.3) | 8 (4.5) | |
| REM-dominant | 45 (14.1) | 23 (16.3) | 22 (12.4) | |
| AHI≤15 | 125 (39.2) | 51 (36.2) | 74 (41.6) | |
| NA | 13 (4.1) | 6 (4.3) | 7 (3.9) | |
| TST [mins] | 353.08 (76.90) | 354.40 (77.68) | 352.03 (76.48) | 0.785 |
| WASO [mins] | 182.43 (79.74) | 187.70 (86.63) | 178.26 (73.81) | 0.295 |
| SE [%] | 59.34 (12.32) | 59.10 (13.26) | 59.54 (11.56) | 0.752 |
| SL [mins] | 62.49 (42.57) | 62.05 (42.98) | 62.84 (42.35) | 0.869 |
| REML [mins] | 124.07 (164.14) | 113.00 (144.40) | 132.83 (178.15) | 0.285 |
| DL [mins] | 105.08 (248.30) | 102.35 (251.51) | 107.23 (246.42) | 0.862 |
| W [%] | 33.69 (13.32) | 34.11 (14.35) | 33.36 (12.47) | 0.618 |
| N1 [%] | 4.41 (3.13) | 4.17 (2.74) | 4.61 (3.40) | 0.215 |
| N2 [%] | 40.18 (11.10) | 39.96 (10.95) | 40.36 (11.25) | 0.750 |
| N3 [%] | 9.09 (7.72) | 8.90 (7.91) | 9.24 (7.58) | 0.696 |
| REM [%] | 12.62 (5.59) | 12.86 (5.70) | 12.43 (5.51) | 0.497 |

#### 3 Performance of Random Survival Forest model

**Table 12.** Performance of the Random Survival Forest (RSF) model omitting AHI predictor, evaluated across various datasets of subjects with no previous cardiovascular events (E = 0). The $^{CV}$ superscript denotes performance obtained via 5-fold cross-validation (CV) on the in-domain event- and medication-free (E = M = 0) baseline cohort SHHS1(E = 0, M = 0). All other columns evaluate the performance of the final RSF model fitted to the entire baseline cohort, applied to potentially out-of-domain subjects ($^{\dagger}$) from either the baseline (SHHS1) or follow-up (SHHS2) studies, including subgroups taking medication (M = 1). For each scenario, the number of subjects with events and without events (event-free) is reported. Model performance is assessed using Harrell's Concordance Index (C-index), Integrated Brier Score (IBS), and the time-dependent Area Under the Receiver Operating Characteristic curve (tdAUROC) at 1, 5, and 10 years. Discriminatory ability is evaluated via two-sided t-tests comparing predicted mortality between event and non-event subjects, including 95% confidence intervals (CI) for the difference (Diff.). Additionally, log-rank tests with Chi-squared ($\chi^2$) statistics compare event rates between high- and low-risk groups stratified by median predicted mortality. For the in-domain CV, mean (SD) of all metrics is reported.

| Metric | $SHHS1^{CV}$ (E = 0, M = 0) | $SHHS1^{\dagger}$ (E = 0, M = 1) | SHHS2 (E = 0, M = 0) | $SHHS2^{\dagger}$ (E = 0, M = 0) | SHHS2 (E = 0, M = 1) | $SHHS2^{\dagger}$ (E = 0, M = 1) |
|---|---|---|---|---|---|---|
| Events (N) | 65.2 (2.4) | 567 | 43 | 19 | 64 | 137 |
| Event-free (N) | 450.6 (2.7) | 1961 | 591 | 158 | 489 | 794 |
| C-index | 73.3 (2.5) | 69.7 | 74.1 | 78.3 | 70.1 | 66.6 |
| IBS | 6.7 (0.4) | 11.9 | 4.1 | 6.2 | 7.1 | 8.8 |
| 1-year tdAUROC | 77.1 (12.2) | 69.8 | 85.2 | 77.1 | 72 | 64.1 |
| 5-year tdAUROC | 74.9 (6.2) | 72.7 | 73.6 | 83.4 | 71.4 | 69.2 |
| 10-year tdAUROC | 75.3 (2.3) | 74.2 | - | 100 | - | 69 |
| Mortality (E = 1) | 21.1 (1.8) | 23.5 | 24.9 | 34.5 | 29.2 | 31.7 |
| Mortality (E = 0) | 11.3 (0.5) | 14.5 | 14.1 | 16 | 17.9 | 21.6 |
| Mortality Diff. | 9.8 (1.8) | 8.9 | 10.7 | 18.5 | 11.3 | 10.1 |
| Mortality Diff. CI-low | 5.8 (1.3) | 7.6 | 4.7 | 9.1 | 6.4 | 6.6 |
| Mortality Diff. CI-high | 13.7 (2.4) | 10.3 | 16.8 | 28 | 16.2 | 13.6 |
| p-value (t-test) | 0.000013 (0.000014) | $< 10^{-6}$ | 0.000856 | 0.000587 | 0.000019 | $< 10^{-6}$ |
| Events (N), high-risk | 50.8 (1.9) | 405 | 36 | 16 | 48 | 95 |
| Events (N), low-risk | 14.4 (1.9) | 162 | 7 | 3 | 16 | 42 |
| $\chi^2$ | 26 (4.7) | 162.5 | 22.1 | 10.4 | 20 | 28.4 |
| p-value (log-rank test) | 0.000005 (0.00001) | $< 10^{-6}$ | 0.000003 | 0.001237 | 0.000008 | $< 10^{-6}$ |

**Table 13.** Performance of the Random Survival Forest (RSF) model including AHI predictor, evaluated across various datasets of subjects with previous cardiovascular events (E = 1). The columns evaluate the performance of the final RSF model fitted to the entire event- and medication-free (E = M = 0) baseline cohort SHHS1(E = 0, M = 0), applied to potentially out-of-domain subjects (†) from either the baseline (SHHS1) or follow-up (SHHS2) studies, including subgroups taking medication (M = 1). For each scenario, the number of subjects with events and without events (event-free) is reported. Model performance is assessed using Harrell's Concordance Index (C-index), Integrated Brier Score (IBS), and the time-dependent Area Under the Receiver Operating Characteristic curve (tdAUROC) at 1, 5, and 10 years. Discriminatory ability is evaluated via two-sided t-tests comparing predicted mortality between event and non-event subjects, including 95% confidence intervals (CI) for the difference (Diff.). Additionally, log-rank tests with Chi-squared ($\chi^2$) statistics compare event rates between high- and low-risk groups stratified by median predicted mortality.

| Metric | SHHS1† (E = 1, M = 0) | SHHS1† (E = 1, M = 1) | SHHS2 (E = 1, M = 0) | SHHS2† (E = 1, M = 0) | SHHS2 (E = 1, M = 1) | SHHS2† (E = 1, M = 1) |
|---|---|---|---|---|---|---|
| Events (N) | 60 | 320 | 2 | 13 | 15 | 163 |
| Event-free (N) | 52 | 252 | 0 | 22 | 0 | 141 |
| C-index | 60.8 | 62.6 | 0.0 | 56.6 | 62.9 | 60.8 |
| IBS | 27.4 | 28.5 | 22 | 22.6 | 32.8 | 28.9 |
| 1-year tdAUROC | 29.2 | 64.9 | - | 78.1 | 100.0 | 62.5 |
| 5-year tdAUROC | 66.6 | 67.4 | - | 57.2 | 91.7 | 65.5 |
| 10-year tdAUROC | 73.2 | 69.3 | - | - | - | - |
| Mortality (E = 1) | 25.1 | 27.3 | 25.2 | 25 | 29.8 | 34.6 |
| Mortality (E = 0) | 18.1 | 21.0 | - | 25.4 | - | 27.1 |
| Mortality Diff. | 7.0 | 6.3 | - | -0.4 | - | 7.5 |
| Mortality Diff. CI-low | 1.2 | 3.6 | - | -11.5 | - | 3.1 |
| Mortality Diff. CI-high | 12.7 | 9.0 | - | 10.7 | - | 11.9 |
| p-value (t-test) | 0.017856 | 0.000005 | - | 0.94043 | - | 0.000846 |
| Events (N), high-risk | 36 | 184 | 1 | 7 | 7 | 95 |
| Events (N), low-risk | 24 | 136 | 1 | 6 | 8 | 68 |
| $\chi^2$ | 9.2 | 33.1 | 1.0 | 0.6 | 2.6 | 16.3 |
| p-value (log-rank test) | 0.002405 | $< 10^{-6}$ | 0.317311 | 0.442614 | 0.106251 | 0.000053 |

**Table 14.** Performance of the Random Survival Forest (RSF) model omitting AHI predictor, evaluated across various datasets of subjects with previous cardiovascular events (E = 1). The columns evaluate the performance of the final RSF model fitted to the entire event- and medication-free (E = M = 0) baseline cohort SHHS1(E = 0, M = 0), applied to potentially out-of-domain subjects (†) from either the baseline (SHHS1) or follow-up (SHHS2) studies, including subgroups taking medication (M = 1). For each scenario, the number of subjects with events and without events (event-free) is reported. Model performance is assessed using Harrell's Concordance Index (C-index), Integrated Brier Score (IBS), and the time-dependent Area Under the Receiver Operating Characteristic curve (tdAUROC) at 1, 5, and 10 years. Discriminatory ability is evaluated via two-sided t-tests comparing predicted mortality between event and non-event subjects, including 95% confidence intervals (CI) for the difference (Diff.). Additionally, log-rank tests with Chi-squared ($\chi^2$) statistics compare event rates between high- and low-risk groups stratified by median predicted mortality.

| Metric | SHHS1† (E = 1, M = 0) | SHHS1† (E = 1, M = 1) | SHHS2 (E = 1, M = 0) | SHHS2† (E = 1, M = 0) | SHHS2 (E = 1, M = 1) | SHHS2† (E = 1, M = 1) |
|---|---|---|---|---|---|---|
| Events (N) | 60 | 320 | 2 | 13 | 15 | 163 |
| Event-free (N) | 52 | 252 | 0 | 22 | 0 | 141 |
| C-index | 61.7 | 62.2 | 0 | 56.6 | 61.9 | 61.1 |
| IBS | 27.4 | 28.5 | 21.8 | 22.4 | 32.8 | 28.8 |
| 1-year tdAUROC | 30.1 | 64.4 | - | 79.2 | 100 | 63 |
| 5-year tdAUROC | 67.8 | 67 | - | 57.4 | 91.7 | 66 |
| 10-year tdAUROC | 73.7 | 69.1 | - | - | - | - |
| Mortality (E = 1) | 25.0 | 27.3 | 25.1 | 25.5 | 30.1 | 34.6 |
| Mortality (E = 0) | 18.0 | 21.0 | - | 25.5 | - | 26.9 |
| Mortality Diff. | 7.0 | 6.3 | - | 0 | - | 7.7 |
| Mortality Diff. CI-low | 1.1 | 3.6 | - | -11.3 | - | 3.3 |
| Mortality Diff. CI-high | 12.8 | 9.0 | - | 11.3 | - | 12.1 |
| p-value (t-test) | 0.019666 | 0.000006 | - | 0.997352 | - | 0.000583 |
| Events (N), high-risk | 36 | 181 | 1 | 7 | 7 | 95 |
| Events (N), low-risk | 24 | 139 | 1 | 6 | 8 | 68 |
| $\chi^2$ | 9.2 | 30.4 | 1.0 | 0.6 | 1.1 | 16.1 |
| p-value (log-rank test) | 0.002405 | $< 10^{-6}$ | 0.317311 | 0.442614 | 0.286982 | 0.000061 |

### 4 Survival Plots for RSF with AHI predictor

#### 4.1 Primary study cohort

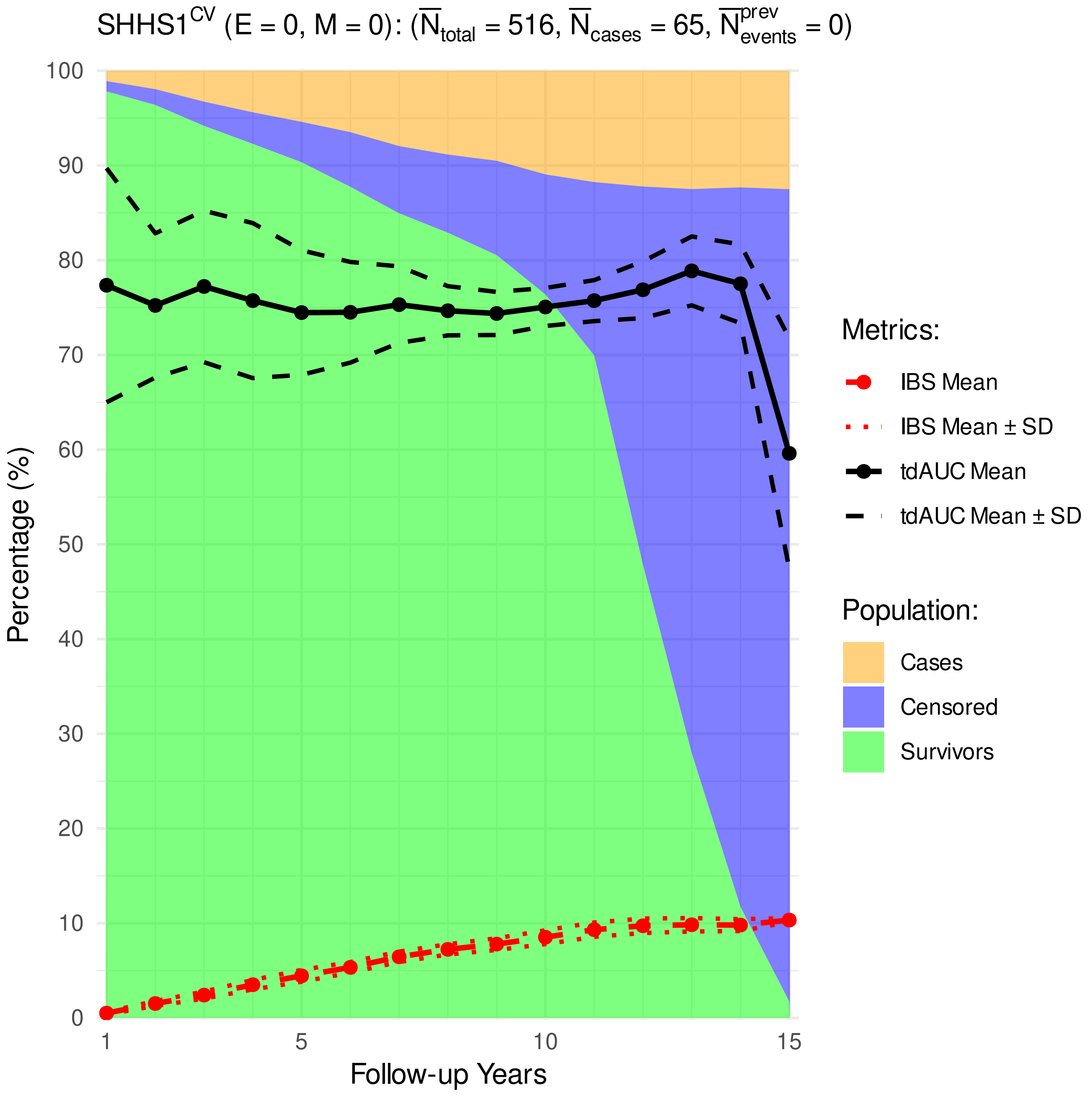


**Figure 1.** Distribution of cardiovascular cases, survivors, and censoring, as well as the performance metrics (time-dependent Area Under Receiver Operating Characteristic [tdAUROC] curve, Integrated Brier Score [IBS].)

##### 4.2 SHHS1 test subjects

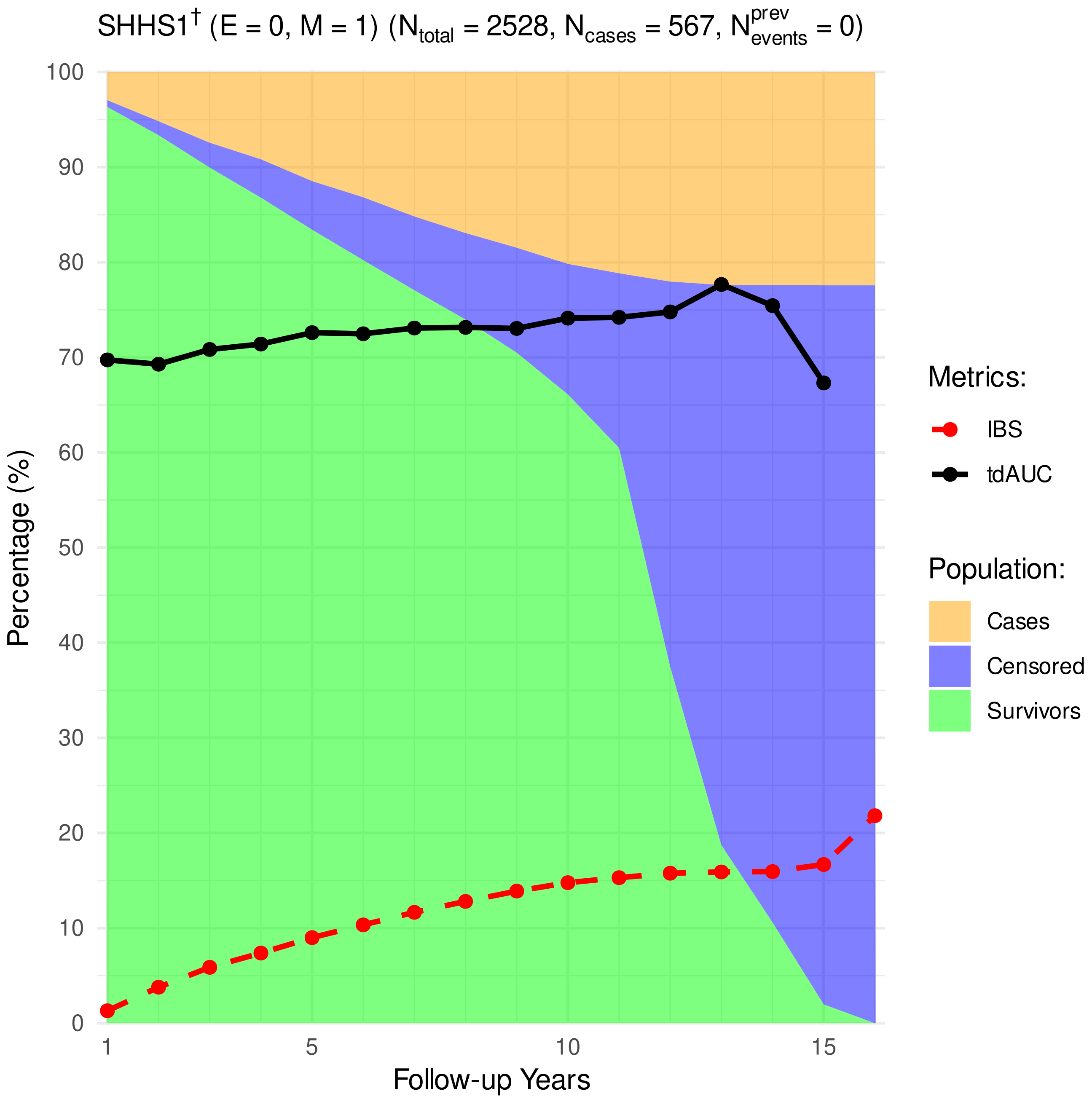


**Figure 2.** Distribution of cardiovascular cases, survivors, and censoring, as well as the performance metrics (time-dependent Area Under Receiver Operating Characteristic [tdAUROC] curve, Integrated Brier Score [IBS].)

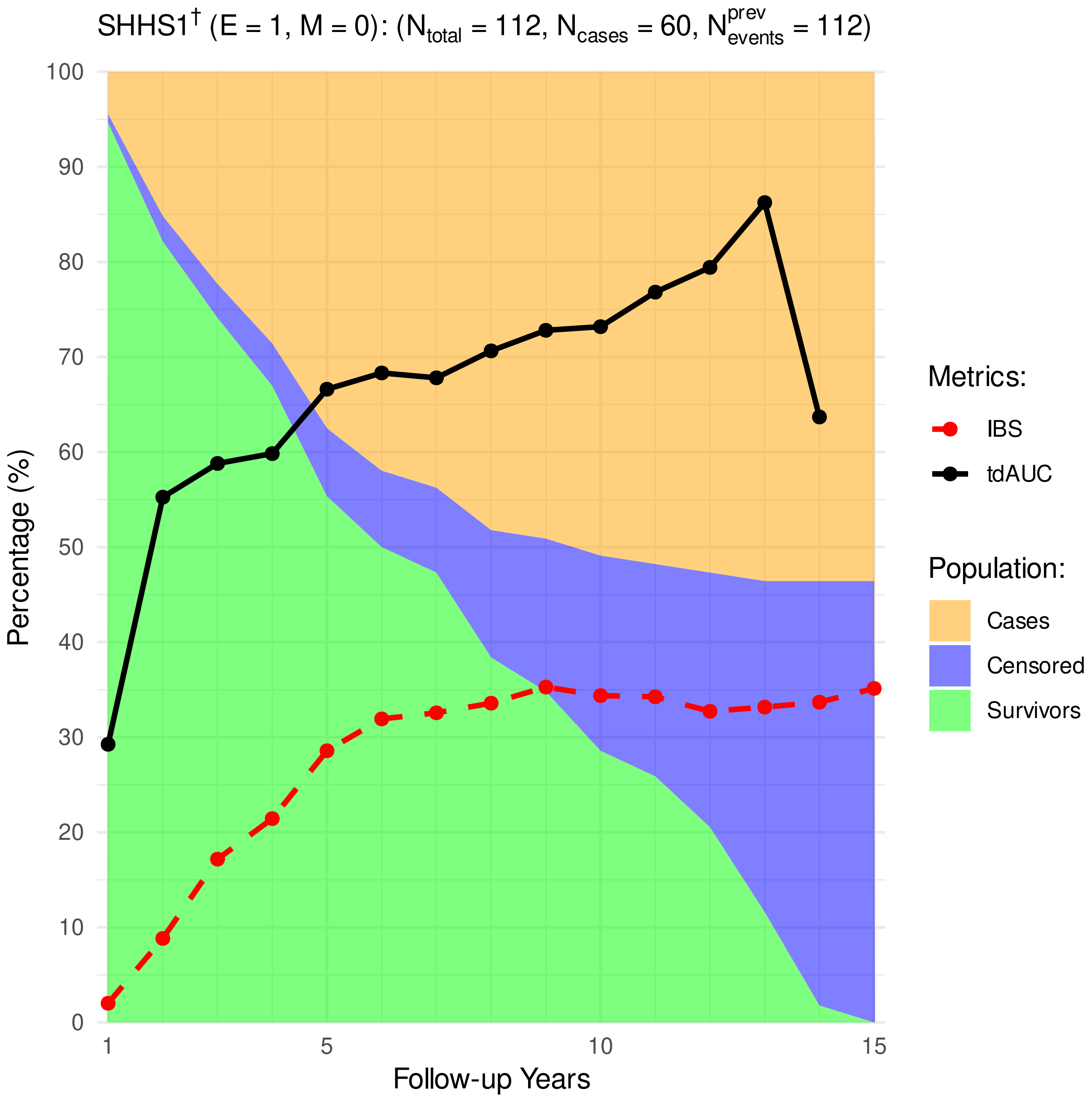


**Figure 3.** Distribution of cardiovascular cases, survivors, and censoring, as well as the performance metrics (time-dependent Area Under Receiver Operating Characteristic [tdAUROC] curve, Integrated Brier Score [IBS].)

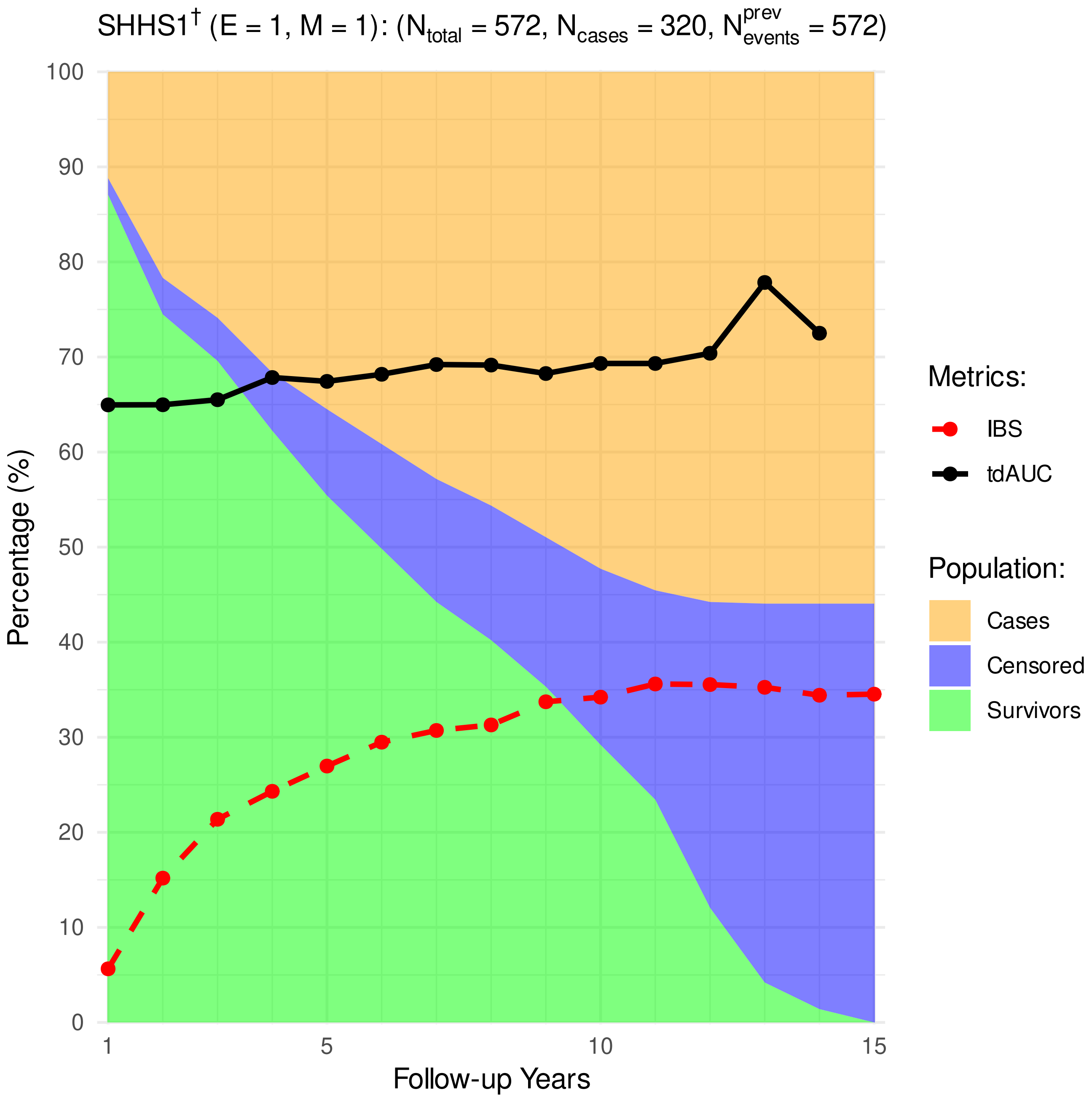


**Figure 4.** Distribution of cardiovascular cases, survivors, and censoring, as well as the performance metrics (time-dependent Area Under Receiver Operating Characteristic [tdAUROC] curve, Integrated Brier Score [IBS].)

##### 4.3 SHHS2 train subjects

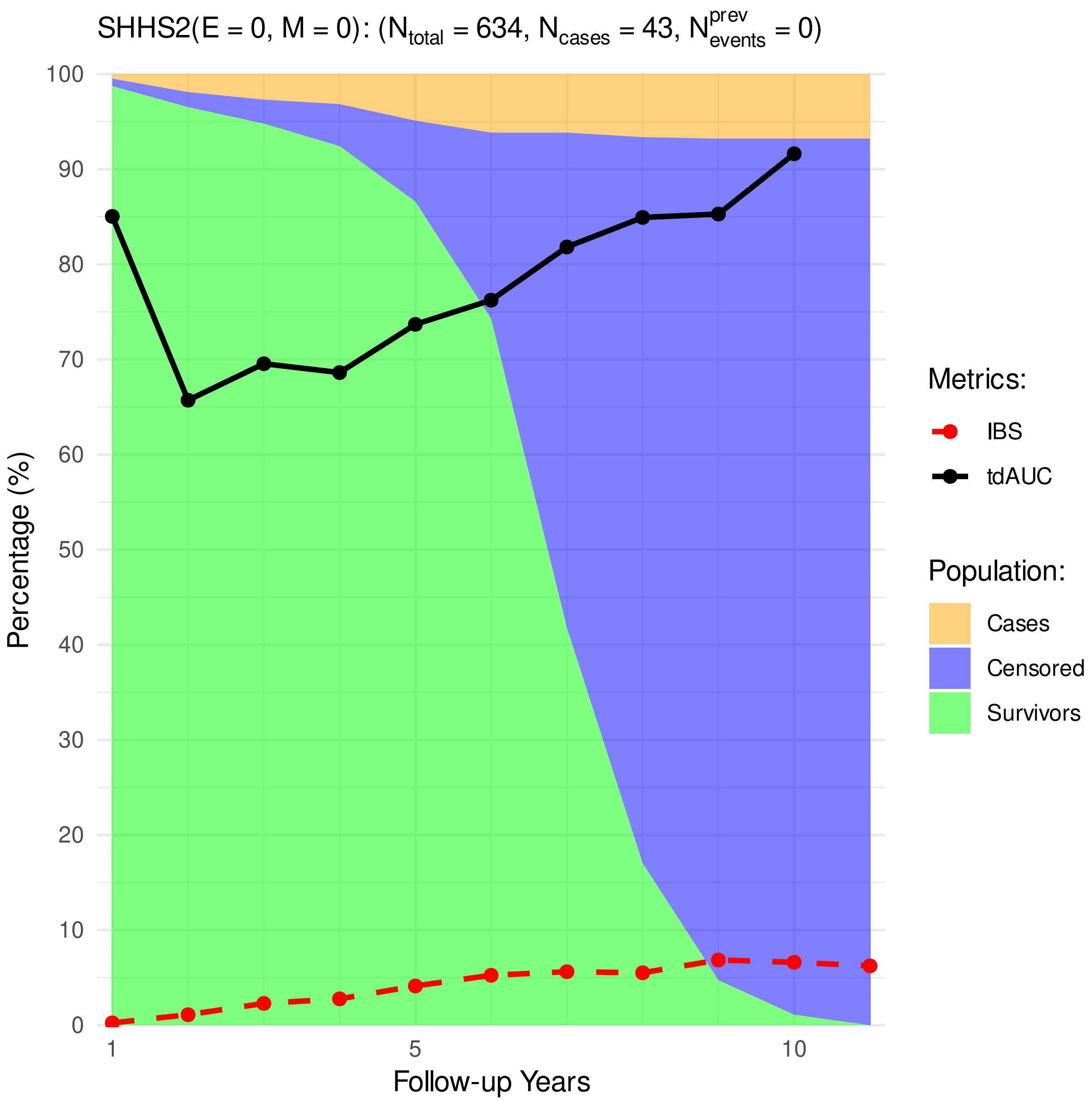


**Figure 5.** Distribution of cardiovascular cases, survivors, and censoring, as well as the performance metrics (time-dependent Area Under Receiver Operating Characteristic [tdAUROC] curve, Integrated Brier Score [IBS].)

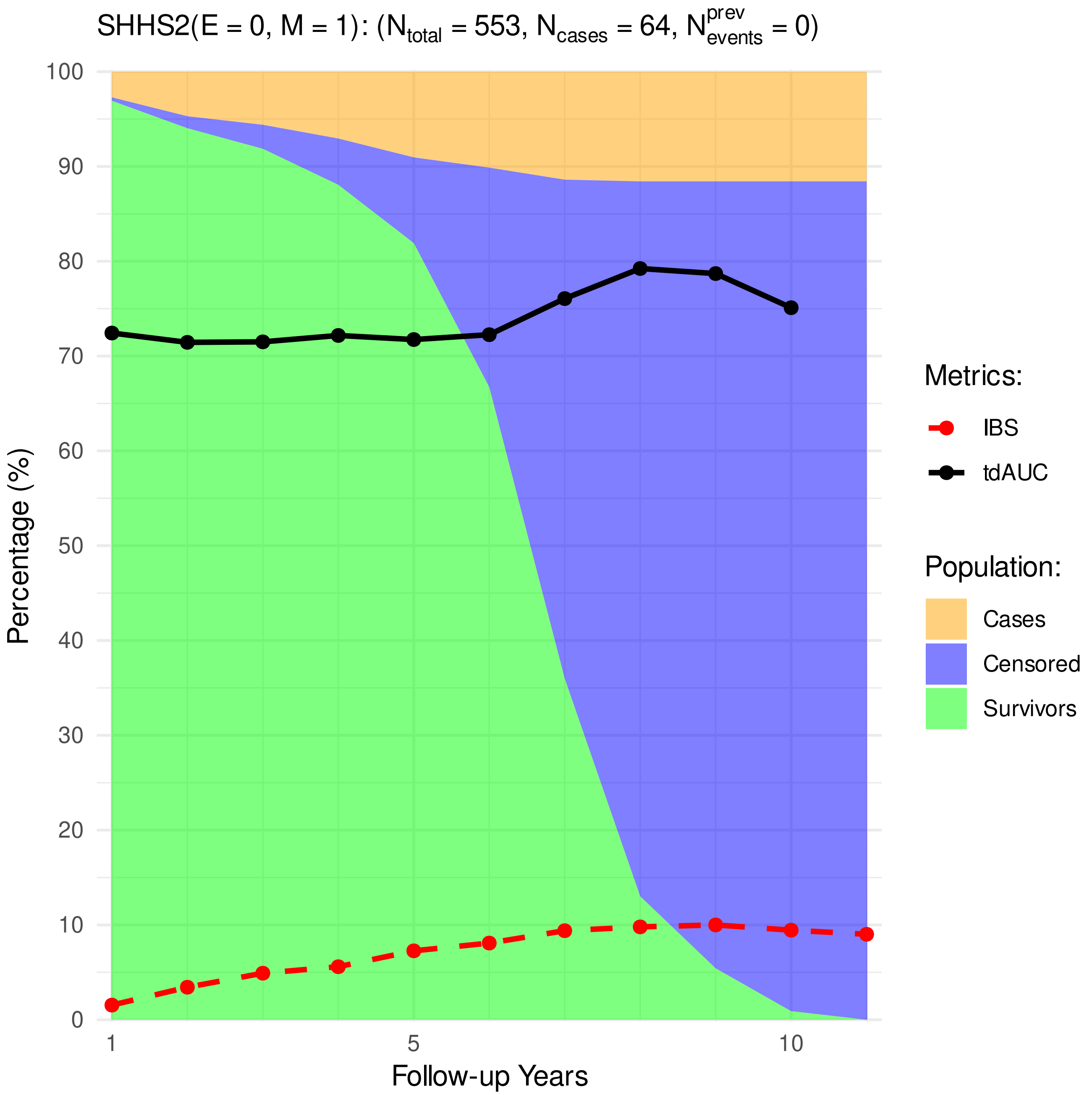


**Figure 6.** Distribution of cardiovascular cases, survivors, and censoring, as well as the performance metrics (time-dependent Area Under Receiver Operating Characteristic [tdAUROC] curve, Integrated Brier Score [IBS].)

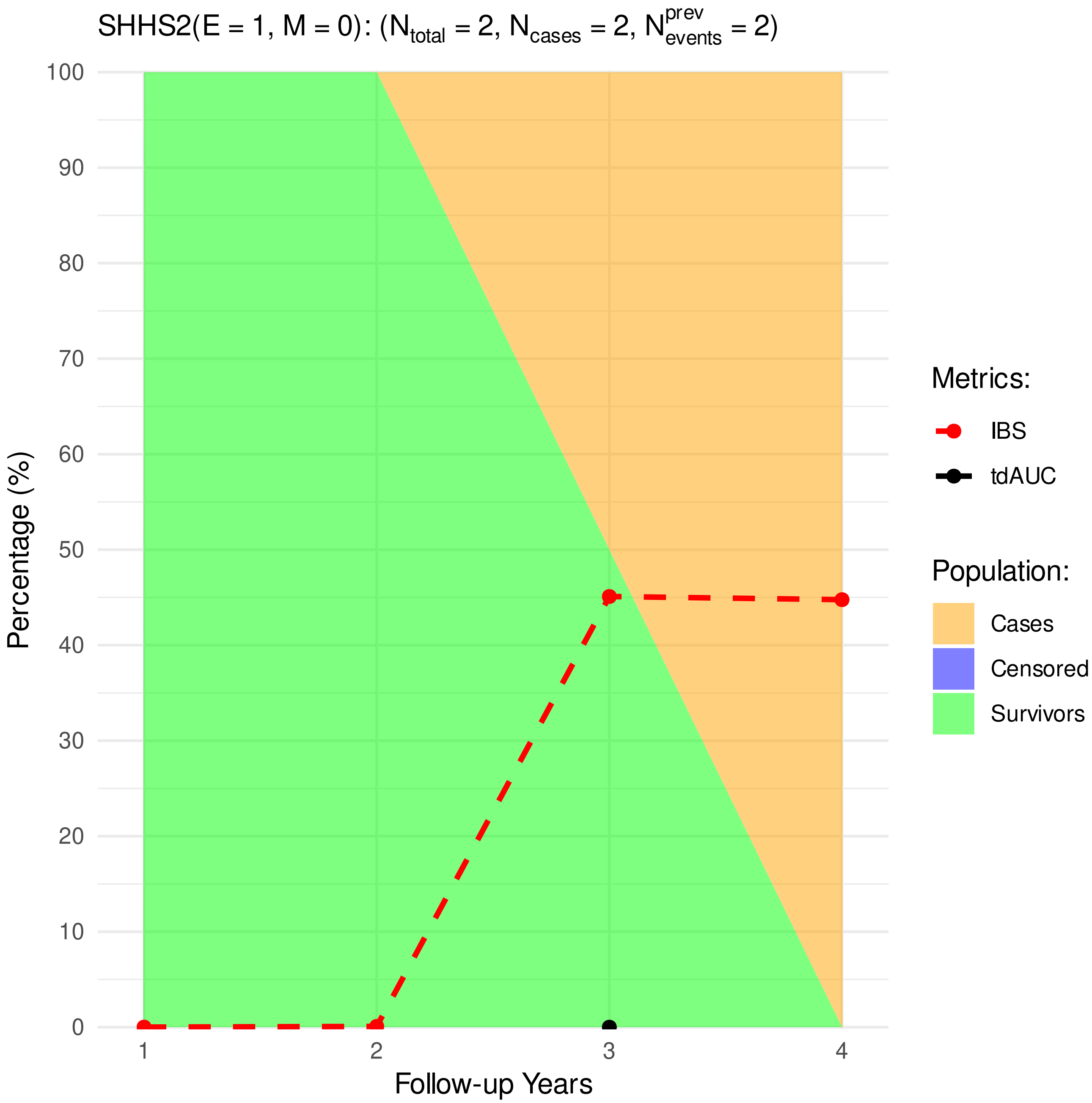


**Figure 7.** Distribution of cardiovascular cases, survivors, and censoring, as well as the performance metrics (time-dependent Area Under Receiver Operating Characteristic [tdAUROC] curve, Integrated Brier Score [IBS].)

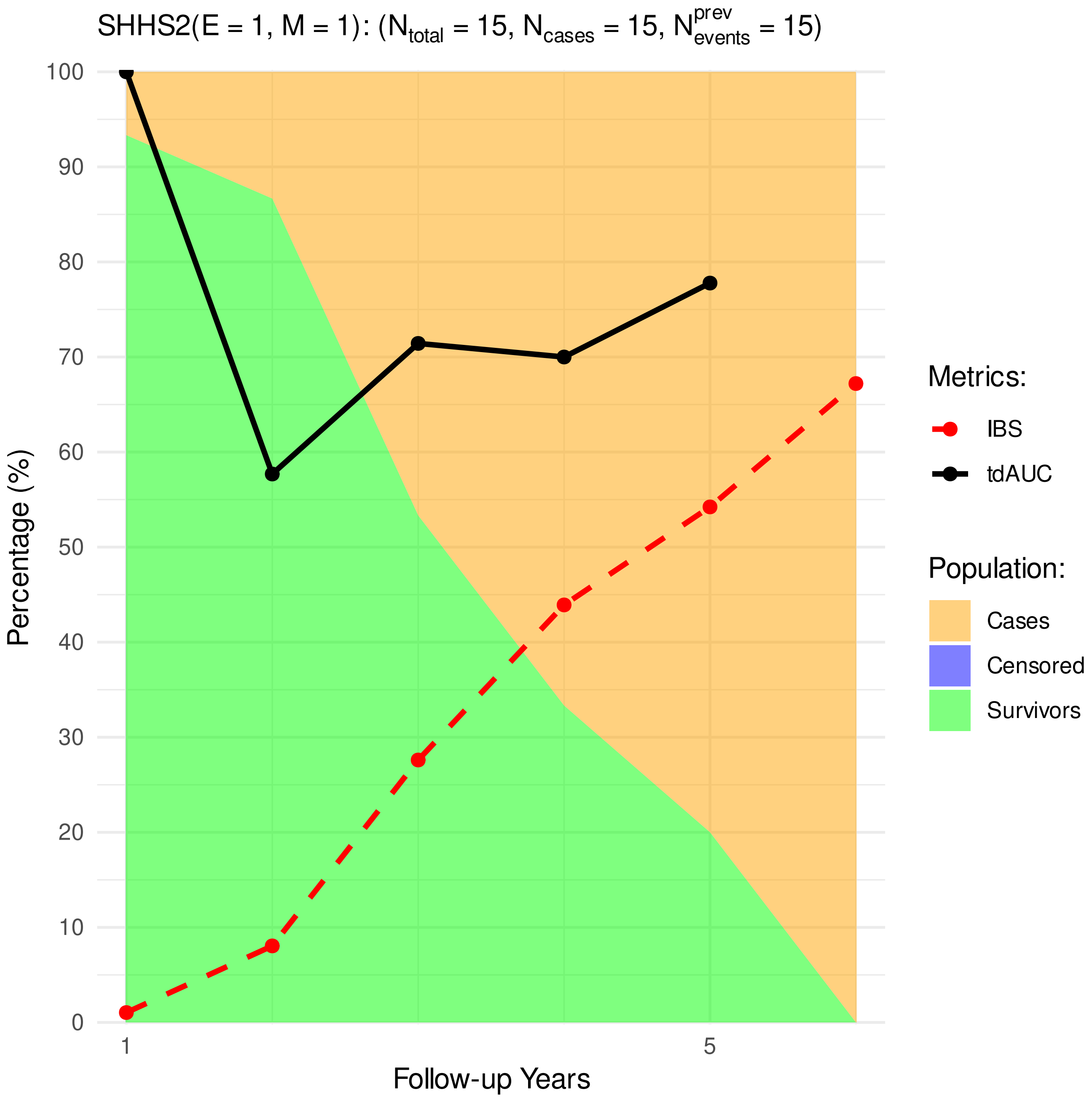


**Figure 8.** Distribution of cardiovascular cases, survivors, and censoring, as well as the performance metrics (time-dependent Area Under Receiver Operating Characteristic [tdAUROC] curve, Integrated Brier Score [IBS].)

#### 5 SHHS2 test subjects

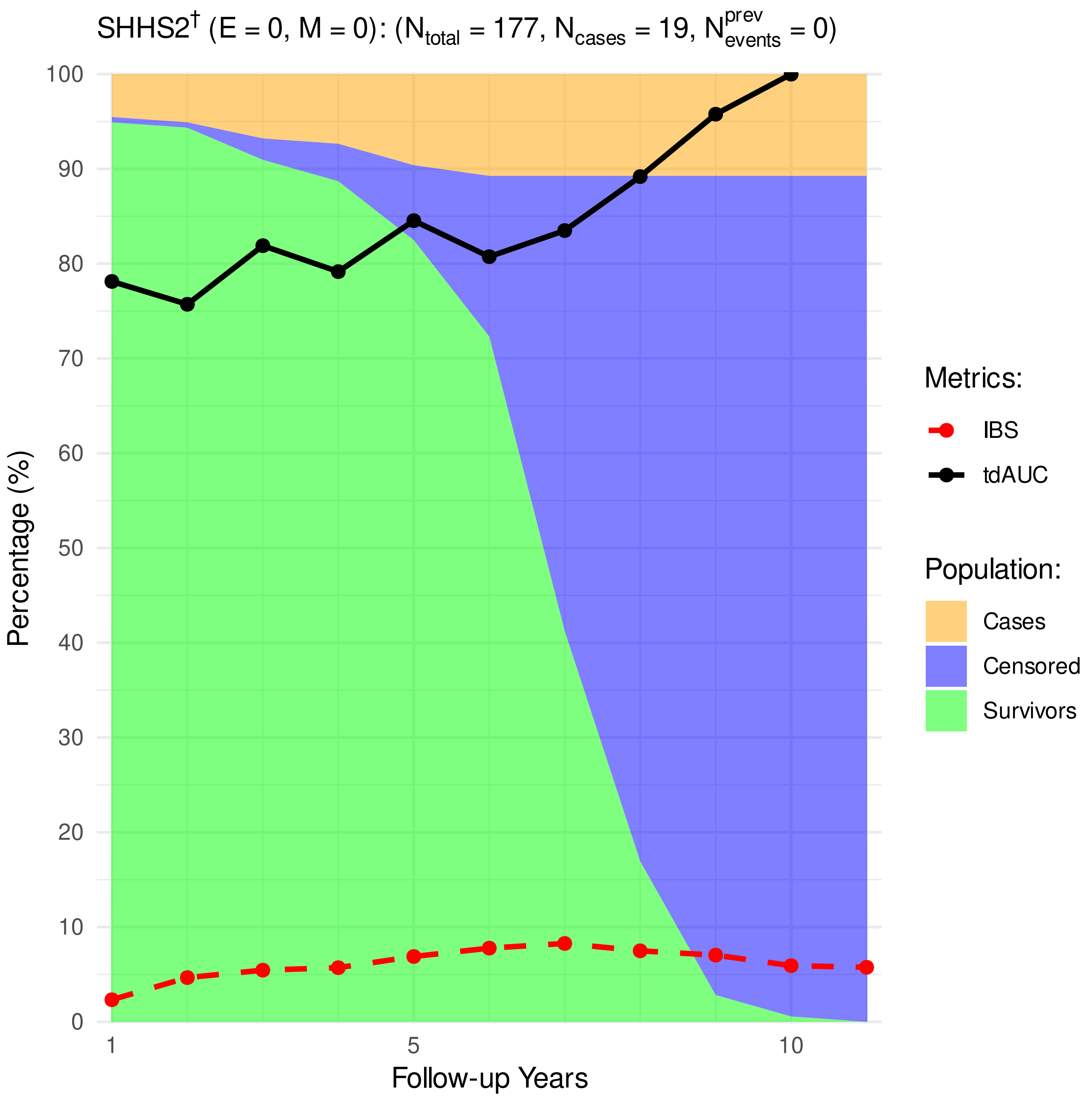


**Figure 9.** Distribution of cardiovascular cases, survivors, and censoring, as well as the performance metrics (time-dependent Area Under Receiver Operating Characteristic [tdAUROC] curve, Integrated Brier Score [IBS].)

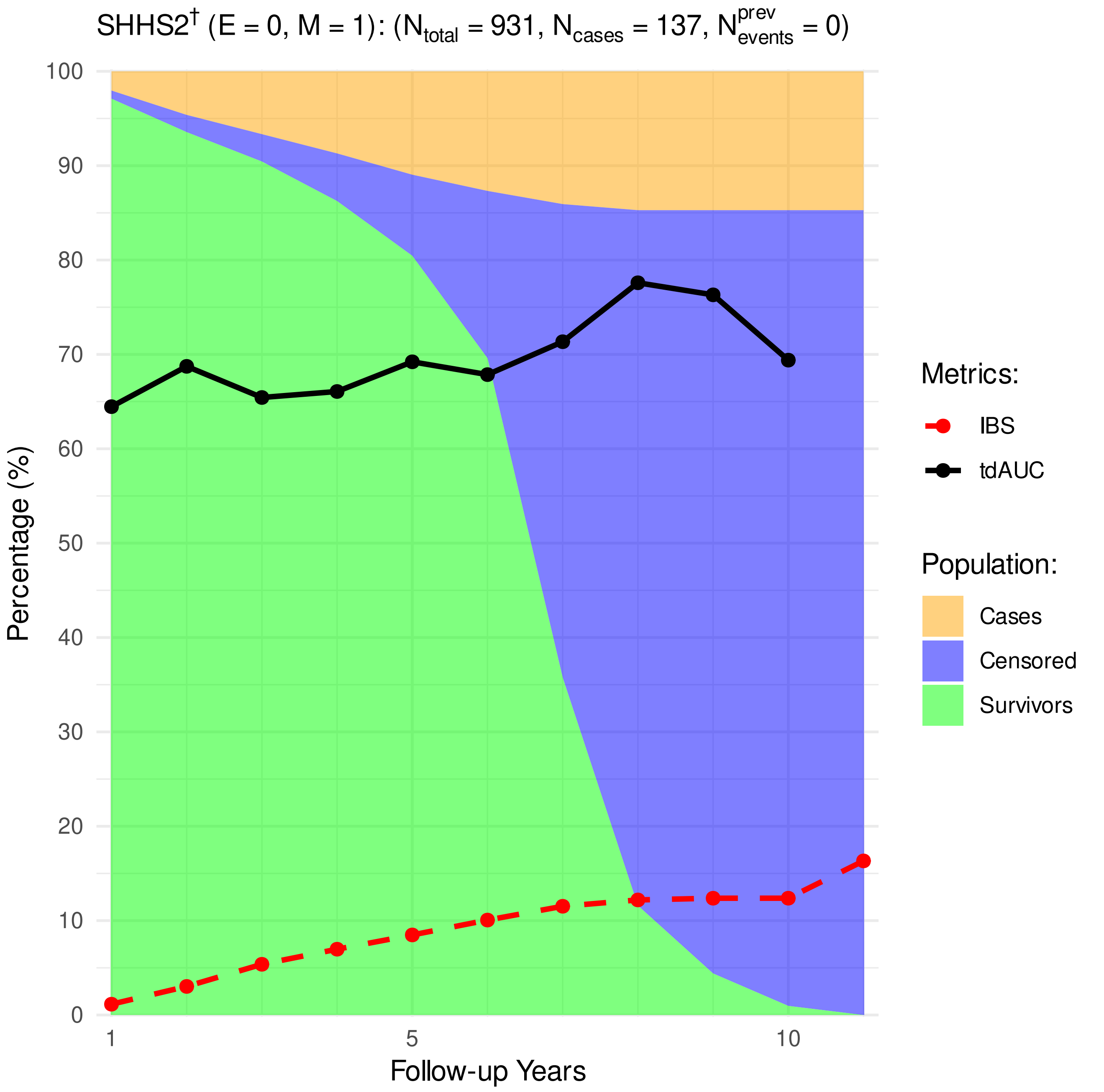


**Figure 10.** Distribution of cardiovascular cases, survivors, and censoring, as well as the performance metrics (time-dependent Area Under Receiver Operating Characteristic [tdAUROC] curve, Integrated Brier Score [IBS].)

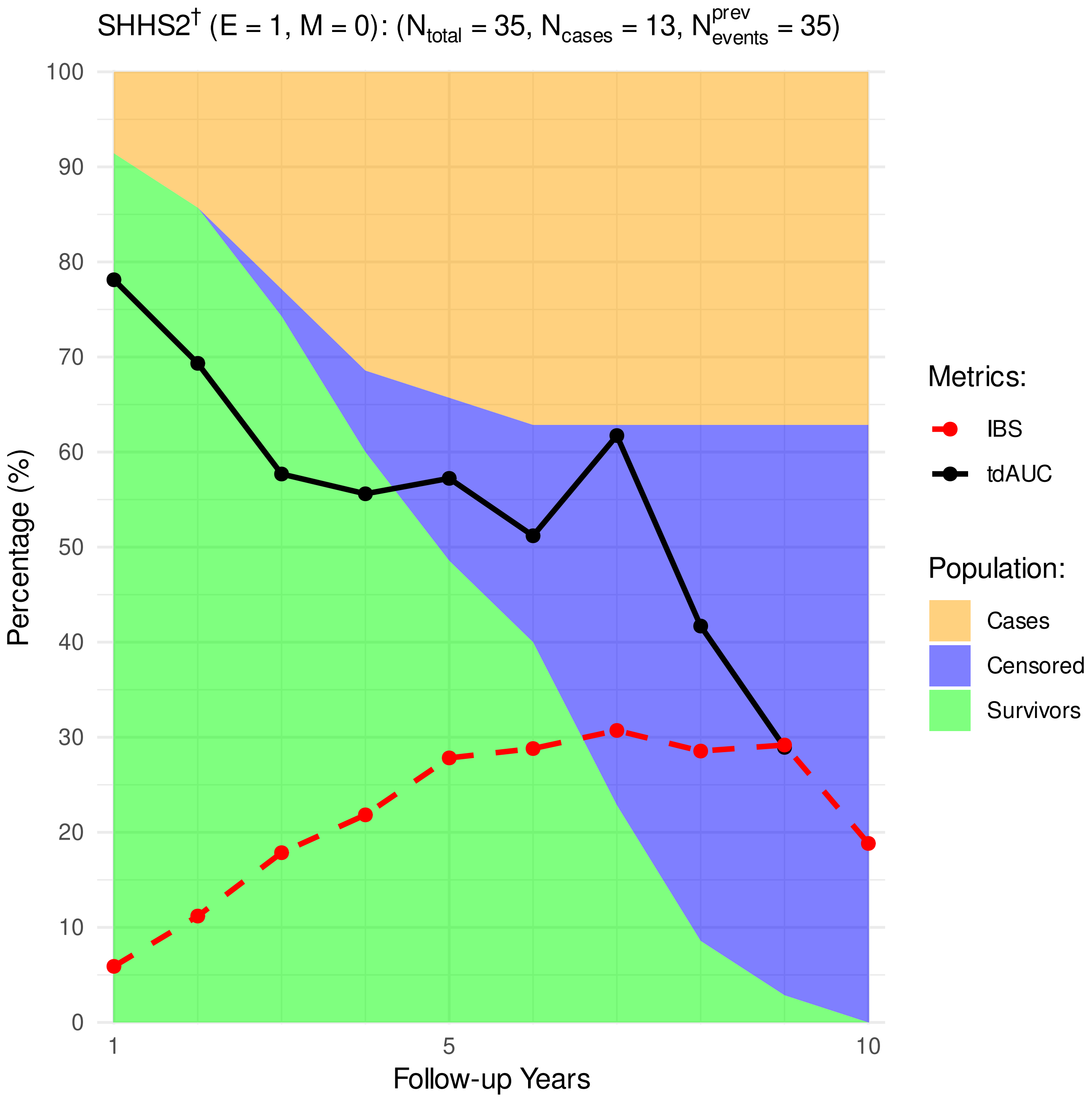


**Figure 11.** Distribution of cardiovascular cases, survivors, and censoring, as well as the performance metrics (time-dependent Area Under Receiver Operating Characteristic [tdAUROC] curve, Integrated Brier Score [IBS].)

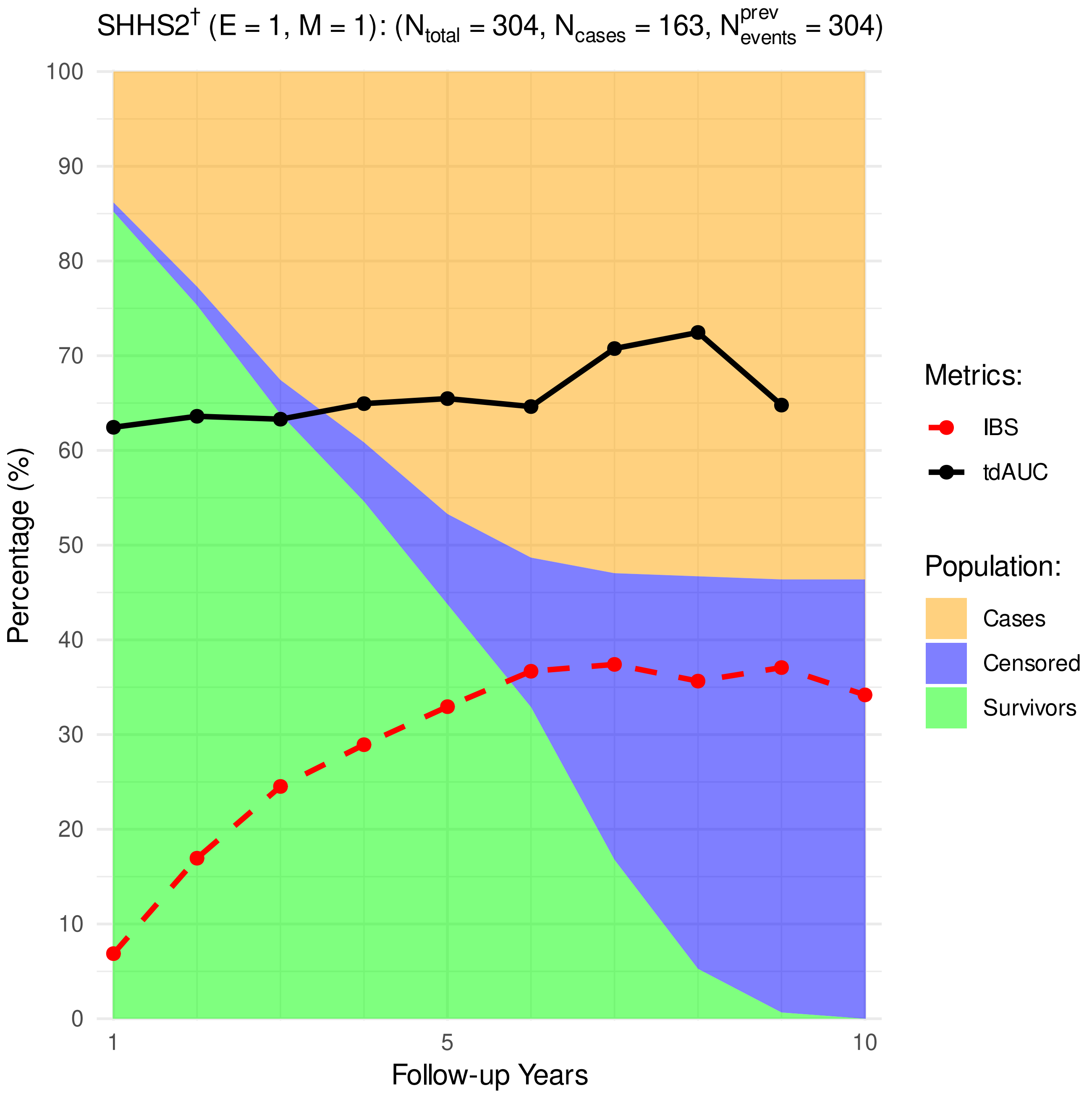


**Figure 12.** Distribution of cardiovascular cases, survivors, and censoring, as well as the performance metrics (time-dependent Area Under Receiver Operating Characteristic [tdAUROC] curve, Integrated Brier Score [IBS].)

#### 6 Survival Plots without AHI predictor

##### 6.1 Primary study cohort

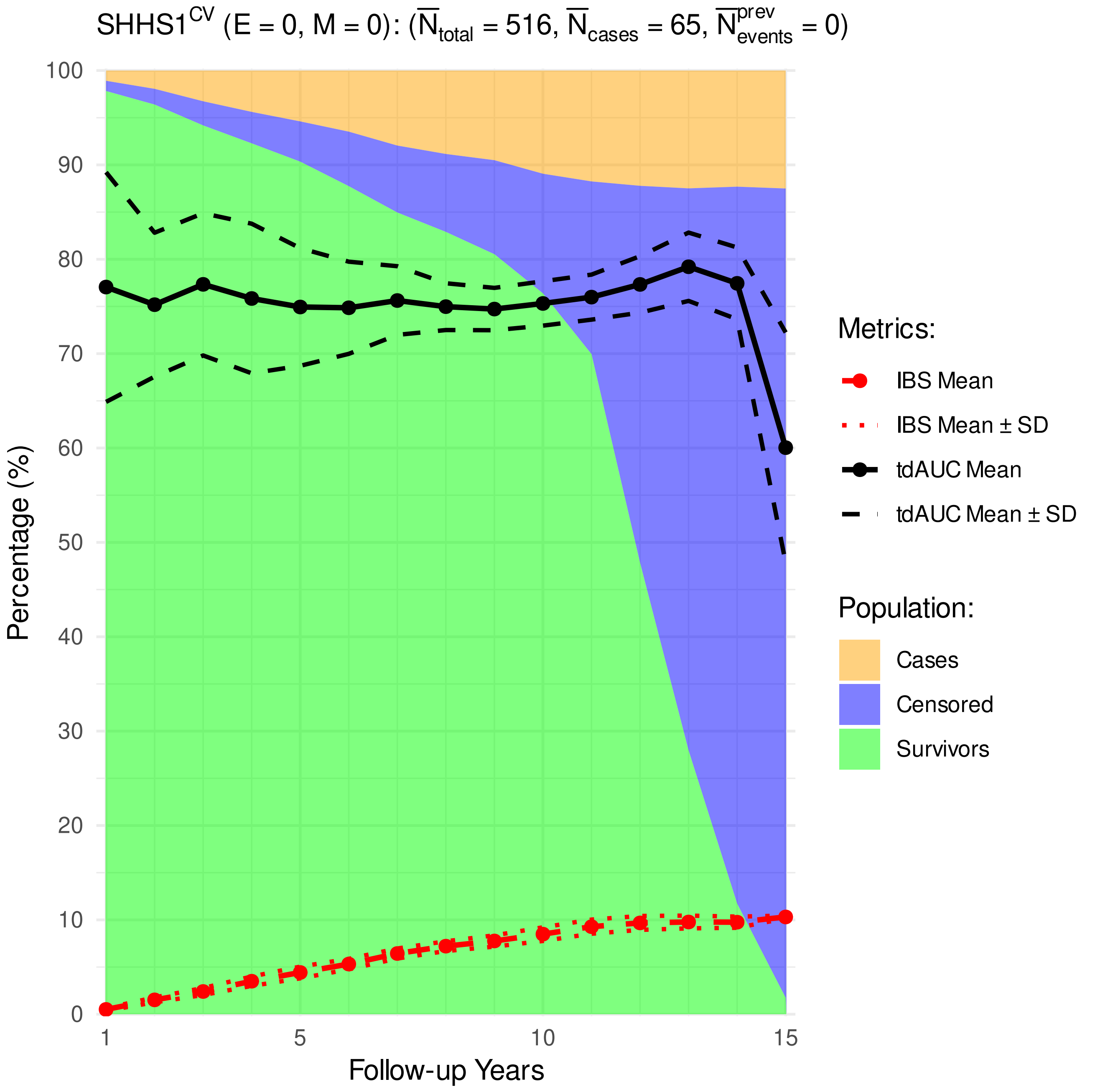


**Figure 13.** Distribution of cardiovascular cases, survivors, and censoring, as well as the performance metrics (time-dependent Area Under Receiver Operating Characteristic [tdAUROC] curve, Integrated Brier Score [IBS].)

##### 6.2 SHHS1 test subjects

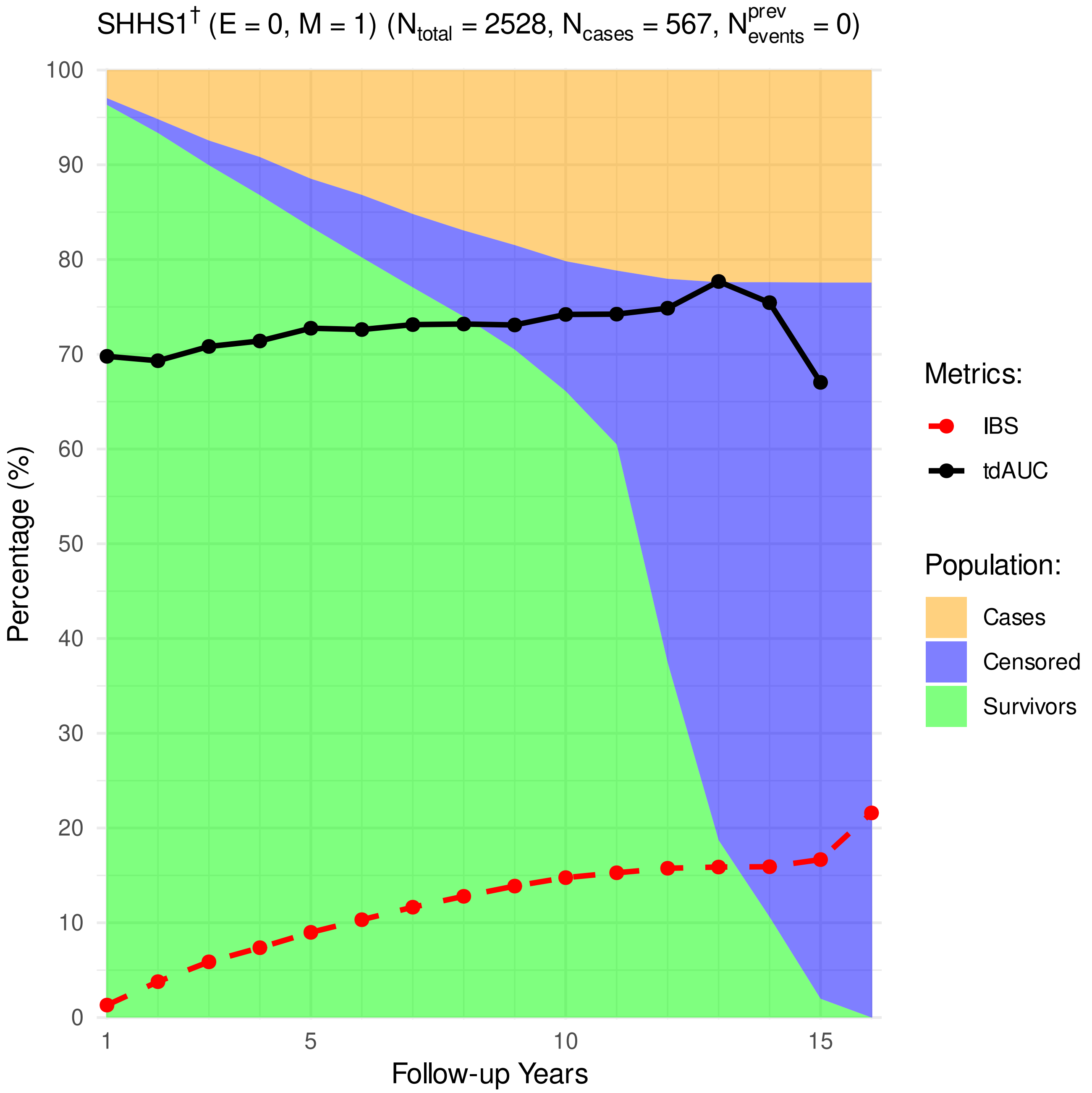


**Figure 14.** Distribution of cardiovascular cases, survivors, and censoring, as well as the performance metrics (time-dependent Area Under Receiver Operating Characteristic [tdAUROC] curve, Integrated Brier Score [IBS].)

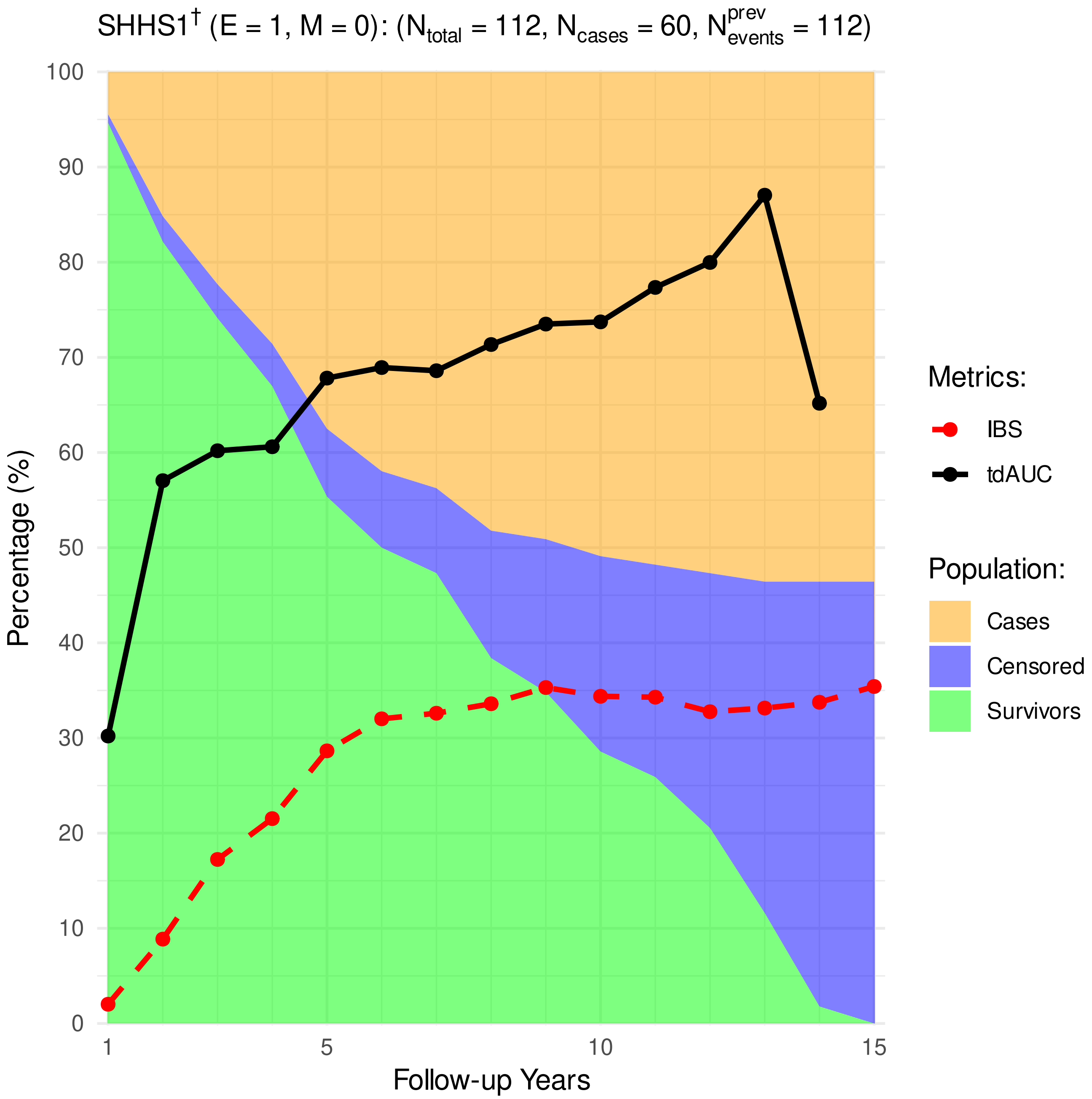


**Figure 15.** Distribution of cardiovascular cases, survivors, and censoring, as well as the performance metrics (time-dependent Area Under Receiver Operating Characteristic [tdAUROC] curve, Integrated Brier Score [IBS].)

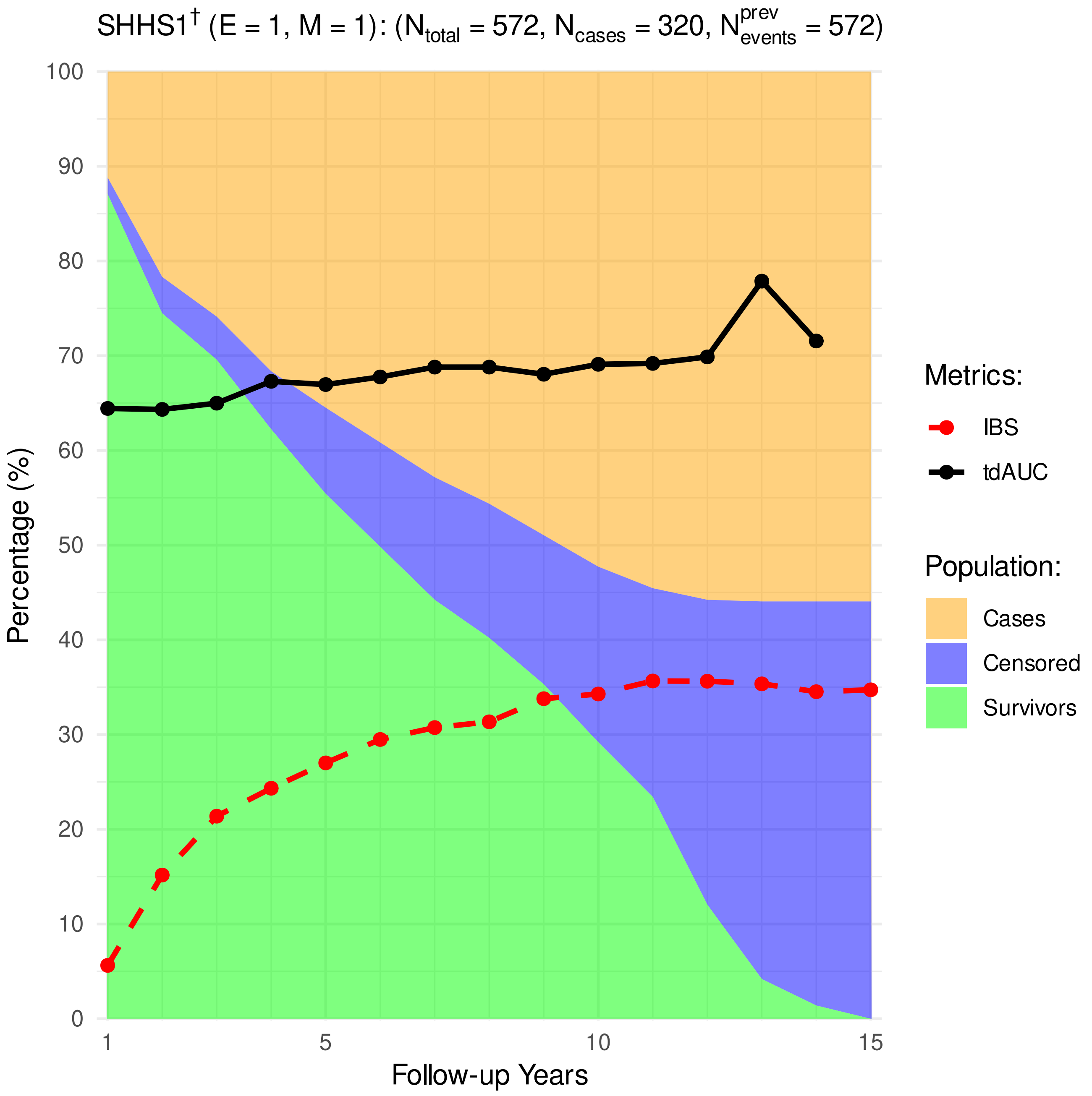


**Figure 16.** Distribution of cardiovascular cases, survivors, and censoring, as well as the performance metrics (time-dependent Area Under Receiver Operating Characteristic [tdAUROC] curve, Integrated Brier Score [IBS].)

##### 6.3 SHHS2 train subjects

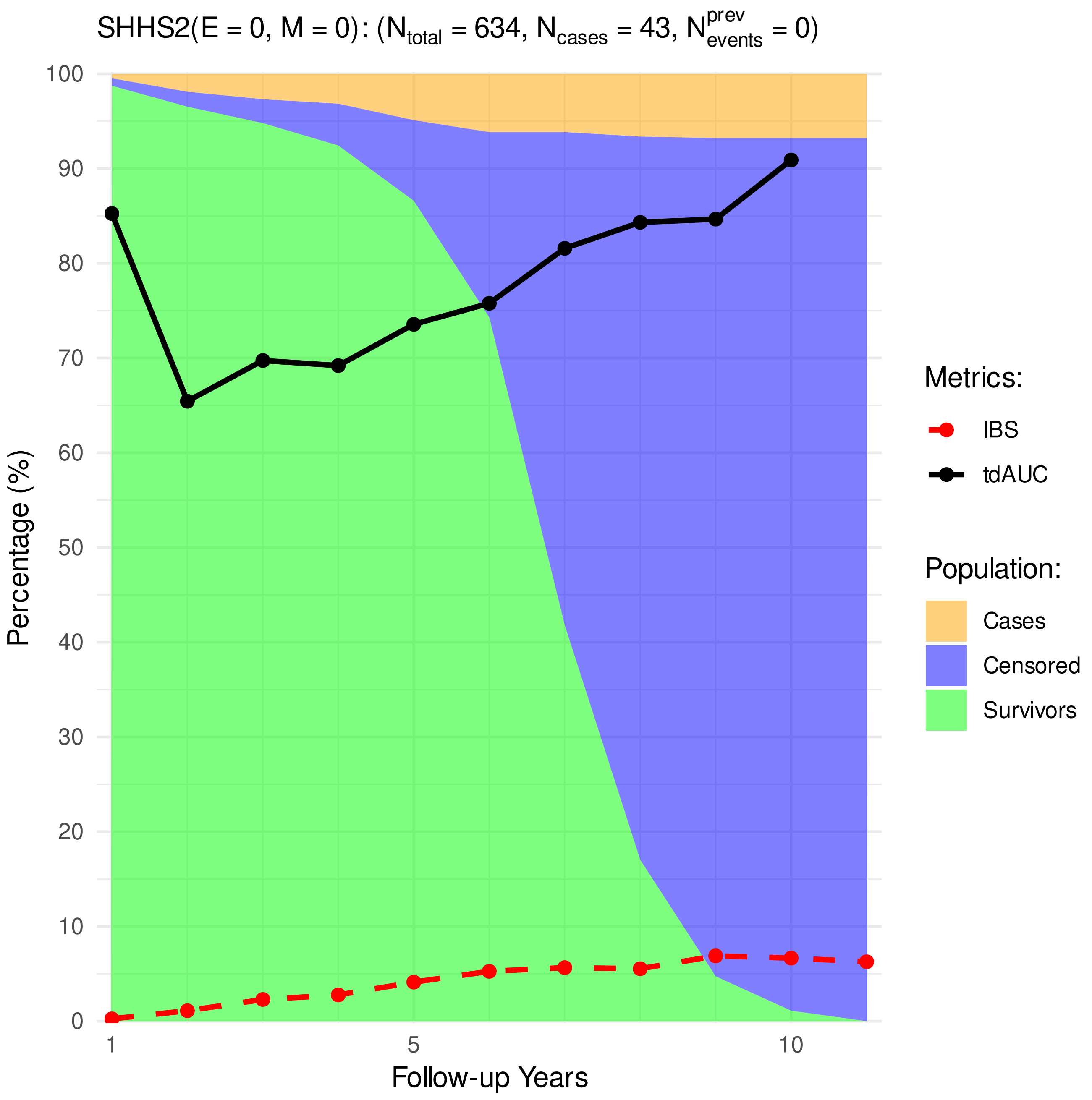


**Figure 17.** Distribution of cardiovascular cases, survivors, and censoring, as well as the performance metrics (time-dependent Area Under Receiver Operating Characteristic [tdAUROC] curve, Integrated Brier Score [IBS].)

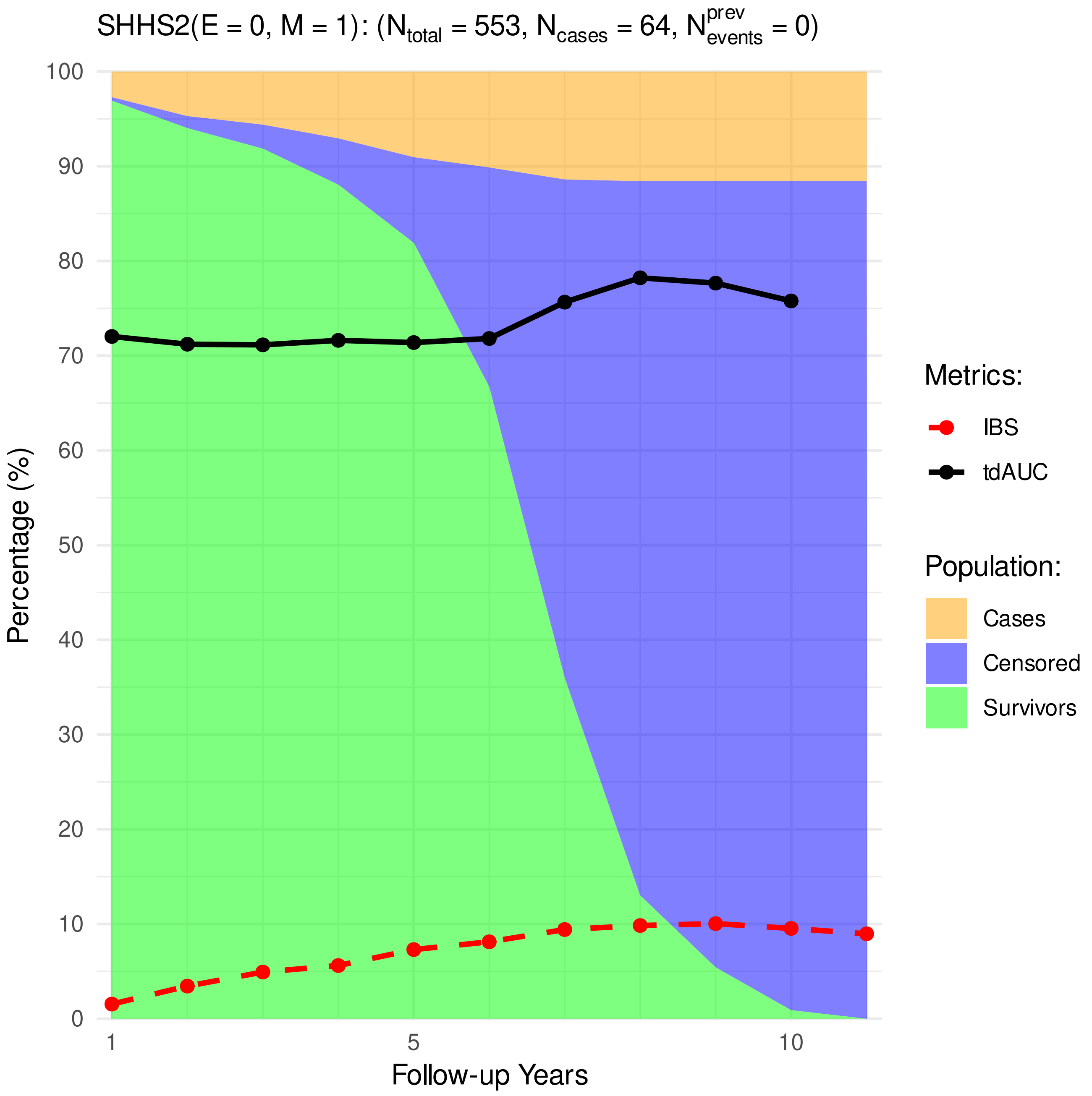


**Figure 18.** Distribution of cardiovascular cases, survivors, and censoring, as well as the performance metrics (time-dependent Area Under Receiver Operating Characteristic [tdAUROC] curve, Integrated Brier Score [IBS].)

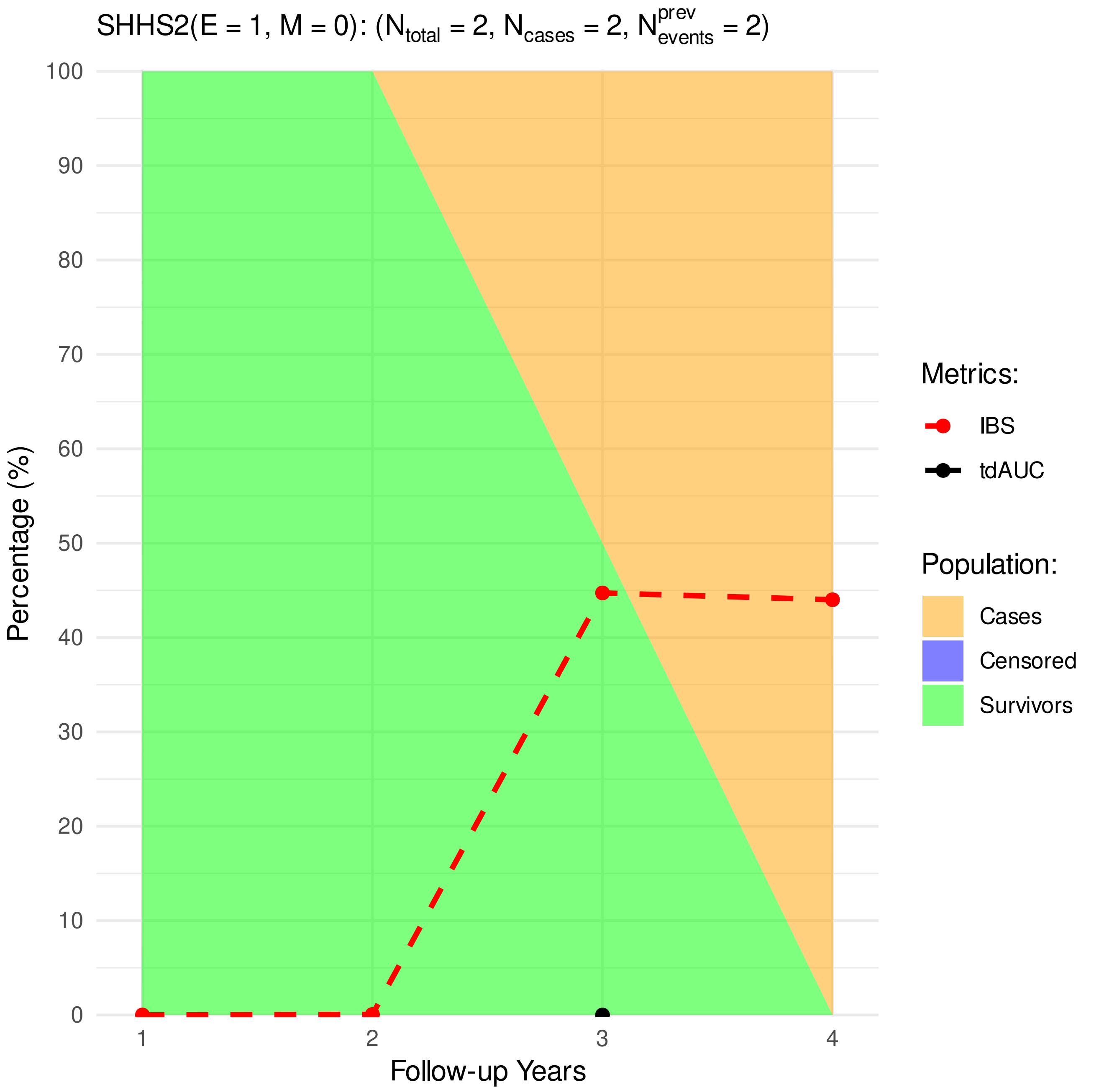


**Figure 19.** Distribution of cardiovascular cases, survivors, and censoring, as well as the performance metrics (time-dependent Area Under Receiver Operating Characteristic [tdAUROC] curve, Integrated Brier Score [IBS].)

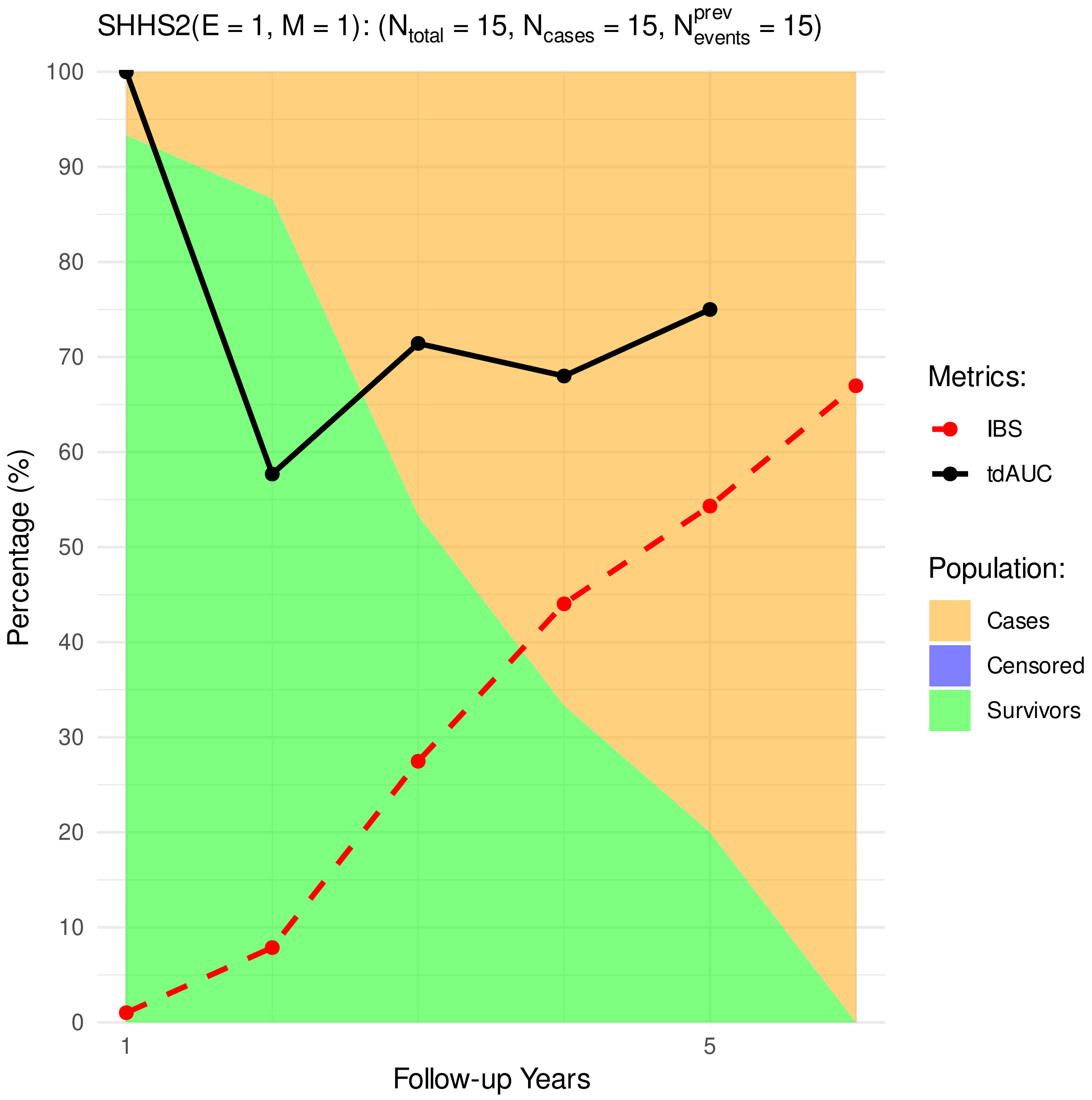


**Figure 20.** Distribution of cardiovascular cases, survivors, and censoring, as well as the performance metrics (time-dependent Area Under Receiver Operating Characteristic [tdAUROC] curve, Integrated Brier Score [IBS].)

#### 7 SHHS2 test subjects

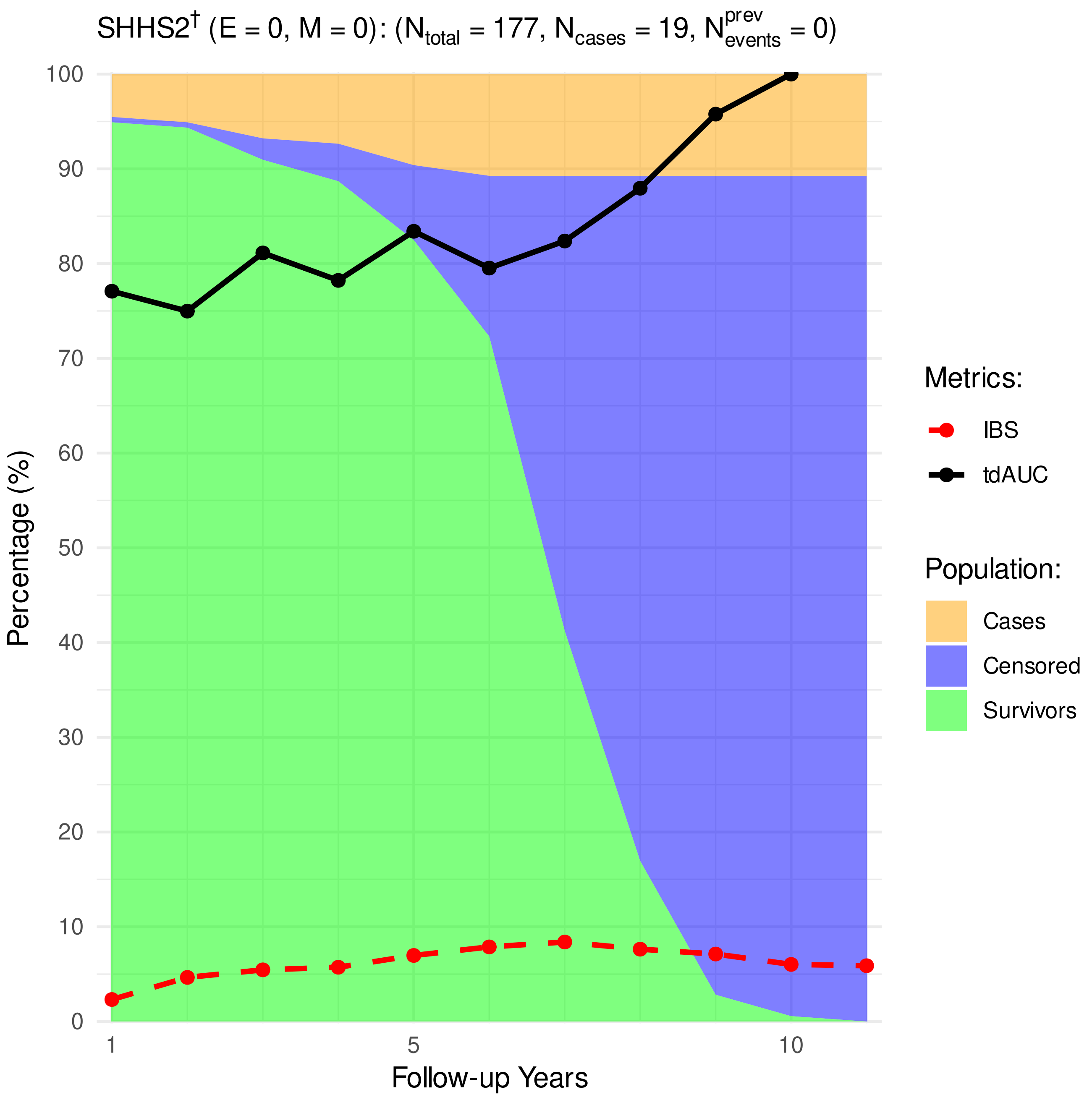


**Figure 21.** Distribution of cardiovascular cases, survivors, and censoring, as well as the performance metrics (time-dependent Area Under Receiver Operating Characteristic [tdAUROC] curve, Integrated Brier Score [IBS].)

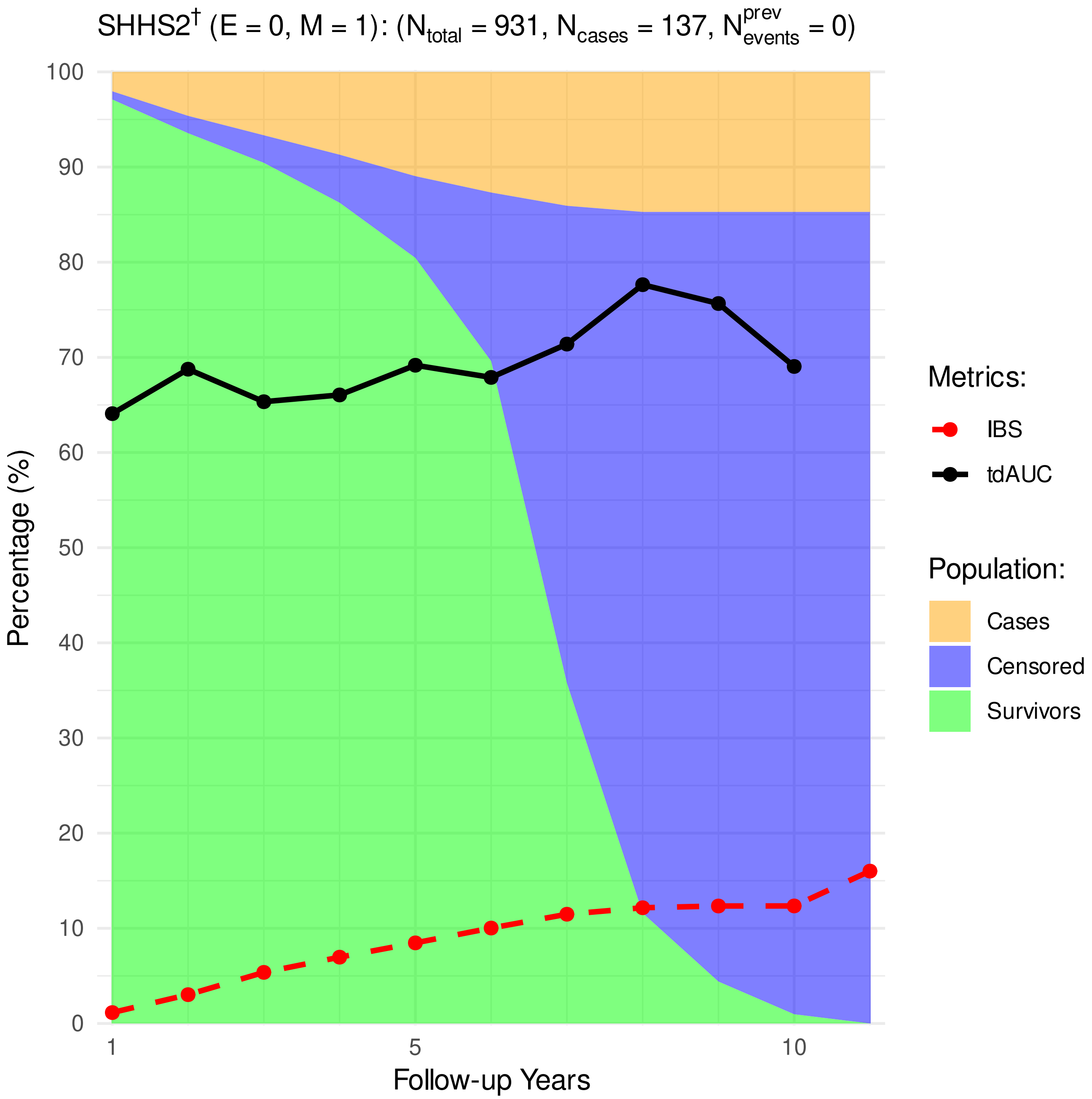


**Figure 22.** Distribution of cardiovascular cases, survivors, and censoring, as well as the performance metrics (time-dependent Area Under Receiver Operating Characteristic [tdAUROC] curve, Integrated Brier Score [IBS].)

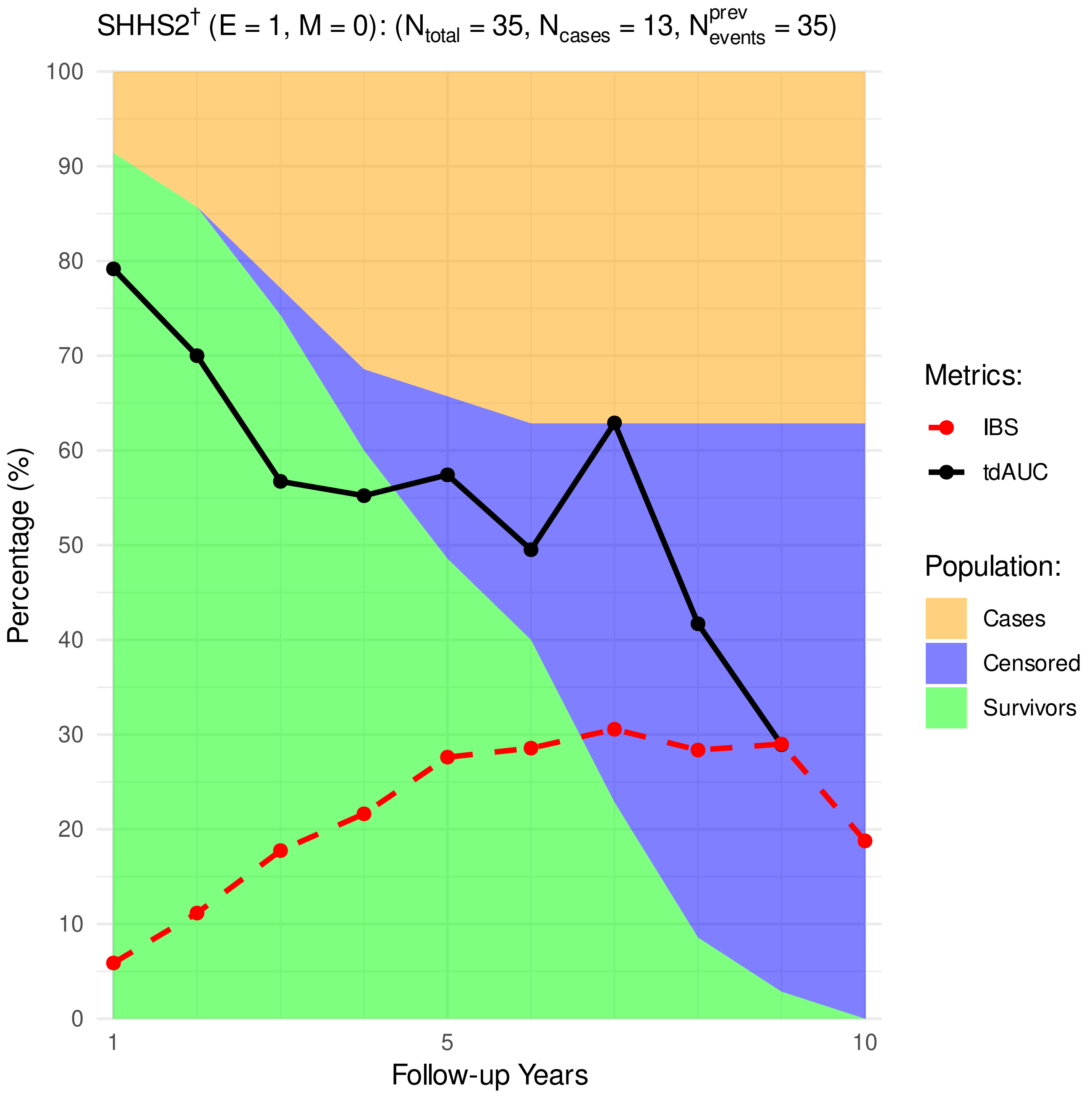


**Figure 23.** Distribution of cardiovascular cases, survivors, and censoring, as well as the performance metrics (time-dependent Area Under Receiver Operating Characteristic [tdAUROC] curve, Integrated Brier Score [IBS].)

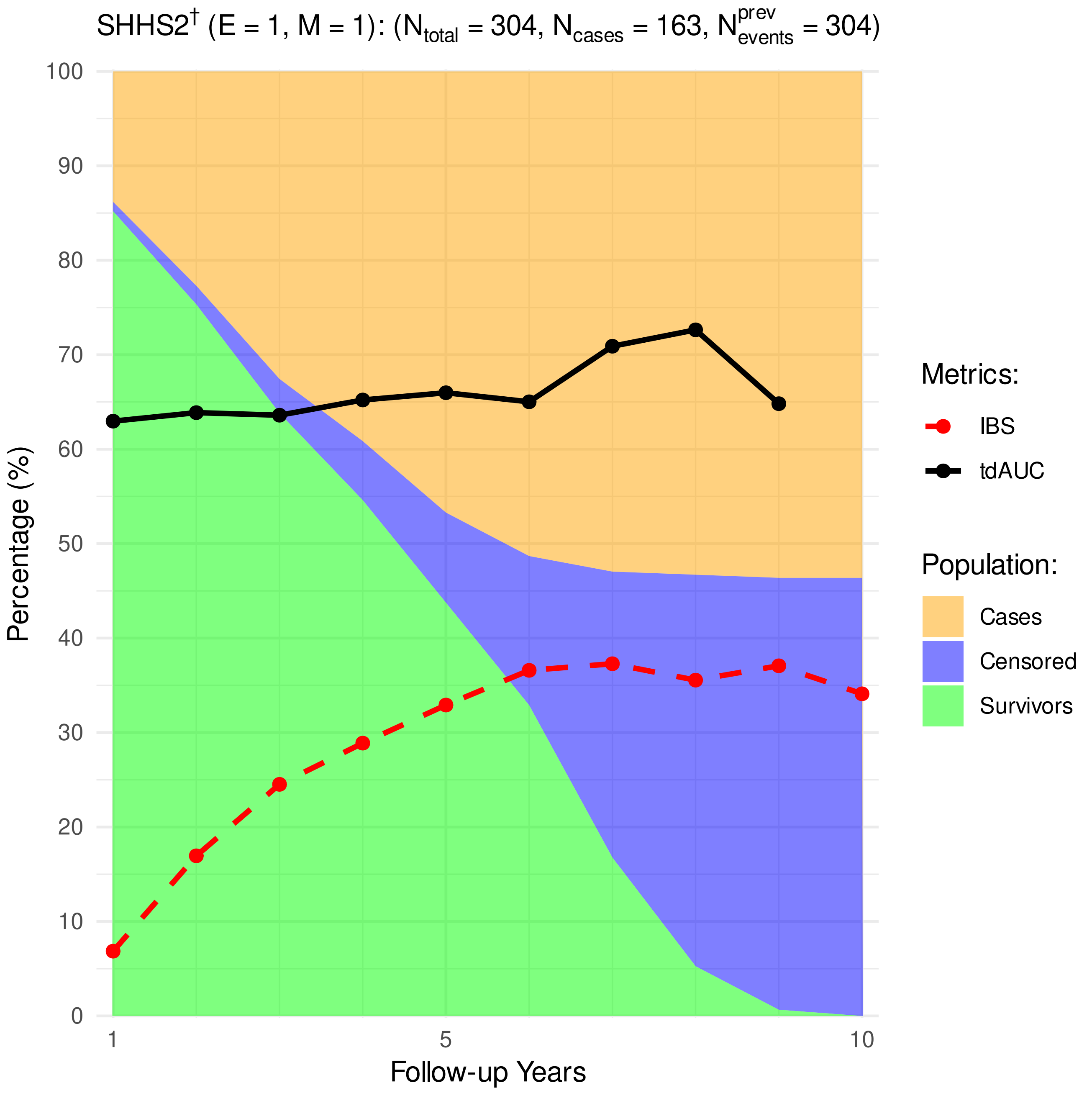


**Figure 24.** Distribution of cardiovascular cases, survivors, and censoring, as well as the performance metrics (time-dependent Area Under Receiver Operating Characteristic [tdAUROC] curve, Integrated Brier Score [IBS].)

#### 8 Partial Effects for RSF without AHI predictor

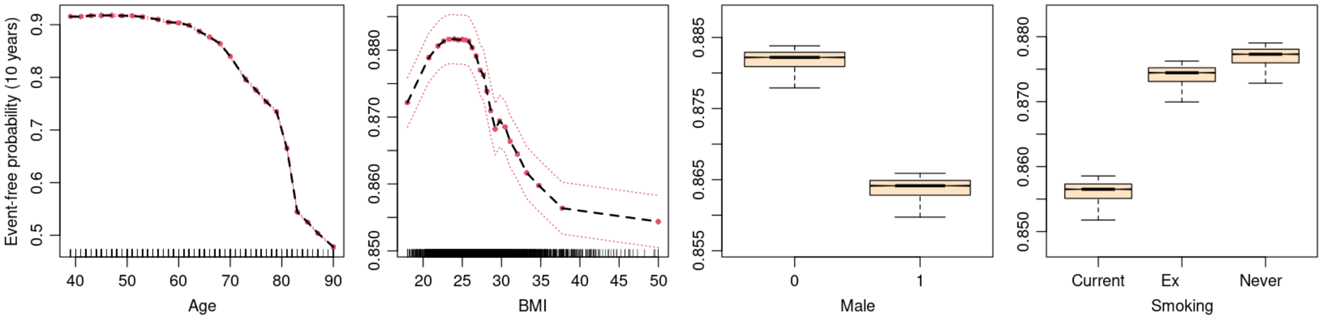


**Figure 25.** Partial effects and their 95% CIs for 10-year cardiovascular event-free probability for the age in years, Body Mass Index (BMI), Apnea-Hypopnea Index (AHI), gender (0 = female, 1 = male), and smoking status, for RSF without AHI predictor. Data points for continuous predictors are shown as ticks on the x-axis.

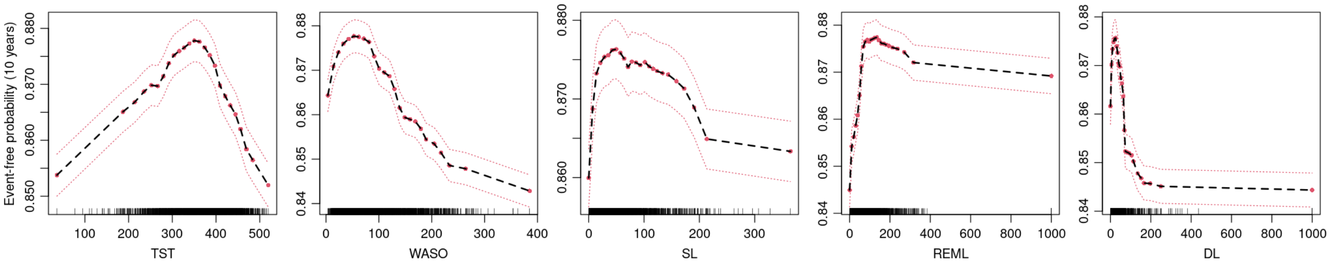


**Figure 26.** Partial effects and their 95% CIs for 10-year cardiovascular event-free probability for the minutes of Total Sleep Time (TST), Wake After Sleep Onset (WASO), Sleep Latency (SL), REM Latency (REM), and Deep-sleep Latency (DL), for RSF without AHI predictor. Data points are shown as ticks on the x-axis.

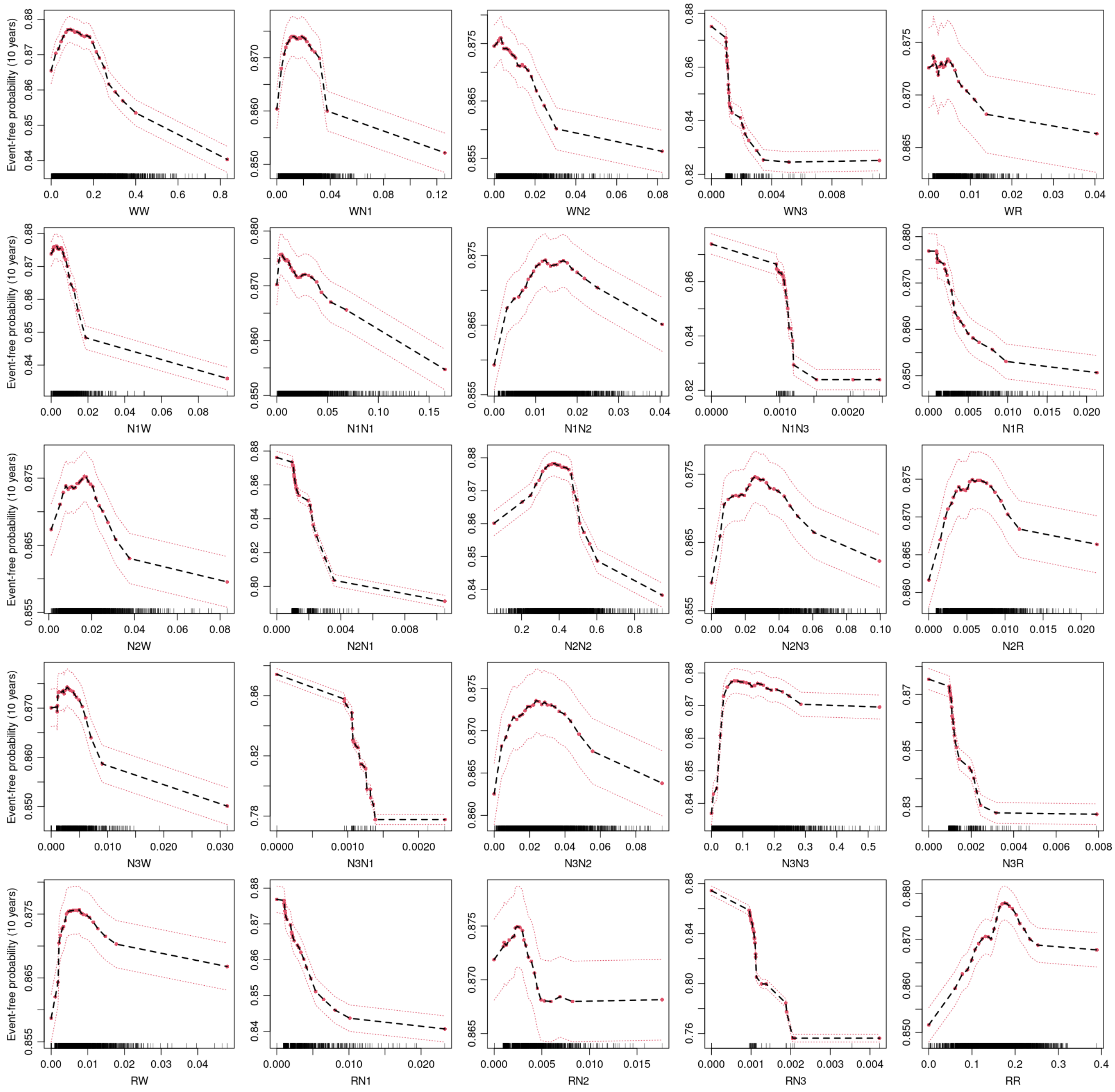


**Figure 27.** Partial effects and their 95% CIs for 10-year cardiovascular event-free probability for the relative frequencies of transitions between sleep-stage (W, N1, N2, N3, REM), for RSF without AHI predictor. Each subplot's x-axis label indicates the transition's direction (e.g., WN1 corresponds to transitions from W to N1). Data points are shown as ticks on the x-axis.
